# Trends in gout management before and after publication of NICE guidance in England: a population-based study in OpenSAFELY

**DOI:** 10.64898/2026.11.05.26367349

**Authors:** Mark D Russell, Brian MacKenna, Samantha Goodliffe, Edward Roddy, Abhishek Abhishek, Shaun Rowark, Katie Bechman, Mark Gibson, Jeremy Brown, Edoardo Cipolletta, Andrea Schaffer, Seb Bacon, Amir Mehrkar, Simon de Lusignan, Laura C Coates, Andrew P Cope, Sam Norton, Ben Goldacre, James B Galloway

## Abstract

**Objectives:** To evaluate gout management in England before and after publication of a national clinical guideline by the National Institute for Health and Care Excellence (NICE) and assess the feasibility of using routinely collected health data to monitor guideline implementation at scale.

**Design:** Observational cohort study with interrupted time-series analysis.

**Setting:** Primary care and hospital admission data, with the approval of NHS England.

**Participants:** 22,618,945 individuals aged 18 to 110 years registered with general practices in England contributing data to the NHS OpenSAFELY Data Analytics Service between July 1, 2016, and June 30, 2025.

**Intervention:** Publication of NICE guidance for the diagnosis and management of gout in June 2022.

**Main outcome measures:** Trends in evidence-based measures for gout management, including new prescription of urate-lowering therapy (ULT) and attainment of a target serum urate below 360 micromol/L. Changes in temporal trends after guideline publication were estimated using interrupted time-series models.

**Results:** Of 245,445 individuals newly diagnosed with gout who had available follow-up, 87,885 (35.8%) were prescribed ULT within 12 months of diagnosis. ULT initiation increased from 26.6% in July 2016 to 43.0% in June 2025; however, the rate of increase slowed after publication of the NICE guideline (change in trend: 0.76 percentage-point decrease per year; 95% CI 0.35 to 1.17; p<0.001). Among 108,380 people initiating ULT at any point after diagnosis, 35,865 (33.1%) had at least one recorded serum urate level below target within 12 months of initiation. Urate target attainment improved modestly following publication of the NICE guideline (change in trend: 1.25 percentage-point increase per year; 95% CI 0.64 to 1.86; p<0.001), although sensitivity analyses suggested that was a continuation of an existing trend rather than a guideline-specific effect. Younger adults and males were more likely to be prescribed ULT but less likely to achieve urate targets than older adults and females.

**Conclusions:** Substantial gaps remain between observed gout management and NICE guideline recommendations, particularly for ULT initiation and serum urate target attainment. Guideline publication was not associated with marked improvements beyond existing temporal trends, highlighting the need for implementation strategies to improve gout care. Importantly, routinely collected health data can be used to monitor guideline implementation at scale and identify priorities for improvement.

## Introduction

Gout is a common and debilitating form of inflammatory arthritis caused by the deposition of monosodium urate crystals in the joints and other tissues.^1^ The crystal burden in gout can be reduced through sustained urate-lowering therapy (ULT, e.g. allopurinol or febuxostat), titrated to achieve a serum urate concentration below 360 micromol/L, or below 300 micromol/L in people with tophaceous gout.^2–4^ Treat-to-target ULT can prevent recurrent gout flares and irreversible joint damage, promote resolution of tophi, and improve quality of life.^1^ Increasing evidence from observational studies also suggests that treat-to-target ULT is associated with a lower risk of major adverse cardiovascular events and progression of chronic kidney disease (CKD).^5,6^

Despite ULT being available for over 60 years, earlier population-based studies from the UK and many other countries had shown persistently low rates of ULT initiation and serum urate target attainment.^7–9^ Recognising the need for improvement, the National Institute for Health and Care Excellence (NICE) published its first guideline on the diagnosis and management of gout in June 2022.^2^ NICE guidance is of particular relevance to primary care clinicians, where most people with gout are managed in the UK.^2^ NICE recommendations include offering ULT to people with gout who have multiple or troublesome flares, CKD stages 3 to 5, diuretic therapy, chronic gouty arthritis or tophaceous gout; discussing the option of ULT with people with gout who are not within these groups; titrating the dose of ULT to achieve a serum urate target; and consideration of prophylactic use of colchicine, NSAIDs or corticosteroids during ULT initiation and titration.^2^

It remains unknown whether these recommendations have led to improvements in gout care. This reflects broader challenges in evaluating the implementation of guidelines across healthcare systems. Current approaches rely heavily on local clinical audit and manual data collection, which is resource intensive and limits the ability to identify unwarranted variation at a national level.^10,11^ Access to electronic health records in primary care has the potential to overcome many of these limitations. The NHS OpenSAFELY Data Analytics Service, a Trusted Research Environment run by NHS England,^12^ provides secure access to these data, creating an opportunity for continuous monitoring of guideline implementation and quality of care across a wide range of clinical conditions.

Our objective was to use data in OpenSAFELY to evaluate gout management in England before and after publication of NICE guidance, assess variation across demographic groups, and demonstrate a scalable approach to monitoring guideline implementation using routinely collected health data.

## Methods

### Study design

We conducted a population-level, observational cohort study using the NHS OpenSAFELY Data Analytics Service. This service provides secure access to full, coded GP records and linked hospital admission data for patients registered with general practices in England using SystmOne (TPP) software, representing approximately 40% of the population of England.^12^ There is substantial variation in the regional coverage of SystmOne practices across England, although their registered patients are representative of the general population of England with respect to age, sex, socioeconomic status, ethnicity and causes of death.^13^ With support from the Royal College of General Practitioners and the British Medical Association, the Department of Health and Social Care expanded the use of OpenSAFELY for research beyond COVID-19 in 2025.^14,15^

The reference population for this study consisted of all individuals aged 18 to 110 years who were registered for at least 12 months with SystmOne general practices in England at any point between 1 July 2016 and 30 June 2025. Individuals with type 1 opt-outs recorded with their general practice were excluded, indicating that they did not consent to personal data being shared with organisations outside their direct care (approximately 3% of the population of England in 2017).^16,17^

### Case definition

Incident gout diagnoses were defined as individuals from the reference population who had an index diagnostic code for gout in their primary care record or a hospitalisation with a primary discharge diagnosis of gout during the study period (see Supplementary Appendix for codelists).^18^ Individuals were required to have at least 12 months of continuous practice registration before their index diagnosis to minimise the likelihood of misclassifying prevalent cases as incident. Individuals who had received a prescription for ULT more than 30 days before their index diagnostic code for gout were deemed not to be incident diagnoses.

### Outcomes

Outcomes were selected *a priori* to reflect NICE recommendations for gout management that were measurable using data available in OpenSAFELY, as follows: the proportion of individuals with index gout diagnoses who: i) had a serum urate level recorded within 2 months before or after their index diagnosis, provided this was before ULT initiation; ii) were prescribed ULT (allopurinol or febuxostat) within 12 months of diagnosis; iii) had at least one recorded risk factor indicating a need to be offered ULT (recurrent gout flares, CKD stages 3 to 5, diuretic use, chronic gouty arthritis or tophaceous gout)^2^ and who were prescribed ULT within 12 months of risk factor onset (or within 12 months of gout diagnosis if risk factors were present at diagnosis); iv) received allopurinol versus febuxostat as their first urate-lowering medication; v) were initially prescribed higher dose allopurinol (300 mg tablets) or febuxostat (120 mg tablets) as their first ULT prescription; vi) were prescribed febuxostat with pre-existing major cardiovascular disease (coronary heart disease, stroke or transient ischaemic attack); vii) were prescribed colchicine, NSAIDs or corticosteroids on the same day as their first ULT prescription, with a further prescription 3 months later (used as a proxy for gout flare prophylaxis); viii) were still prescribed ULT 12 months after their initial ULT prescription; ix) had at least one serum urate level, or two or more serum urate levels, recorded within 12 months of initiating ULT; x) had a recorded serum urate concentration below 360 micromol/L within 12 months of ULT initiation; xi) had a repeat serum urate level recorded 6 to 12 months after obtaining a serum urate level below 360 micromol/L; xii) remained below 360 micromol/L on the first repeat serum urate measurement 6 to 12 months after target attainment; xiii) had prescriptions issued for colchicine, NSAIDs or corticosteroids on the same day as a recorded gout flare (see Supplementary Appendix for further details on outcome definitions).

### Statistical analysis

Baseline sociodemographic characteristics and comorbidities were summarised for the reference population at cohort entry and at the time of diagnosis for individuals with gout. Monthly trends in outcome attainment were presented for all individuals with index gout diagnoses, and separately by sex, age group (18-39 years; 40-59 years; 60-79 years; and 80 years and over), socioeconomic status (measured using quintiles of Index of Multiple Deprivation [IMD], ranging from quintile 1 [most deprived] to quintile 5 [least deprived]), and region (categorised into 9 Nomenclature of Territorial Units for Statistics Level 1 regions of England).^12^

Single group interrupted time-series analysis (ITSA) was performed to estimate changes in trends in monthly outcome attainment following publication of the NICE gout management guideline in June 2022.^19^ Annualised trends before and after guideline publication, expressed as percentage-point change per year, were presented alongside the difference in trends (post versus pre-guideline) and the percentage step change occurring in June 2022. Autocorrelation between monthly observations was accounted for using Newey-West standard errors with a bandwidth of 5 months.^19^ Residual autocorrelation was assessed using the Cumby-Huizinga test for autocorrelation,^20^ with robustness examined using alternative lag specifications. Sensitivity analyses included adjustment for calendar-month effects to account for seasonality, and for the COVID-19 pandemic using an indicator for March 2020 to February 2022.

To explore the potential impact of delays in guideline implementation, sensitivity ITSAs were performed using an alternative intervention point of 12 months after NICE guideline publication (June 2023), comparing differences in trends with the primary analysis. Additionally, an intervention point of 12 months before NICE guideline publication (June 2021) was utilised as a negative control to investigate whether changes in trends began before guideline publication.

Data management was conducted using Python 3.14, with statistical analyses performed in Stata version 19 and R version 4.5. Analysis code and codelists are available online (https://github.com/opensafely/nice_gout). To meet statistical disclosure control requirements, all frequency counts were rounded to the nearest five, and non-zero counts fewer than eight were suppressed.

### Study approval and ethics

This work was conducted with the approval of NHS England [POS-2026-3001; https://www.opensafely.org/project/pos-2026-3001] and approved for publication following review by NHS England/Department for Health and Social Care. This study was approved by the King’s College London BDM Research Ethics Panel (LRS/RGO-24/25-50854).

### Patient and public involvement

People with lived experience of gout were involved in the design of this study and will play a central role in developing strategies to improve gout care based upon these findings. More broadly, OpenSAFELY has involved patients and the public in various ways, including a public website, participation in two citizen juries, co-development of an explainer video, patient representation on the OpenSAFELY Oversight Board, and partnering with Understanding Patient Data.^21^

## Results

### Study population and baseline characteristics

From 22,618,945 eligible adults in the OpenSAFELY reference population, there were 262,925 newly recorded gout diagnoses between 1 July 2016 and 30 June 2025. The mean age at diagnosis was 60.7 years (standard deviation [SD] 16.3 years), compared with a mean age of 46.4 years (SD 18.8 years) in the reference population at cohort entry. Individuals with gout were more likely to be male than the reference population (73.9% vs. 49.6%, respectively); more likely to be of White ethnicity (89.2% vs. 79.5%); less likely to be of Asian ethnicity (5.5% vs. 8.3%), Black ethnicity (1.7% vs. 3.1%), mixed ethnicity (0.7% vs. 1.5%), or Chinese or other ethnicity (1.2% vs. 3.0%); and more likely to be from the least deprived socioeconomic quintile (20.1% vs. 17.7%). Individuals with gout were more likely to have comorbidities at baseline than the reference population, including CKD stages 3 to 5 (23.6% vs. 5.1%), hypertension (42.9% vs. 16.3%), obesity (45.9% vs. 25.5%), type 2 diabetes mellitus (14.3% vs. 5.8%), coronary heart disease (12.0% vs. 3.7%), previous stroke or transient ischaemic attack (5.4% vs. 2.1%), or diuretic use (25.3% vs. 6.0%) (Supplementary Table S1).

### Serum urate measurement at diagnosis

Serum urate levels were recorded at diagnosis in 154,585 (58.8%) individuals with gout, with a mean concentration of 464 micromol/L (SD 98.7). The proportion of individuals with baseline serum urate measurements decreased during the early COVID-pandemic, from 57.9% in January 2020 to 27.5% in April 2020, before returning to pre-pandemic trends by August 2020 (Supplementary Figure S1). Individuals 80 years and above were less likely to have baseline serum urate measurements than younger adults (49.8% in individuals aged ≥80 years; 59.8% in 18-39 years; 59.8% in 40-59 years; and 60.7% in 60-79 years; Supplementary Figure S2). Females were more likely than males to have baseline serum urate levels (61.2% vs. 57.9%; Supplementary Figure S3). Trends were comparable by socioeconomic status and region (Supplementary Figures S4 and S5).

### New urate-lowering therapy prescription

Of 262,925 people with incident gout, 245,445 (93.4%) remained continuously registered for at least 12 months after diagnosis. Among these individuals, 87,885 (35.8%) were prescribed ULT within 12 months of diagnosis. ULT initiation increased from 26.6% among individuals diagnosed in July 2016 to 43.0% among those diagnosed in June 2025. A reduction in the rate of increase in ULT initiation was observed after NICE guidance was published (trend before NICE guideline: 1.97 percentage-point increase per year; trend after: 1.21 percentage-point increase per year; trend difference: 0.76 percentage-point decrease per year, 95% CI 0.35 to 1.17, p<0.001) (Supplementary Figure S6).

138,790 (52.8%) individuals with gout had at least one recorded risk factor at diagnosis or during available follow-up for which NICE specifically recommends that ULT should be offered to patients (recurrent flares; CKD stages 3 to 5, diuretic use, chronic gouty arthritis or tophaceous gout). Of these individuals, 60,580 (43.6%) were prescribed ULT within 12 months of risk factor onset, or within 12 months of gout diagnosis if risk factors were present at diagnosis. By month of risk factor onset, ULT initiation increased from 32.4% in July 2016 to 48.7% in June 2025. The rate of increase slowed after NICE guidance was published: trend before NICE guideline: 1.90 percentage-point increase per year; trend after: 0.82 percentage-point increase per year; trend difference: 1.07 percentage-point decrease per year, 95% CI 0.47 to 1.67, p=0.001 (Figure 1A).

**Figure 1.**
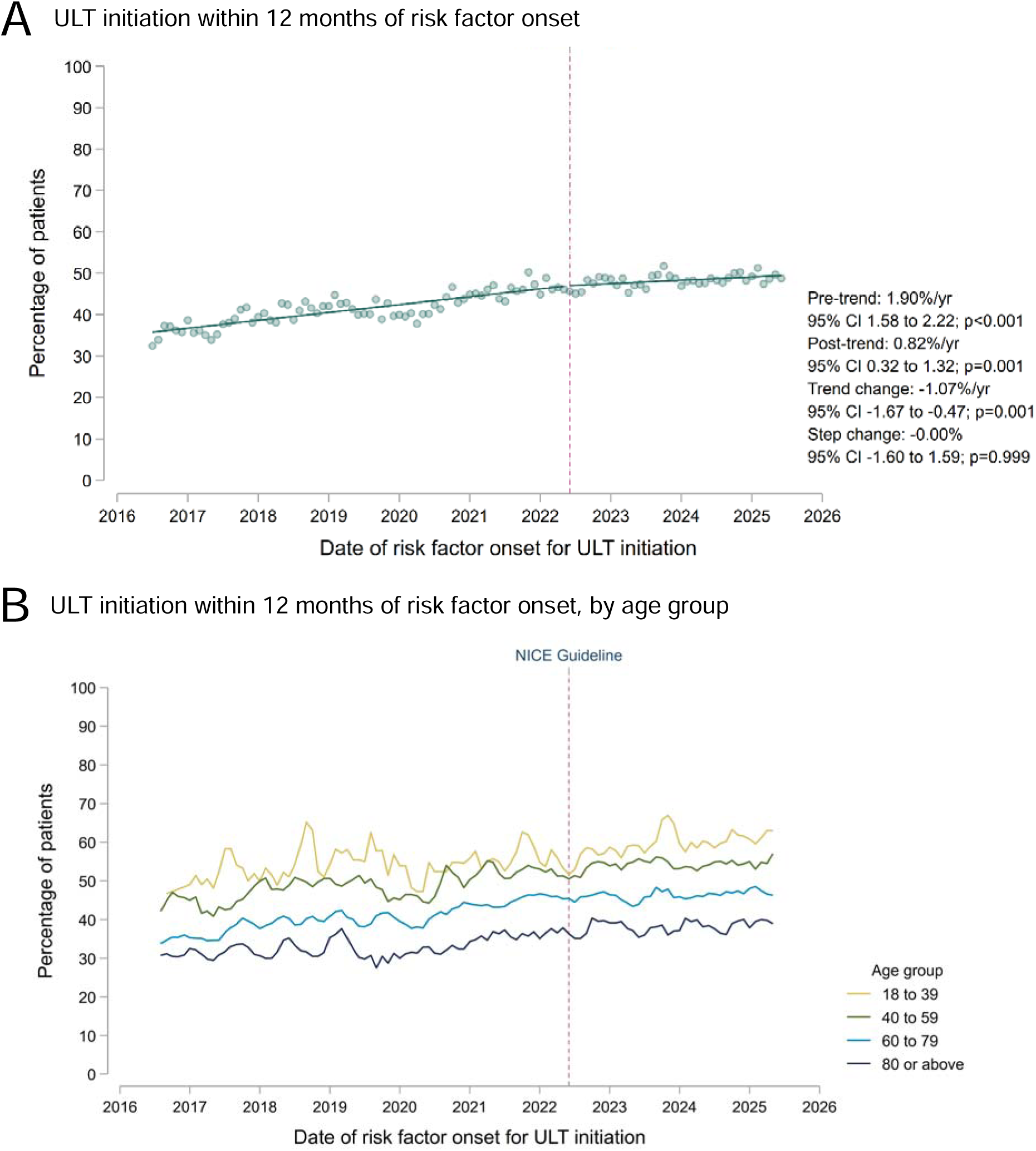
Temporal trends in the initiation of urate-lowering therapy (ULT) within 12 months of onset of risk factors indicating a need to be offered ULT in line with NICE guidance (recurrent gout flares, CKD stages 3 to 5, diuretic use, chronic gouty arthritis or tophaceous gout). The top panel (A) shows trends before and after publication of the NICE Gout Management Guideline in June 2022, fitted using single-group interrupted time-series analysis; annualised coefficients are presented as percentage-point change per year, alongside the change in trends and the immediate step change at the date of publication. The bottom panel shows temporal trends in ULT initiation by age group, with individual trend lines representing 3-monthly rolling averages of the current, preceding and subsequent months.

Older adults were less likely to be prescribed ULT than younger age groups (Figure 1B and Supplementary Figure S7), while males were more likely than females to be initiated on ULT (Supplementary Figure S8). Trends in ULT initiation were comparable by socioeconomic status (Supplementary Figure S9). There was divergence in ULT initiation between regions of England, with rates of ULT initiation declining in London towards the end of the study period (Supplementary Figure S10).

Of 118,555 individuals who initiated ULT at any time during available follow-up, 117,240 (98.9%) were prescribed allopurinol as their first ULT medication, and 1,315 (1.1%) were prescribed febuxostat. The choice of first ULT medication remained stable over the study period (Supplementary Figure S11). The proportion receiving higher dose allopurinol (300 mg tablets) or febuxostat (120 mg tablets) as their first ULT prescription decreased from 17.2% in July 2016 to 5.3% in June 2025 (Figure 2), with a slowing in the rate of reduction observed after NICE guideline publication (Supplementary Figure S12). Similarly, the proportion of individuals initiated on febuxostat who had a pre-existing history of major cardiovascular disease decreased from 21.2% in the year beginning July 2016 to 12.0% in the year beginning June 2025 (Supplementary Figure S13).

**Figure 2.**
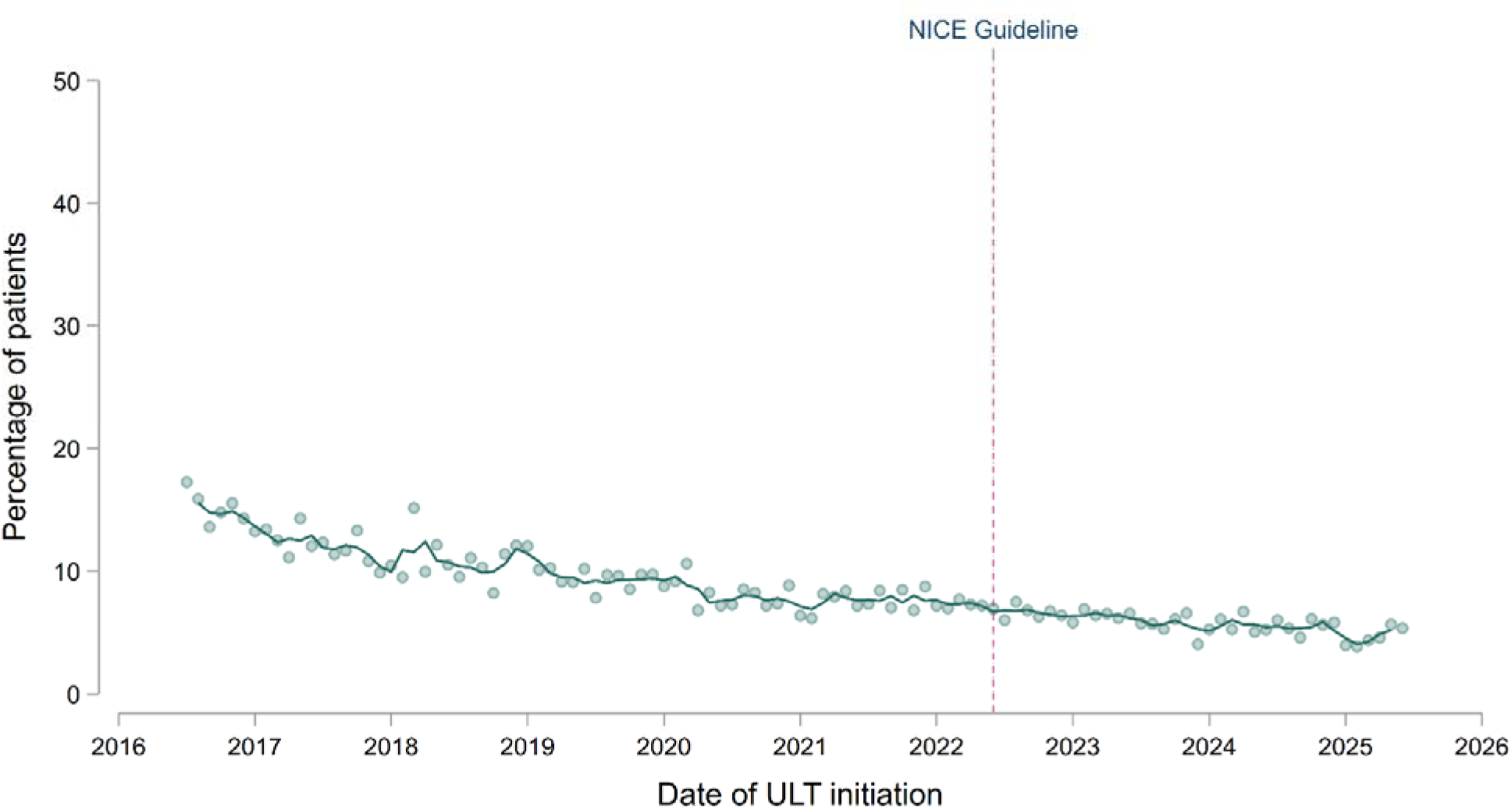
Temporal trends in the proportion of individuals with gout initiating urate-lowering therapy (ULT) who received higher doses of allopurinol (300 mg tablets) or febuxostat (120 mg tablets) as their initial ULT prescription. Individual trend lines represent 3-monthly rolling averages of the current, preceding and subsequent months. The vertical dashed line represents the publication of the NICE Gout Management Guideline in June 2022.

Of 118,555 individuals initiating ULT, 13,135 (11.1%) were prescribed colchicine, NSAIDs or corticosteroids at ULT initiation and again 3 months later, used as a proxy for gout flare prophylaxis. Trends in co-prescription of prophylaxis remained broadly stable over the study period (Supplementary Figures S14 to S17).

Of 108,380 individuals initiating ULT who had available follow-up, 79,125 (73.0%) were still prescribed ULT 12 months after their initial prescription. There were no significant temporal changes in ULT persistence after publication of NICE guidance (Supplementary Figure S18). Adults aged 18 to 39 years were substantially less likely to remain prescribed ULT after 12 months compared with older age groups (56.8% in individuals aged 18-39 years; 70.5% in 40-59 years; 79.1% in 60-79 years; and 77.5% in ≥80 years; Figure 3). Trends in ULT persistence were comparable by sex and socioeconomic status (Supplementary Figures S19 and S20), with lower rates of ULT persistence in London relative to other regions (Supplementary Figure S21). The proportion of individuals initiated on allopurinol who remained on allopurinol at 12 months was higher than for febuxostat (71.9% vs. 50.6%, respectively; Supplementary Figure S22), although the number of people prescribed febuxostat was substantially lower than allopurinol with more variation on a month-by-month basis.

**Figure 3.**
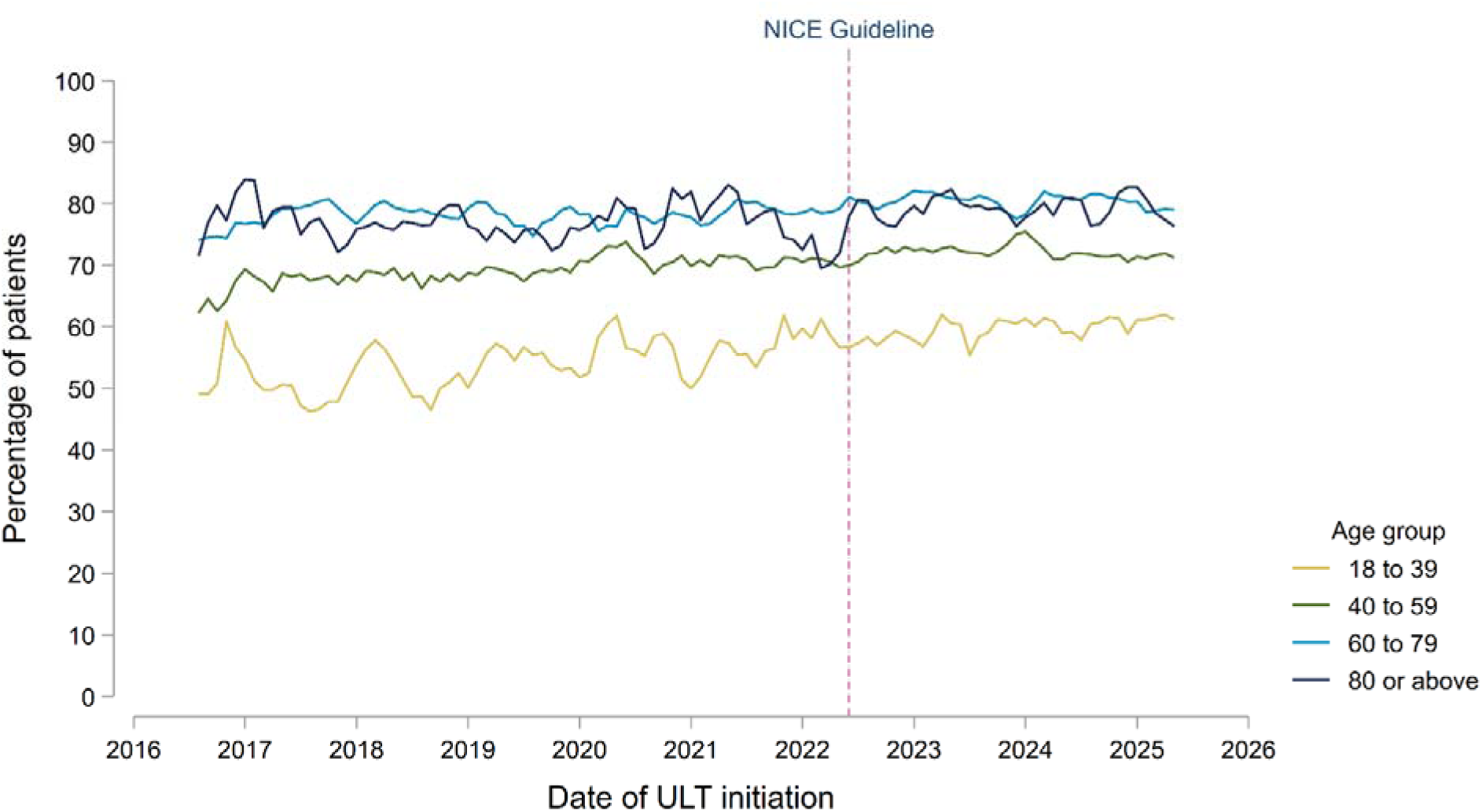
Temporal trends in the proportion of individuals with gout who were still prescribed urate-lowering therapy (ULT) at 12 months after initiation, separated by age group. Individual trend lines represent 3-monthly rolling averages of the current, preceding and subsequent months. The vertical dashed line represents the publication of the NICE Gout Management Guideline in June 2022.

### Urate monitoring and target attainment

Of 108,380 individuals with 12 months of available follow-up after initiating ULT, 73,235 (67.6%) had at least one serum urate measurement during this period, while 35,560 (32.8%) had two or more urate levels recorded. Post-ULT urate monitoring decreased temporarily during the early pandemic, followed by an increase after the NICE guideline (Supplementary Figure S23), which was attenuated after adjustment for the effects of the COVID-19 pandemic (Supplementary Table S2).

Among 108,380 individuals initiating ULT, 35,865 (33.1%) had at least one recorded serum urate level below 360 micromol/L within 12 months of initiation. Target attainment increased modestly towards the end of the study period, from 29.6% in July 2016 to 36.9% in June 2025, with a significant increase after publication of the NICE guideline (trend before NICE guideline: 0.03 percentage-point decrease per year; trend after: 1.22 percentage-point increase per year; trend difference: 1.25 percentage-point increase per year, 95% CI 0.64 to 1.86, p<0.001) (Figure 4A). When restricted to the 73,235 individuals with at least one recorded serum urate measurement within 12 months of ULT initiation, target attainment was 49.0%, with a comparable post-guideline increase (Supplementary Figure S24).

**Figure 4.**
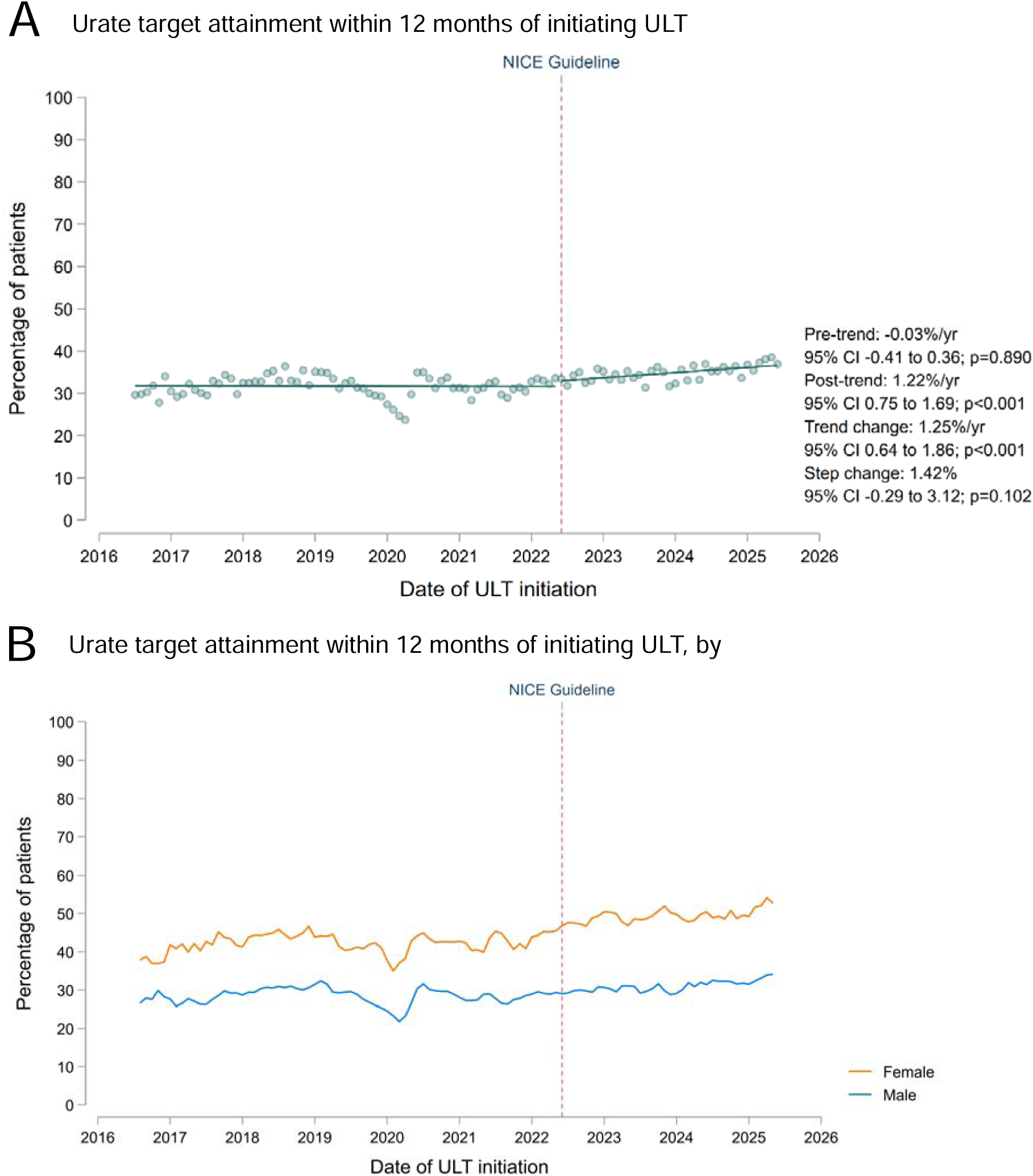
Temporal trends in the attainment of serum urate levels below 360 micromol/L within 12 months of urate-lowering therapy (ULT) initiation. The top panel (A) shows trends before and after publication of the NICE Gout Management Guideline in June 2022, with individual data points representing the mean monthly proportion of individuals achieving urate targets; trend lines were fitted using single-group interrupted time-series before and after publication of the NICE guideline, with annualised coefficients presented as percentage-point change per year, alongside the change in trends and the immediate step change at the date of publication. The bottom panel (B) shows temporal trends separated by sex, with 3-monthly rolling averages of the current, preceding and subsequent months.

Females were substantially more likely to achieve target than males (45.0% vs. 29.6%, respectively; Figure 4B). Younger adults were less likely to achieve target than older adults (18.0% in individuals aged 18-39 years; 29.8% in 40-59 years; 40.2% in 60-79 years; and 35.7% in ≥80 years; Supplementary Figure S25). Individuals from the most deprived socioeconomic quintile were less likely to achieve target than those from the least deprived quintile (28.8% vs. 37.3%, respectively; Supplementary Figure S26). Urate target attainment was broadly comparable by region (Supplementary Figure S27).

Of 48,155 individuals achieving target at any point after commencing ULT who had available follow-up, 12,060 (25.0%) had a repeat serum urate level recorded within 6 to 12 months of urate target attainment, with modest improvement after publication of the NICE guideline (Supplementary Figure S28A). Of those with a repeat urate measurement, 9,185 (76.2%) remained below 360 micromol/L (Supplementary Figure S28B).

### Flare management

Of 97,050 non-index gout flares recorded during the study period, 86,810 (89.4%) had a same-day prescription issued for a flare treatment, of which 53,975 (62.2%) were for colchicine, 22,645 (26.1%) were for NSAIDs, and 10,190 (11.7%) were for corticosteroids. The proportion of flares treated with colchicine increased over the study period, while NSAID usage declined and corticosteroid usage remained stable (Supplementary Figure S29).

### Sensitivity analyses

Results for the primary ITSA models were consistent across alternative Newey-West lag specifications and following adjustment for seasonality (Supplementary Table S2). Estimates for baseline serum urate testing and urate monitoring after ULT initiation were attenuated after accounting for reductions in testing during the pandemic (Supplementary Table S2).

ITSA models with alternative intervention dates were used to assess the sensitivity of findings to intervention timing. Moving the intervention date to June 2023, to allow for delayed guideline implementation, produced a similar magnitude but non-significant slowing in ULT initiation compared with the primary model (Supplementary Figure S30A). Moving the intervention date to June 2021, as a negative control, produced a smaller, non-significant change, suggesting that the change in trend in ULT initiation became more apparent around the time of guideline publication (Supplementary Figure S30B).

For urate target attainment within 12 months of ULT initiation, moving the intervention date to June 2023 produced a larger post-intervention increase than in the primary model, consistent with improvement continuing over time (Supplementary Figure S31A). Moving the intervention date to June 2021 produced a similar increase to the primary model, suggesting that the improving trend in urate target attainment was already underway before guideline publication and could not be clearly attributed to the NICE guideline (Supplementary Figure S31B).

## Discussion

In this nationwide study from England, we showed that routinely collected health data in OpenSAFELY can be used to monitor the implementation of a clinical guideline at scale. Despite improvements in some aspects of gout care, including greater use of low starting doses when initiating ULT, substantial gaps in management remain evident. By June 2025, only 43% of individuals with gout were prescribed ULT within 12 months of diagnosis, and only 33% of those initiating ULT achieved target serum urate levels. There was little evidence of guideline-specific effects on these trends, highlighting the need for targeted strategies to translate evidence-based recommendations into routine care.

Earlier studies from the UK and several other countries have highlighted suboptimal gout management.^7–9,22–25^ We previously showed that gout management guidelines from the British Society for Rheumatology (BSR 2017) and European Alliance of Associations for Rheumatology (EULAR 2016) were not associated with significant improvements in ULT initiation or urate target attainment in the UK.^7^ However, these guidelines were developed by specialist societies, whereas most patients with gout in the UK are managed in primary care. Publication of the NICE guideline in 2022 therefore represented an important opportunity to improve gout care, given the central role of NICE guidance in setting standards for clinical practice across the NHS.^2^

As with many long-term conditions, gout lacks a national quality improvement programme in the UK through which guideline implementation can be monitored. In this study, we demonstrated that routinely collected health data can help address this gap. ULT initiation increased gradually over the study period but remained below 50% in 2025. Rather than accelerating following publication of the NICE guideline, the rate of improvement in ULT prescribing slowed. This should not, however, be interpreted as evidence that the guideline adversely affected gout care. It is plausible that these changes would have occurred irrespective of guideline publication. Indeed, the post-intervention period coincided with recovery from the COVID-19 pandemic and sustained pressure on healthcare services, which are likely to have impacted on the management of long-term conditions such as gout.

Although improvements in serum urate target attainment were observed following publication of the NICE guideline, the magnitude of increase was relatively small, equivalent to approximately 3.8 additional people per 100 achieving target over three years. Sensitivity analyses suggested that the improving trend had begun before guideline publication, but despite this, target attainment remained low in June 2025, at only 37%. Achieving urate targets is central to preventing gout flares and complications associated with gout,^5,6,26^ and is possible for the vast majority of patients when a treat-to-target approach is followed.^26^ Our findings suggest that treat-to-target therapy is still not being implemented for the majority of gout patients in the UK.

We were also able to identify variation in gout care across population groups. Females were less likely than males to be started on ULT but more likely to achieve serum urate targets once treated, while individuals from more deprived areas were less likely to achieve target than those from less deprived areas. Younger adults were more likely than older adults to be initiated on ULT but substantially less likely to achieve target, which may in part reflect their lower likelihood of remaining prescribed ULT at 12 months. Similar age-related differences in treatment persistence have been reported for type 2 diabetes in UK primary care.^27^ Identifying these differences using routinely collected data may help target interventions to groups with the greatest unmet need. For younger adults, this could include support to maintain long-term ULT use and reduce the risk of recurrent gout flares and longer-term complications of inadequately treated gout, including progressive joint damage, CKD and cardiovascular disease.^5,6,26^

We showed that ULT prescribing remains dominated by allopurinol, despite NICE recommending either drug as first-line treatment.^2^ This recommendation differs from BSR and other gout guidelines, and was informed by updated economic analyses reflecting the reduced cost of febuxostat.^28^ We found that persistence at 12 months was lower among patients initiating febuxostat than allopurinol, although the small number of patients receiving febuxostat warrants caution. There may also have been clinical reasons for the selection of febuxostat, such as previous use of allopurinol in secondary care that were not captured in our data.

An aspect of gout care that improved considerably over the study period was prescribing safety. There were large reductions in the proportion of patients starting ULT at higher doses, which increases the risk of adverse cutaneous reactions. Febuxostat prescribing in patients with major cardiovascular disease also declined from 2019 onwards, coinciding with warnings from the Medicines and Healthcare products Regulatory Agency (MHRA) advising against its use in patients with pre-existing major cardiovascular disease unless no suitable alternatives were available.^29^ Following publication of the FAST trial in 2020,^30^ the MHRA softened this advice in May 2023 to recommend cautious use in this population.^31^ Despite this revision, we found that febuxostat prescribing in people with cardiovascular disease continued to decline, suggesting a sustained influence of the initial safety warning.

There are several potential explanations for why publication of a NICE guideline has not yet been associated with substantial improvements in gout care beyond existing trends. The guideline was published during a period of health service recovery following the COVID-19 pandemic, and the post-intervention period in this study may have been too short to capture the full effects of implementation. However, the lack of substantial improvement beyond existing trends is consistent with previous studies highlighting challenges in improving gout care.^7–9,22–25^ Implementing treat-to-target ULT requires repeated blood tests and coordinated dose titration, which need to be delivered alongside competing and increasing demands in primary care.^32^ Unlike long-term conditions such as diabetes, gout is absent from the Quality and Outcomes Framework in the UK, providing little financial incentive to systematically identify patients who remain undertreated. Previous studies have also highlighted multiple barriers to effective gout management, including stigma around the diagnosis and an excessive focus on dietary modification.^33^

Implementation strategies are therefore essential to improving gout care. Education should promote a shared understanding of gout as a long-term condition requiring sustained urate-lowering treatment. Nurse-led treat-to-target care has been shown to be highly effective in trial settings,^26^ although the intensity of follow-up presents challenges for delivery within stretched healthcare services. Digital approaches to supporting implementation are particularly attractive in UK primary care, where electronic health records are used almost universally. Automated recall systems for serum urate testing, clinical decision support templates and prescribing prompts, and dashboards identifying patients who have not commenced ULT or remain above target could support delivery of treat-to-target care while reducing the need for repeated manual review.

Importantly, our study demonstrates how routinely collected data can support near real-time monitoring of guideline implementation and care quality. Once quality metrics have been defined and computed, they can be updated regularly without repeated manual data collection.^34^ For organisations such as NICE, this approach could enable more routine monitoring of guideline implementation, helping to identify gaps or variation in care and inform where further action may be required. This, in turn, allows more time to be directed towards improving care rather than measuring it, while data-enabled trials could be used to evaluate quality improvement interventions in real-world practice.^35^ Our openly shared, adaptable code provides a basis for applying this approach across many other diseases and guidelines.

Several limitations should also be considered. Some aspects of gout management, such as patient education, are challenging to ascertain from electronic health records. We therefore focused on outcomes that could be captured more consistently, including ULT initiation and urate target attainment; however, we could not determine whether ULT was offered to patients but declined, and prescribing records do not confirm medication adherence. Prescriptions and laboratory tests recorded exclusively in secondary care were unavailable, although most individuals with gout in England are managed in primary care. Similarly, the use of non-prescription medications, such as over-the-counter ibuprofen would not be captured when assessing flare treatment. Some analyses were limited by small numbers, which precluded subgroup analyses by ethnicity. Interrupted time-series analysis cannot distinguish guideline effects from concurrent changes in healthcare delivery, including lasting effects of the COVID-19 pandemic. Some diagnostic misclassification is also likely in studies using coded health data, although previous studies have demonstrated good validity of gout coding in UK primary care data.^18,36^ Finally, as our study was conducted in a single health system, the findings may not be directly generalisable beyond the NHS in England.

In conclusion, despite improvements in some aspects of gout care, substantial gaps remain between clinical practice and NICE guideline recommendations, particularly for ULT initiation and serum urate target attainment. Guideline publication was not associated with marked improvements beyond existing temporal trends, highlighting the need for implementation strategies to improve gout care. Importantly, our approach provides an adaptable framework for monitoring quality of care and guideline implementation at scale using routinely collected data.

## Summary box

### What is already known on this topic

• Treat-to-target urate-lowering therapy can prevent recurrent gout flares and complications associated with gout, but previous studies have shown low treatment uptake and serum urate target attainment.

• NICE published its first gout management guideline in June 2022; however, whether its publication was associated with improvements in gout care was unknown, reflecting broader challenges in monitoring guideline implementation in routine care.

### What this study adds

• Among people newly diagnosed with gout in England, 36% started urate-lowering therapy within 12 months of diagnosis, while only 33% of those starting treatment had a recorded serum urate level below target within 12 months.

• Publication of NICE guidance was not associated with substantial improvements in these treatment metrics beyond existing trends.

• This study highlights the potential for routinely collected data in OpenSAFELY to monitor guideline uptake at scale.

## Data Availability

The General Practice data in the NHS England OpenSAFELY Analytics Service used for this study is currently drawn from General Practice practices in England using the SystmOne software (for which TPP data across England is the data processor company). Linked data from hospitals and death registrations are also available. Access to the underlying identifiable and potentially re-identifiable pseudonymised electronic health record data is tightly governed by various legislative and regulatory frameworks, and restricted by best practice. TPP developers initiate an automated process to create pseudonymised records in the core OpenSAFELY database, which are copies of key structured data tables in the identifiable records. These pseudonymised records are linked onto key external data resources that have also been pseudonymised via SHA-512 one-way hashing of NHS numbers using a shared salt. University of Oxford, Bennett Institute for Applied Data Science developers and PIs, who hold contracts with NHS England, have access to the OpenSAFELY pseudonymised data tables to develop the OpenSAFELY tools. These tools in turn enable researchers with OpenSAFELY data access agreements to write and execute code for data management and data analysis without direct access to the underlying raw pseudonymised patient data, and to review the outputs of this code. All OpenSAFELY platform software is shared for inspection on GitHub: https://github.com/OpenSAFELY-Core. All code for the full data management pipeline, from raw data to completed results, for this analysis are available for review and reuse for all OpenSAFELY analyses [https://github.com/opensafely/nice_gout].

## Acknowledgements

We are very grateful for all the support received from the TPP Technical Operations team throughout this work, and for generous assistance from the information governance and database teams at NHS England and the NHS England Transformation Directorate. We acknowledge members of the OpenSAFELY Collaborative, who have contributed to the development and maintenance of the OpenSAFELY platform.

## Contributorship and transparency statement

Contributions were as follows: Conceptualisation: MDR, JBG, SN, BM; Methodology: MDR, JBG, SN, BM, ER, AA, KC, MG; Data curation and software development: MDR, BG, BM, AS, AM, SB; Formal analysis: MDR, JBG; Interpretation of findings: all authors; Writing – original draft: MDR; Writing – revising, review and editing: all authors. All authors read and approved the final manuscript. MDR and JBG are the guarantors for the article; accept full responsibility for the work and/or the conduct of the study; and controlled the decision to publish. MDR and JBG directly accessed and verified the underlying data reported in the manuscript. The corresponding author attests that all listed authors meet authorship criteria and that no others meeting the criteria have been omitted. The lead author affirms that the manuscript is an honest, accurate, and transparent account of the study being reported; that no important aspects of the study have been omitted; and that any discrepancies from the study as originally planned have been explained.

## Data access and verification

The General Practice data in the NHS England OpenSAFELY Analytics Service used for this study is currently drawn from General Practice practices in England using the SystmOne software (for which TPP data across England is the data processor company). Linked data from hospitals and death registrations are also available. Access to the underlying identifiable and potentially re-identifiable pseudonymised electronic health record data is tightly governed by various legislative and regulatory frameworks, and restricted by best practice. TPP developers initiate an automated process to create pseudonymised records in the core OpenSAFELY database, which are copies of key structured data tables in the identifiable records. These pseudonymised records are linked onto key external data resources that have also been pseudonymised via SHA-512 one-way hashing of NHS numbers using a shared salt. University of Oxford, Bennett Institute for Applied Data Science developers and PIs, who hold contracts with NHS England, have access to the OpenSAFELY pseudonymised data tables to develop the OpenSAFELY tools. These tools in turn enable researchers with OpenSAFELY data access agreements to write and execute code for data management and data analysis without direct access to the underlying raw pseudonymised patient data, and to review the outputs of this code. All OpenSAFELY platform software is shared for inspection on GitHub: https://github.com/OpenSAFELY-Core. All code for the full data management pipeline, from raw data to completed results, for this analysis are available for review and reuse for all OpenSAFELY analyses [https://github.com/opensafely/nice_gout]. The data management and analysis code for this paper was led by MDR and contributed to by JBG.

## Information governance

NHS England is the data controller of the NHS OpenSAFELY Analytics Service; TPP is the data processor; all study authors using OpenSAFELY have the approval of NHS England.^37,38^ TPP is accredited to the ISO 27001 information security standard and is NHS IG Data Security and Information Governance Toolkit compliant.^39^ Patient data has been pseudonymised for analysis and linkage using industry standard cryptographic hashing techniques; all pseudonymised datasets transmitted for linkage onto OpenSAFELY are encrypted; access to the NHS OpenSAFELY Analytics Service is via a virtual private network (VPN) connection; the researchers hold contracts with NHS England and only access the platform to initiate database queries and statistical models; all database activity is logged; only aggregate statistical outputs leave the platform environment following best practice for anonymisation of results such as statistical disclosure control for low cell counts.^40^ The service adheres to the obligations of the UK General Data Protection Regulation (UK GDPR) and the Data Protection Act 2018. The Secretary of State has requested that NHS England operates the Service under the NHS OpenSAFELY Data Analytics Service Pilot Directions 2025 which came into force on 17 June 2025.^14^ Taken together, these provide the legal bases to link patient datasets using the service.

