## Supplementary Material for "Trends in gout management before and after publication of NICE guidance in England: a population-based study in OpenSAFELY"

**Supplementary Figure S1.** Temporal trends in the proportion of adults with new gout diagnoses who had serum urate levels recorded at diagnosis. In the top panel (A), individual data points represent the mean proportion of new gout diagnoses who had serum urate levels recorded within 2 months before or after diagnosis, with trend lines representing 3-monthly rolling averages of the current, preceding and subsequent months. The bottom panel (B) shows an interrupted time-series comparing trends before and after the publication of the NICE Gout Management Guideline in June 2022. Trend lines were fitted before and after publication of the NICE guideline, with annualised coefficients presented as percentage-point change per year, alongside the change in trends (after vs. before publication) and the immediate step change at the date of publication.

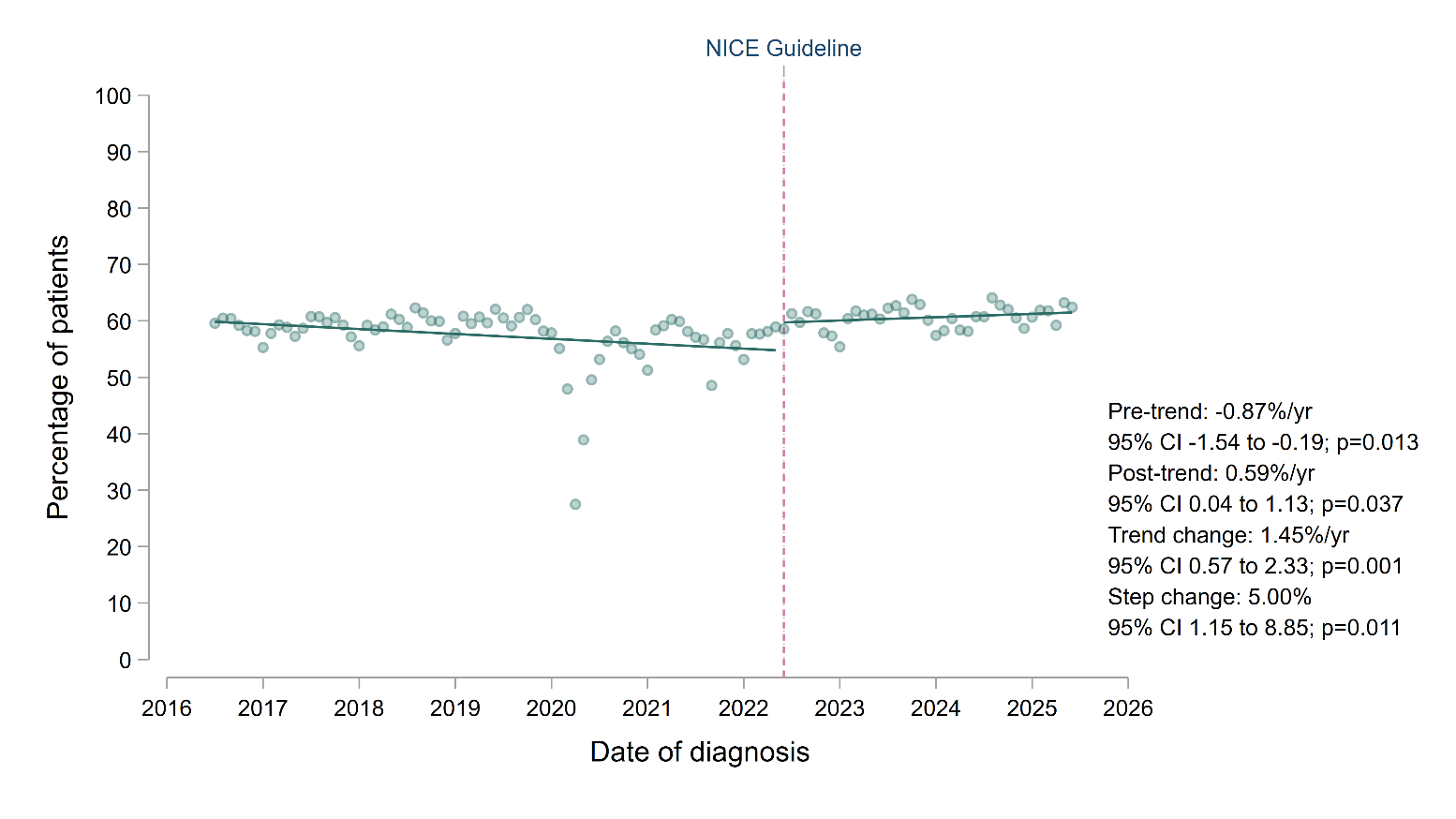

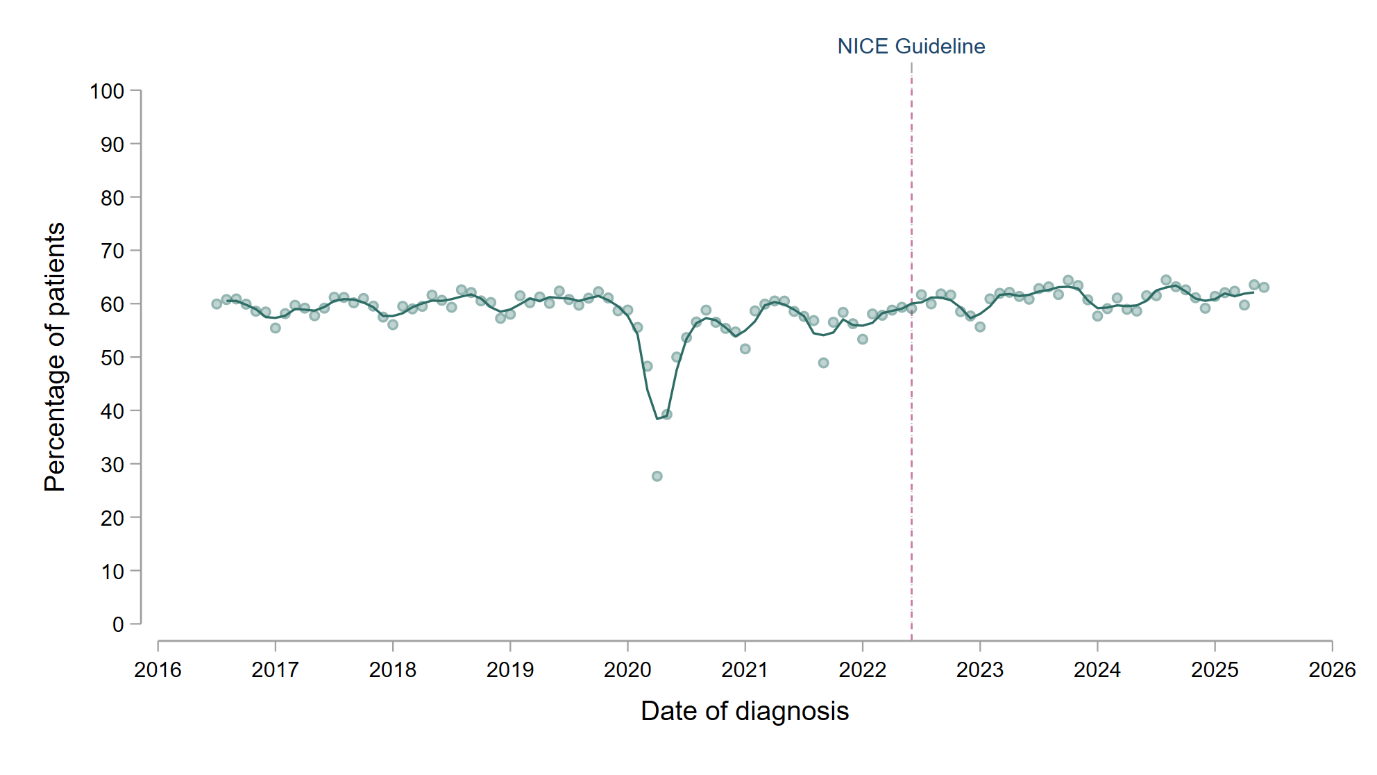

A

B

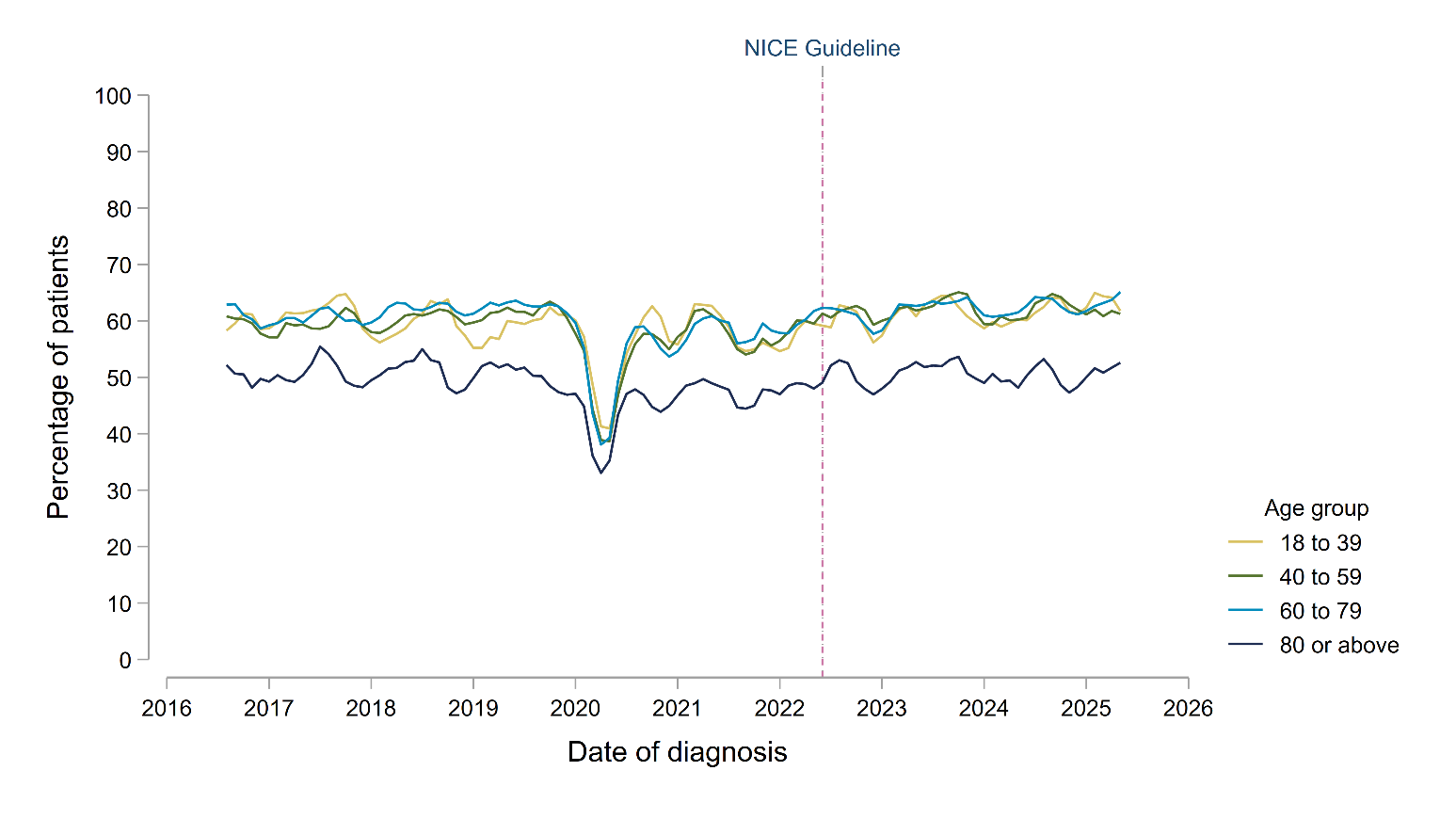
**Supplementary Figure S2.** Temporal trends in the proportion of adults with new gout diagnoses who had serum urate levels recorded within 2 months before or after diagnosis, separated by age group. Individual trend lines represent 3-monthly rolling averages of the current, preceding and subsequent months. The vertical dashed line represents the publication of the NICE Gout Management Guideline in June 2022.

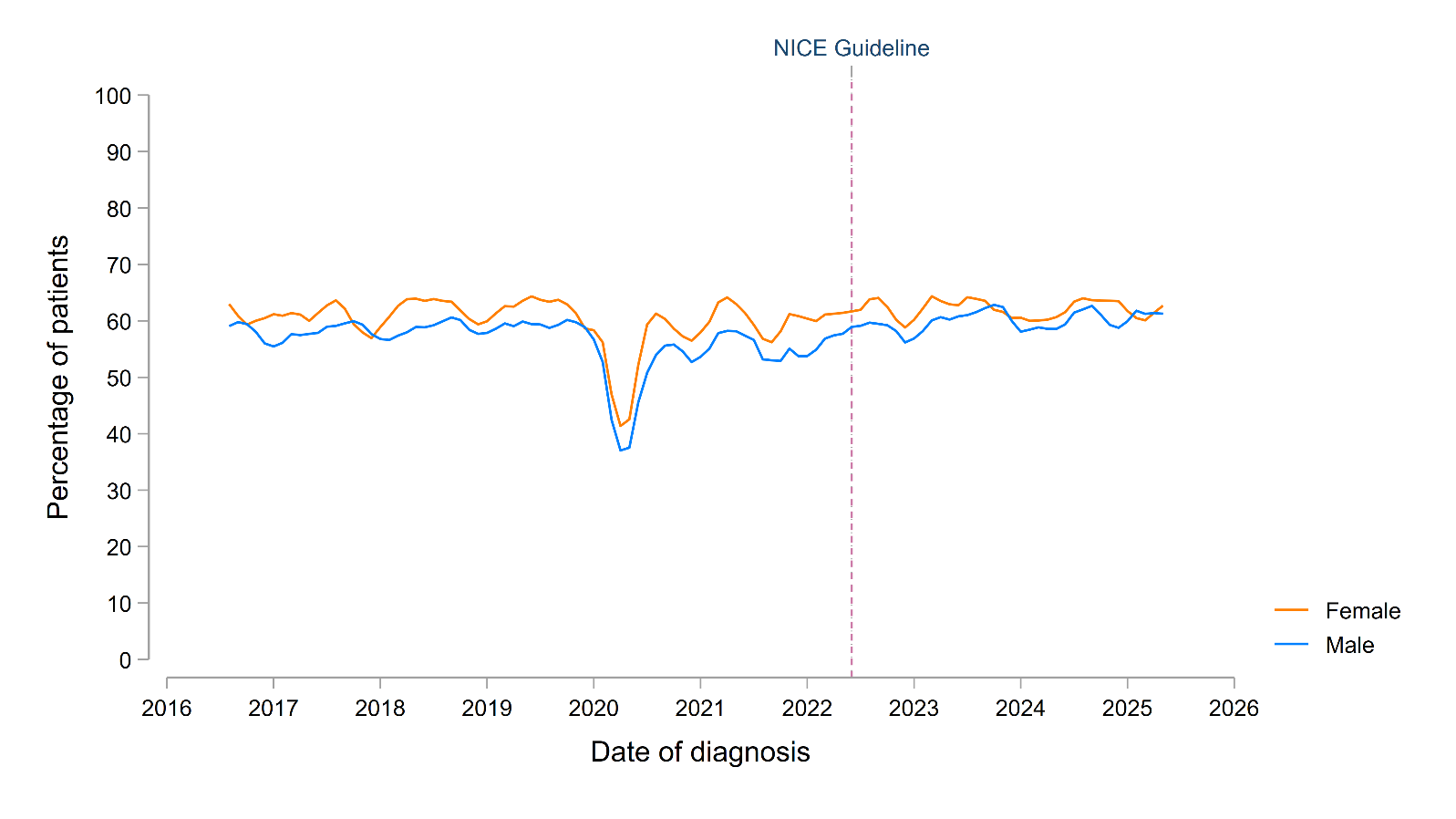
**Supplementary Figure S3.** Temporal trends in the proportion of adults with new gout diagnoses who had serum urate levels recorded within 2 months before or after diagnosis, separated by sex. Individual trend lines represent 3-monthly rolling averages of the current, preceding and subsequent months. The vertical dashed line represents the publication of the NICE Gout Management Guideline in June 2022.

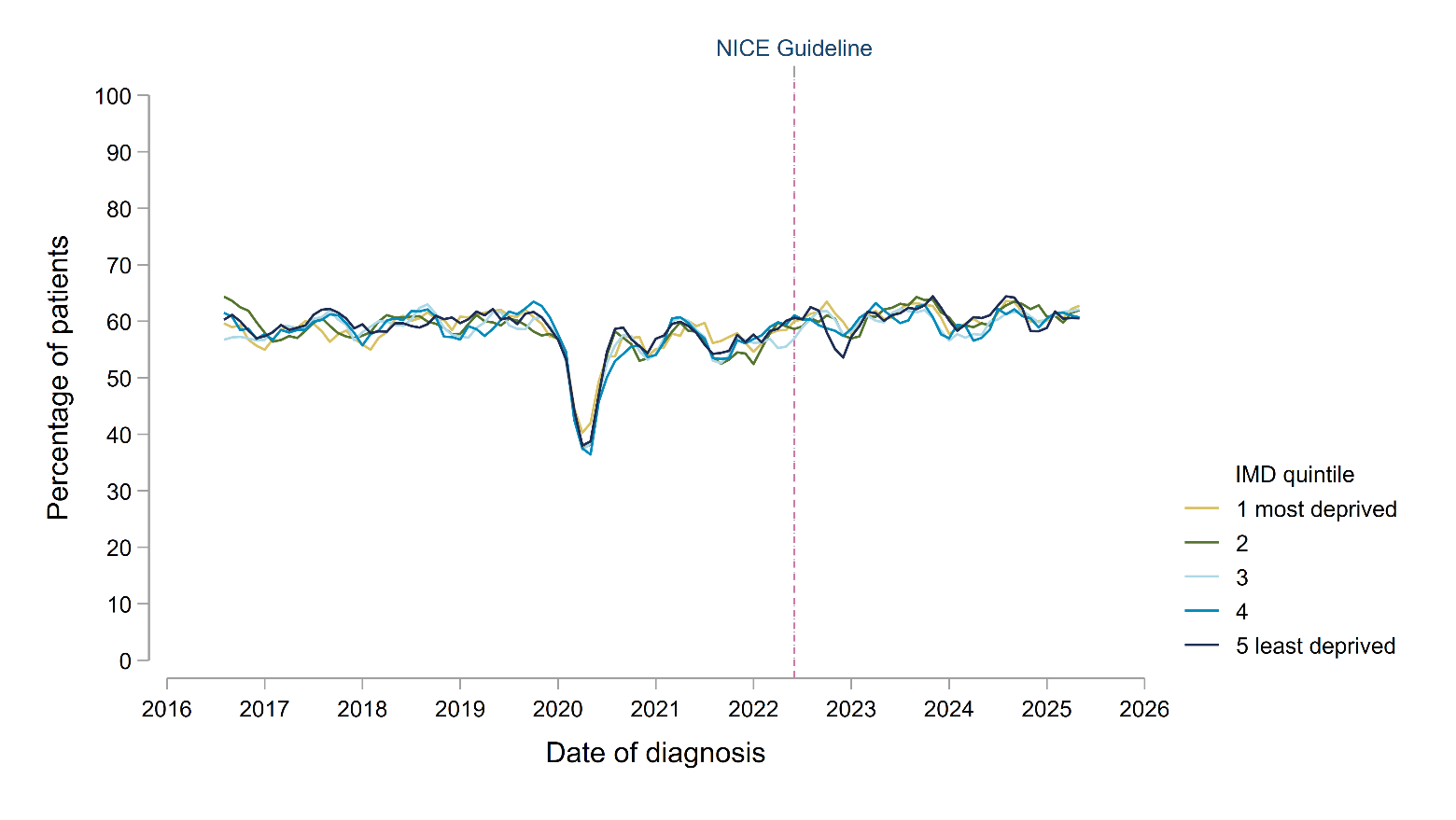
**Supplementary Figure S4.** Temporal trends in the proportion of adults with new gout diagnoses who had serum urate levels recorded within 2 months before or after diagnosis, separated by socioeconomic status (defined by quintiles of index of multiple deprivation [IMD]). Individual trend lines represent 3-monthly rolling averages of the current, preceding and subsequent months. The vertical dashed line represents the publication of the NICE Gout Management Guideline in June 2022.

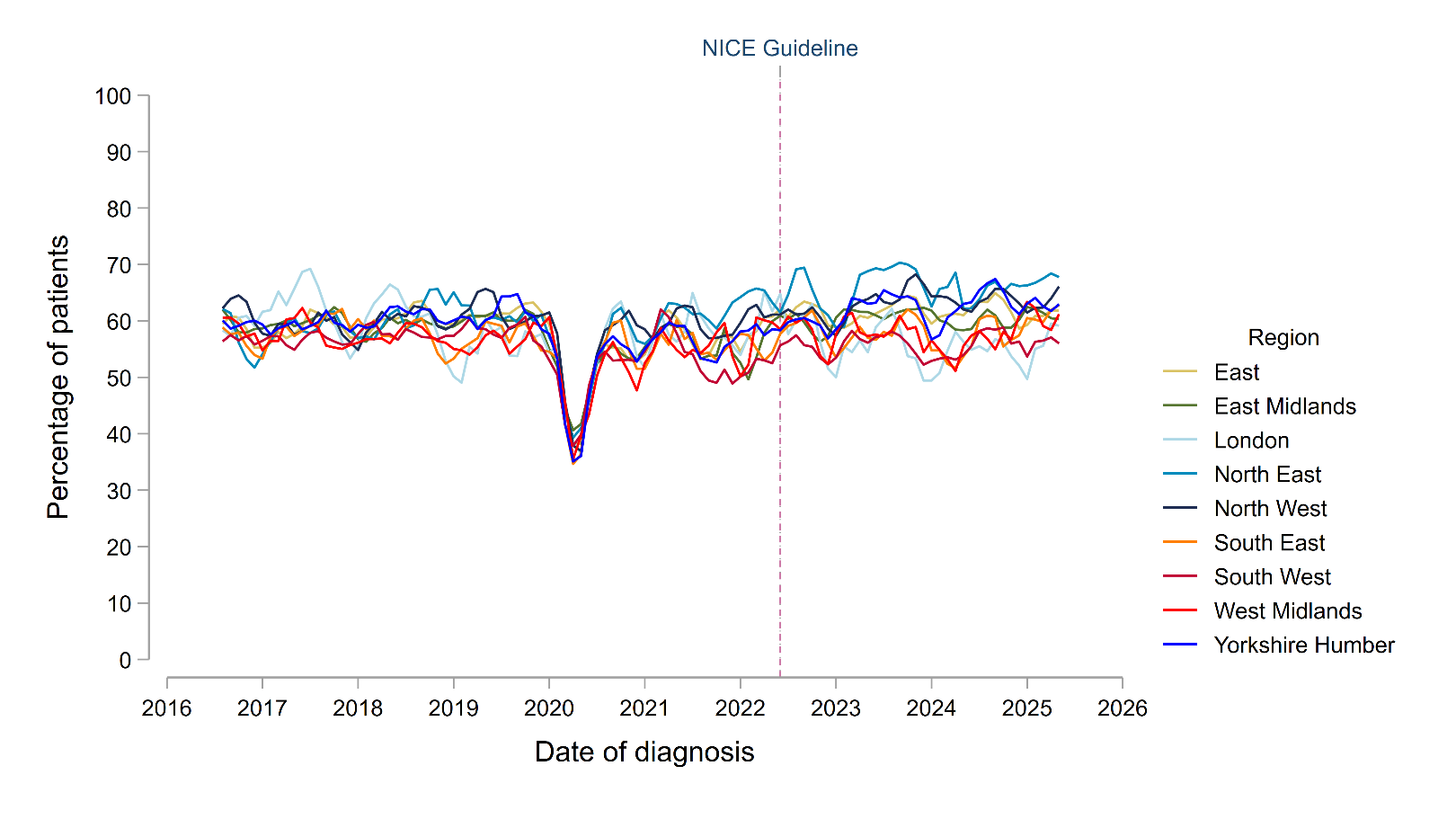
**Supplementary Figure S5.** Temporal trends in the proportion of adults with new gout diagnoses who had serum urate levels recorded within 2 months before or after diagnosis, separated by region of England. Individual trend lines represent 3-monthly rolling averages of the current, preceding and subsequent months. The vertical dashed line represents the publication of the NICE Gout Management Guideline in June 2022.

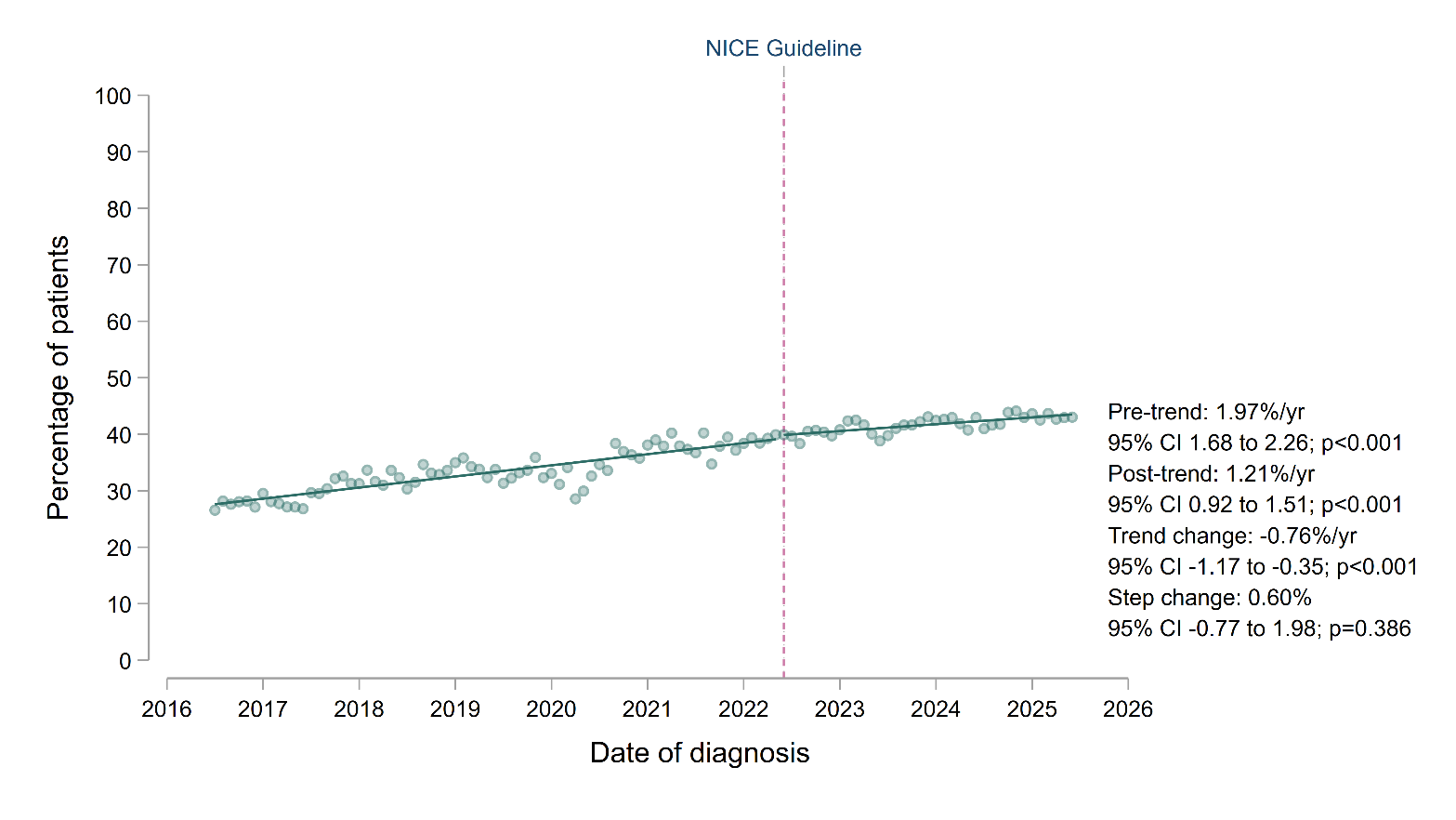
**Supplementary Figure S6.** Interrupted time-series comparing trends in the initiation of urate-lowering therapy (ULT) within 12 months of gout diagnosis before and after the publication of the NICE Gout Management Guideline in June 2022. Individual data points represent the mean proportion who were prescribed ULT within 12 months of diagnosis. Trend lines were fitted before and after publication of the NICE guideline, with annualised coefficients presented as percentage-point change per year, alongside the change in trends and the immediate step change at the date of publication.

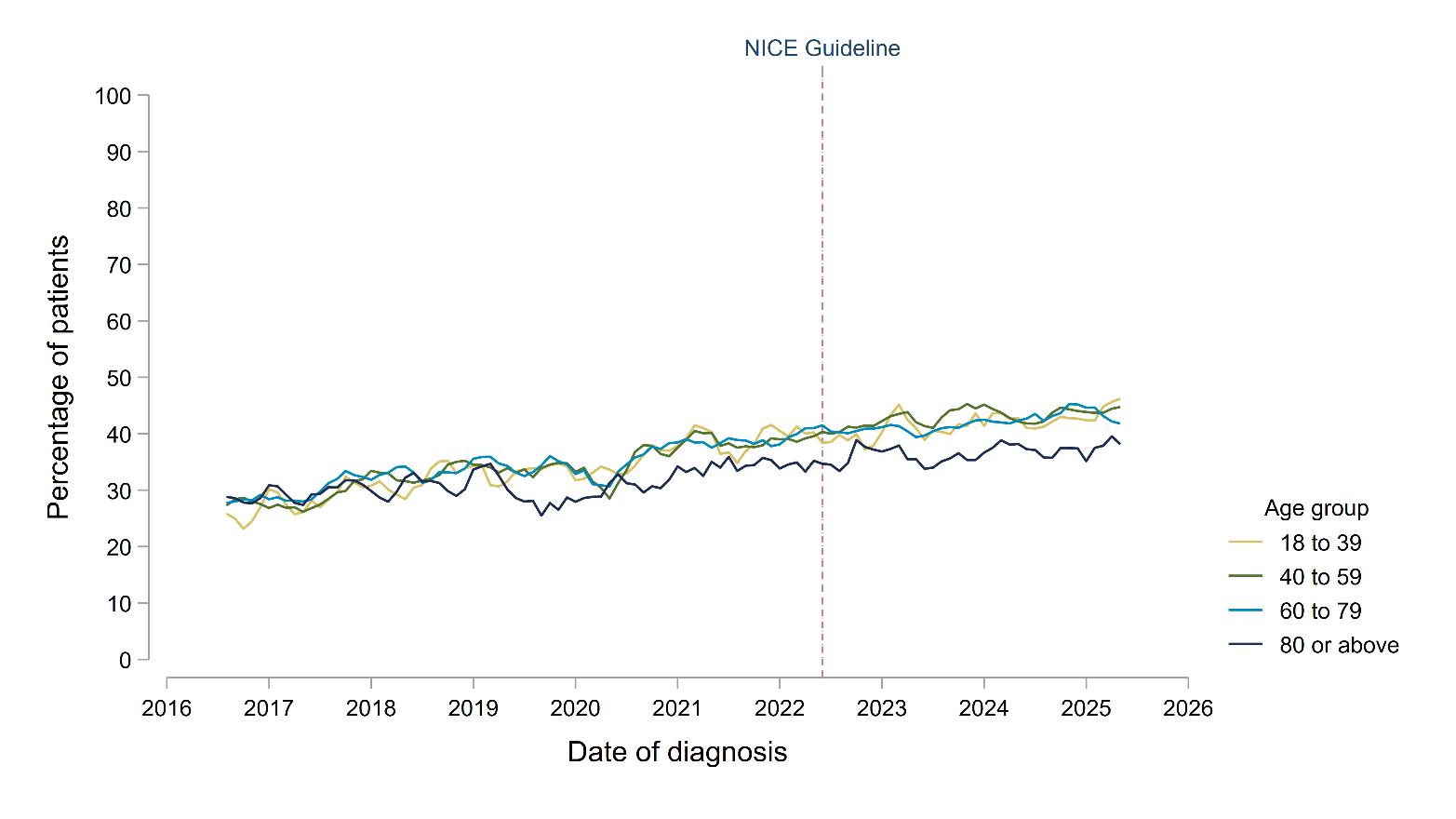
**Supplementary Figure S7.** Temporal trends in the initiation of urate-lowering therapy (ULT) within 12 months of diagnosis for all adults with new gout diagnoses, separated by age group. Individual trend lines represent 3-monthly rolling averages of the current, preceding and subsequent months. The vertical dashed line represents the publication of the NICE Gout Management Guideline in June 2022.

**Supplementary Figure S8.** Temporal trends in the initiation of urate-lowering therapy (ULT), separated by sex. The top panel (A) reports ULT initiation within 12 months of diagnosis for all adults with new gout diagnoses; the bottom panel (B) reports ULT initiation within 12 months of the onset of risk factors indicating a need to be offered ULT in line with NICE guidance (recurrent gout flares, CKD stages 3 to 5, diuretic use, chronic gouty arthritis or tophaceous gout). Individual trend lines represent 3-monthly rolling averages of the current, preceding and subsequent months. The vertical dashed line represents the publication of the NICE Gout Management Guideline in June 2022.

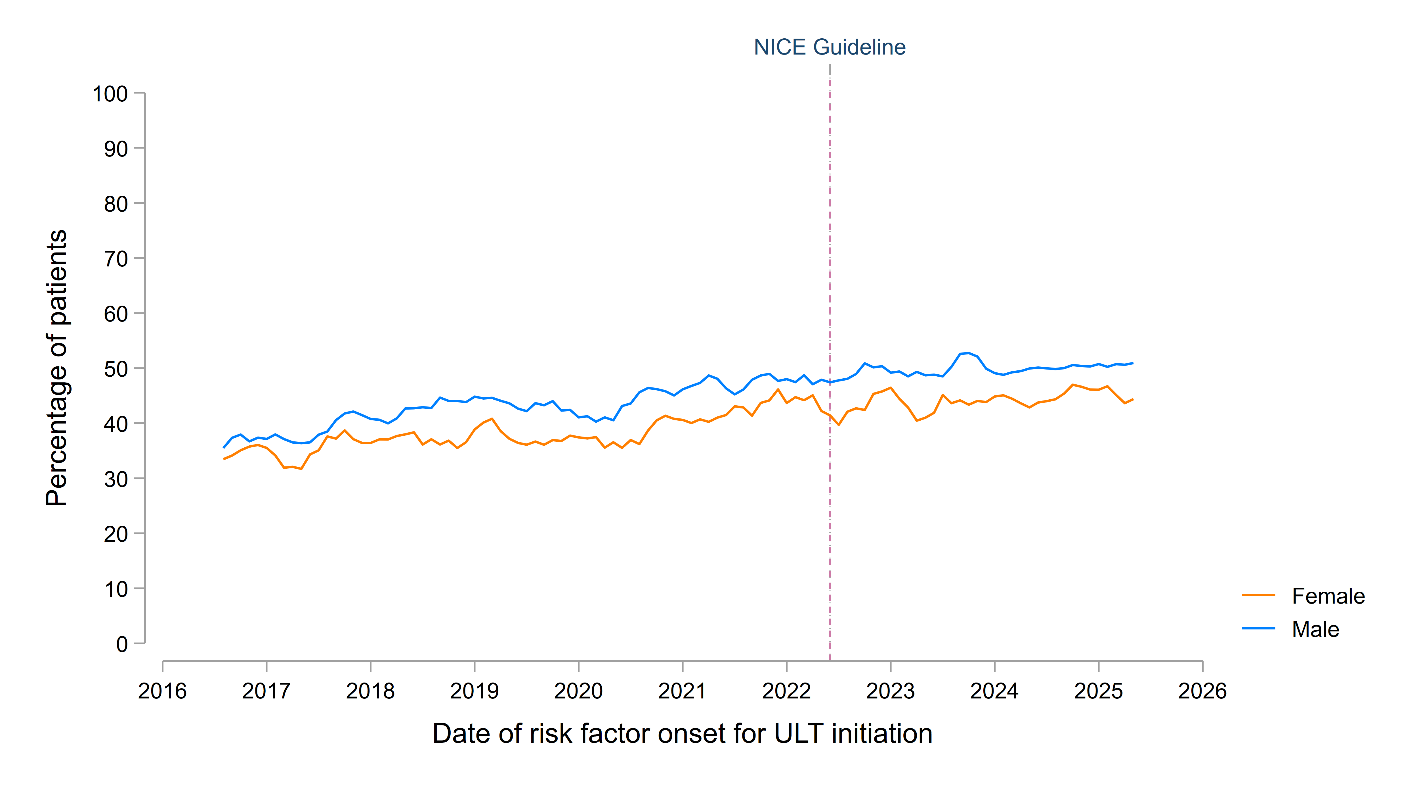

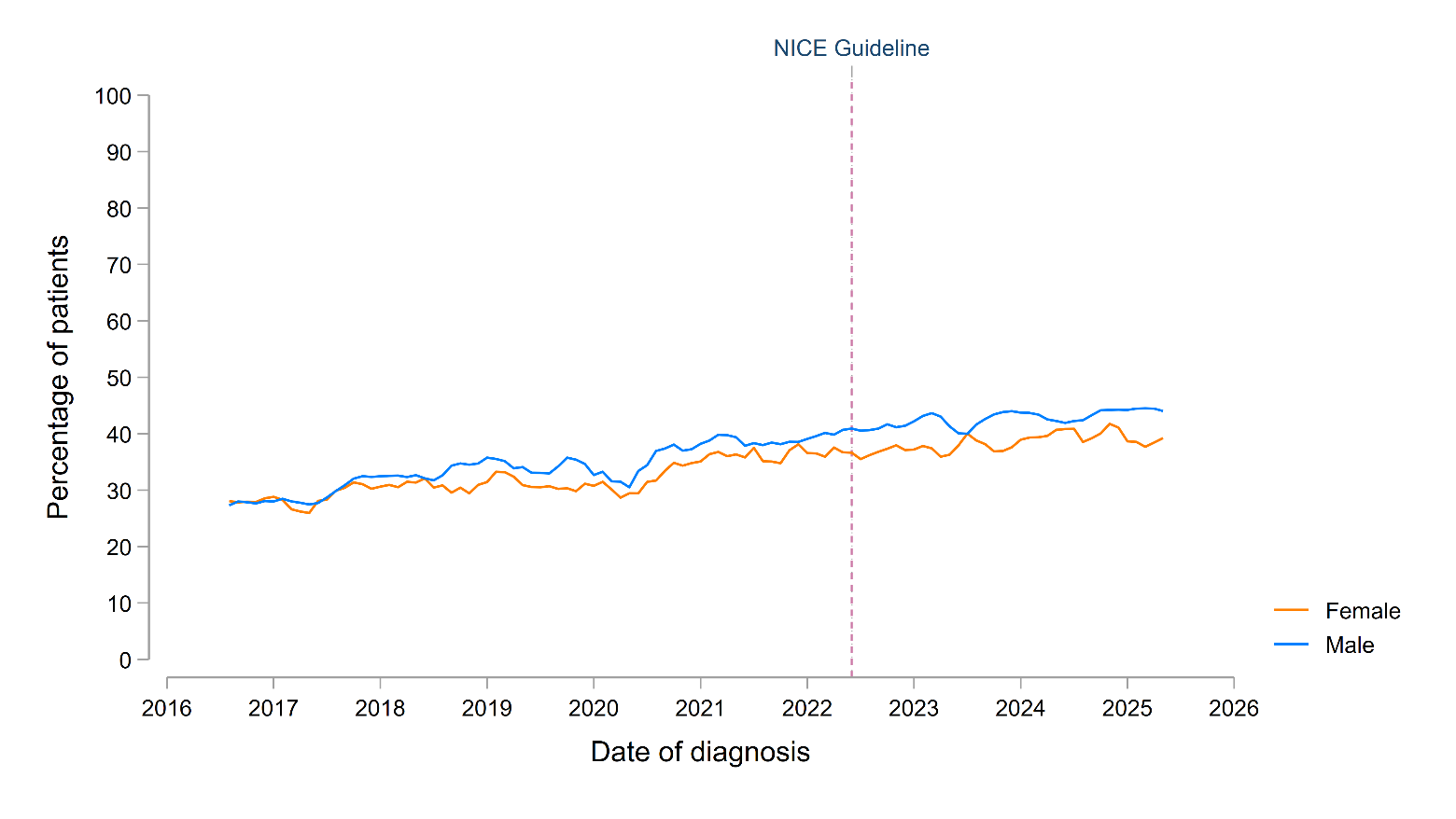

B

A

**Supplementary Figure S9.** Temporal trends in the initiation of urate-lowering therapy (ULT), separated by socioeconomic status (defined by quintiles of index of multiple deprivation [IMD]). The top panel (A) reports ULT initiation within 12 months of diagnosis for all adults with new gout diagnoses; the bottom panel (B) reports ULT initiation within 12 months of the onset of risk factors indicating a need to be offered ULT in line with NICE guidance (recurrent gout flares, CKD stages 3 to 5, diuretic use, chronic gouty arthritis or tophaceous gout). Individual trend lines represent 3-monthly rolling averages of the current, preceding and subsequent months. The vertical dashed line represents the publication of the NICE Gout Management Guideline in June 2022.

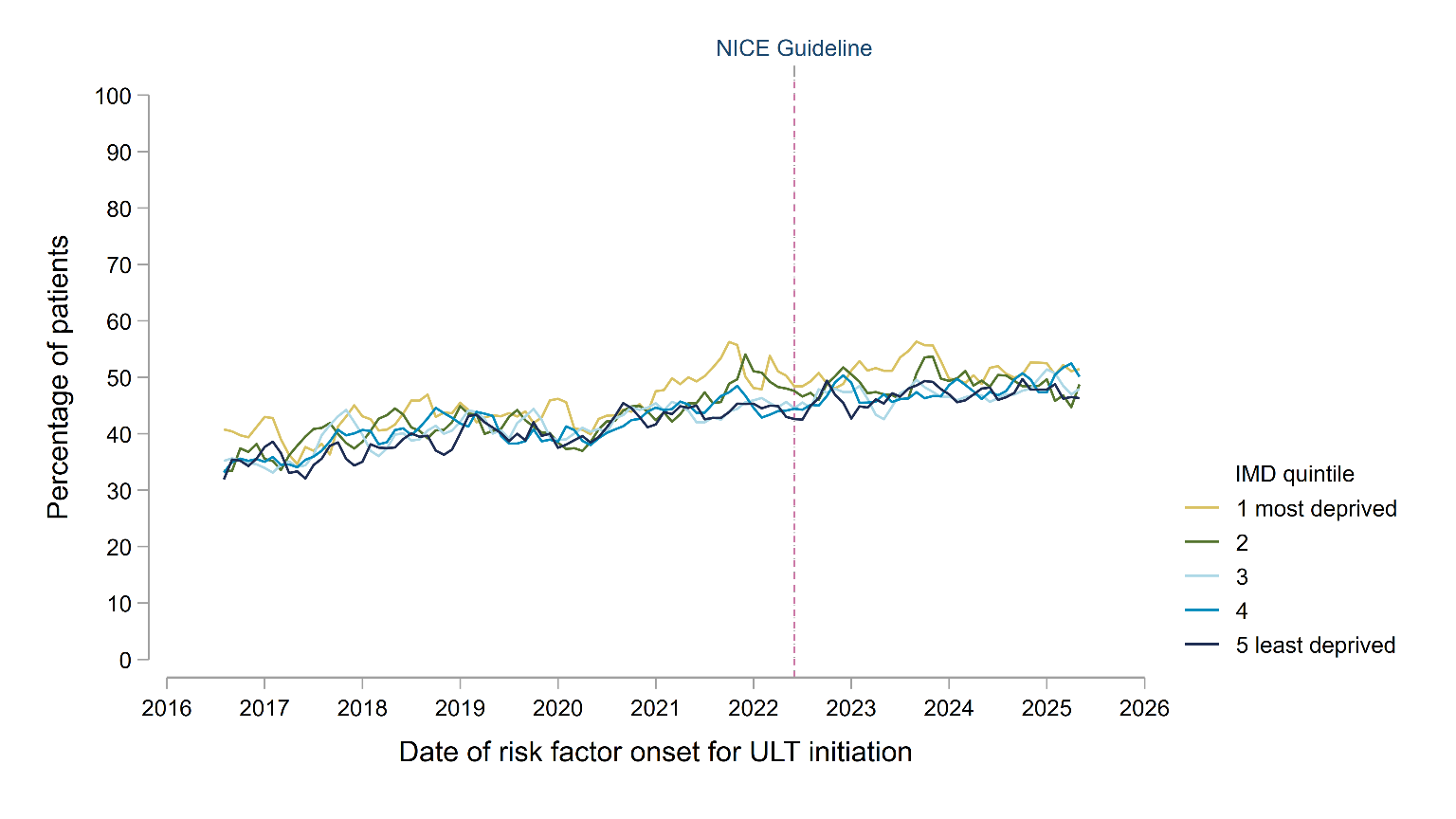

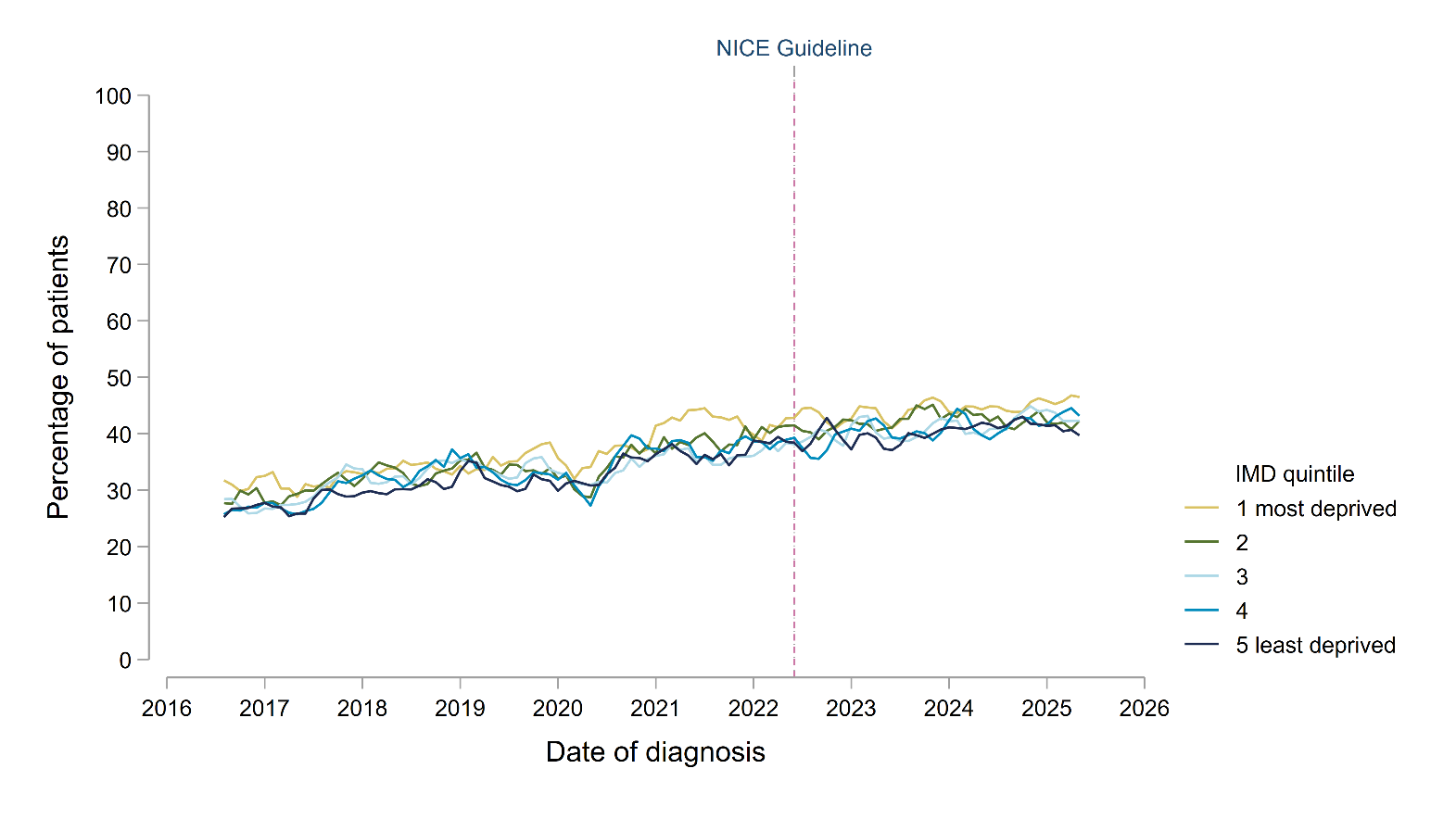

A

B

**Supplementary Figure S10.** Temporal trends in the initiation of urate-lowering therapy (ULT), separated by region. The top panel (A) reports ULT initiation within 12 months of diagnosis for all adults with new gout diagnoses; the bottom panel (B) reports ULT initiation within 12 months of the onset of risk factors indicating a need to be offered ULT in line with NICE guidance (recurrent gout flares, CKD stages 3 to 5, diuretic use, chronic gouty arthritis or tophaceous gout). Individual trend lines represent 3-monthly rolling averages of the current, preceding and subsequent months. The vertical dashed line represents the publication of the NICE Gout Management Guideline in June 2022.

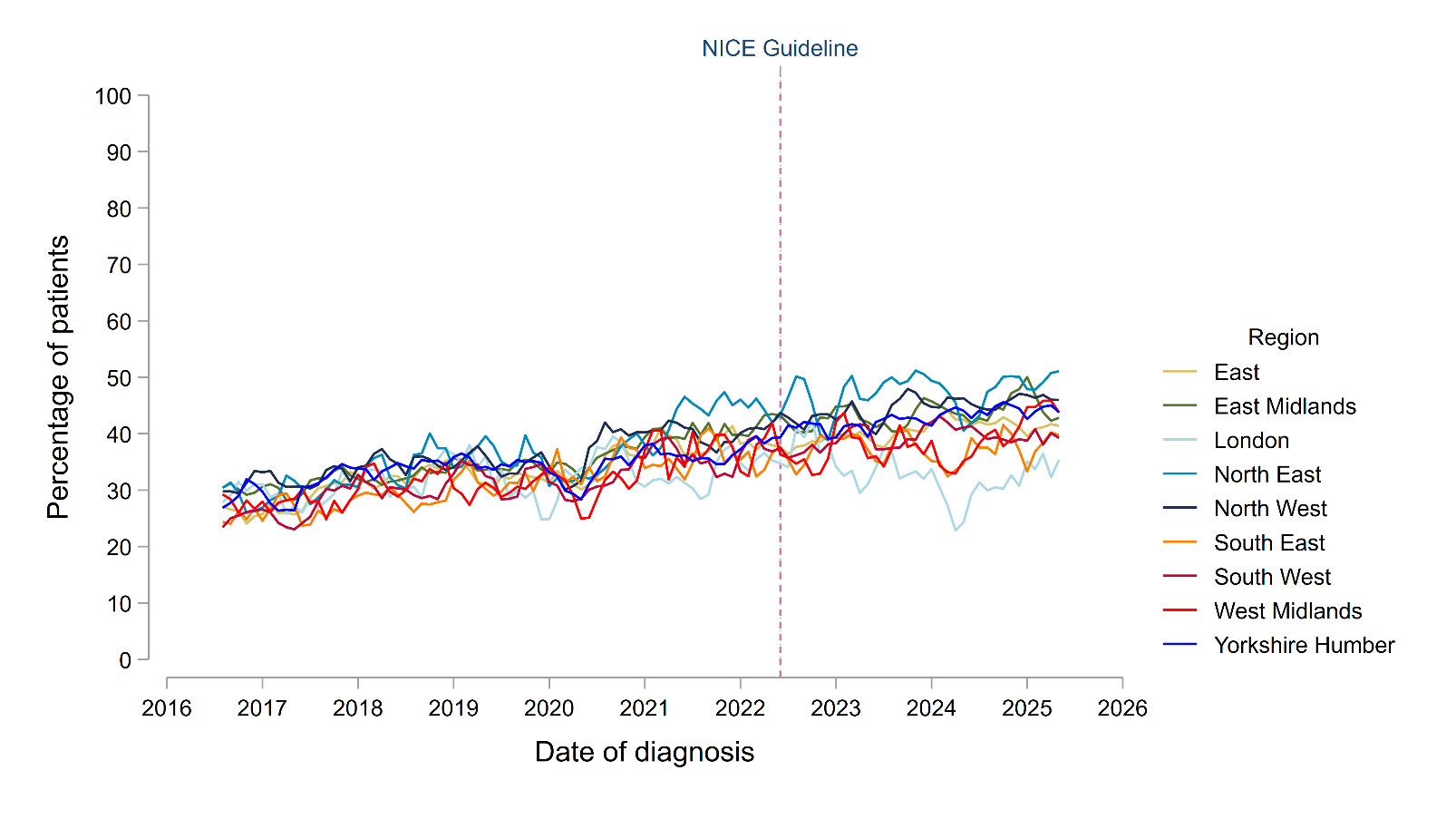

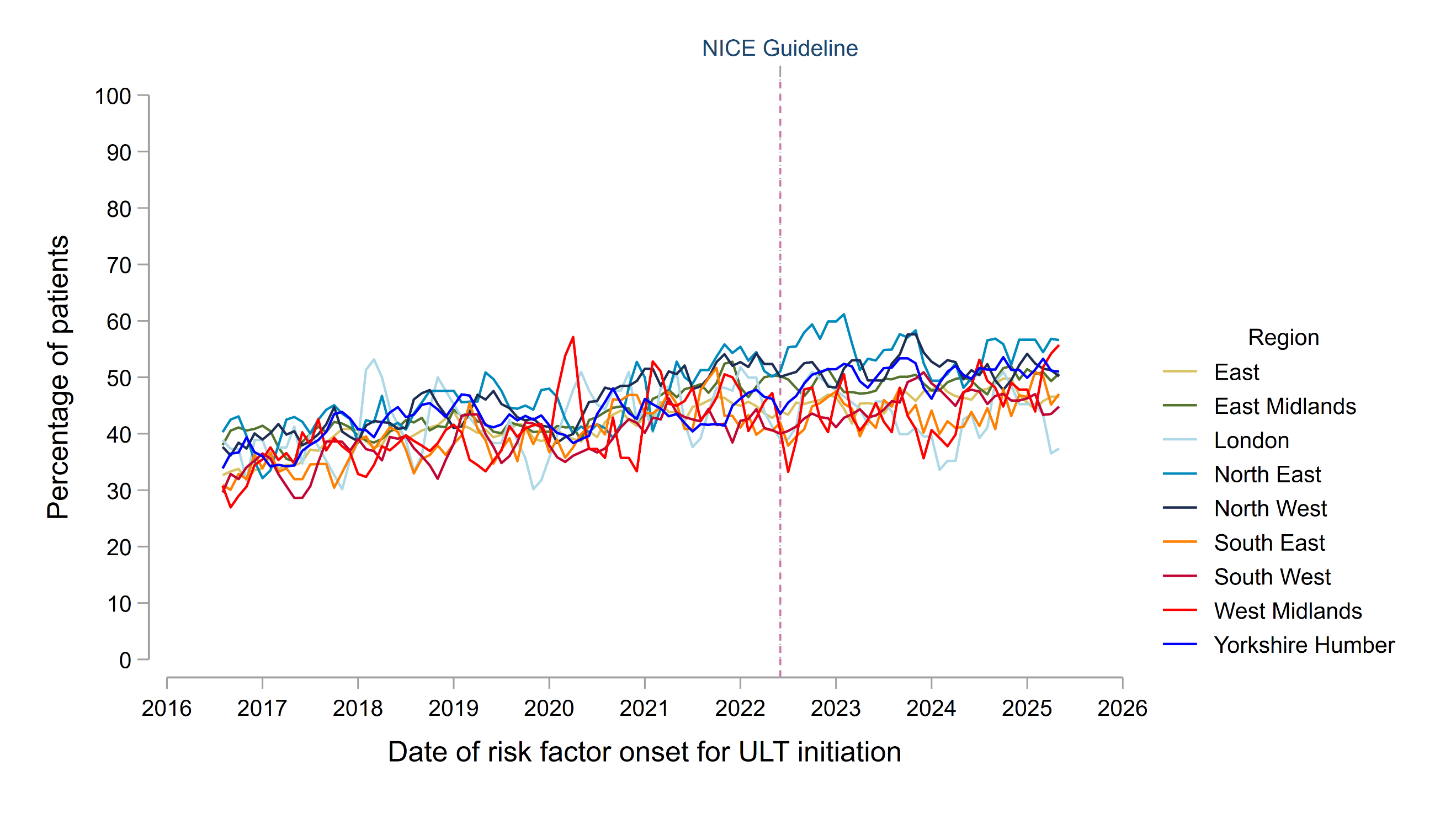

A

B

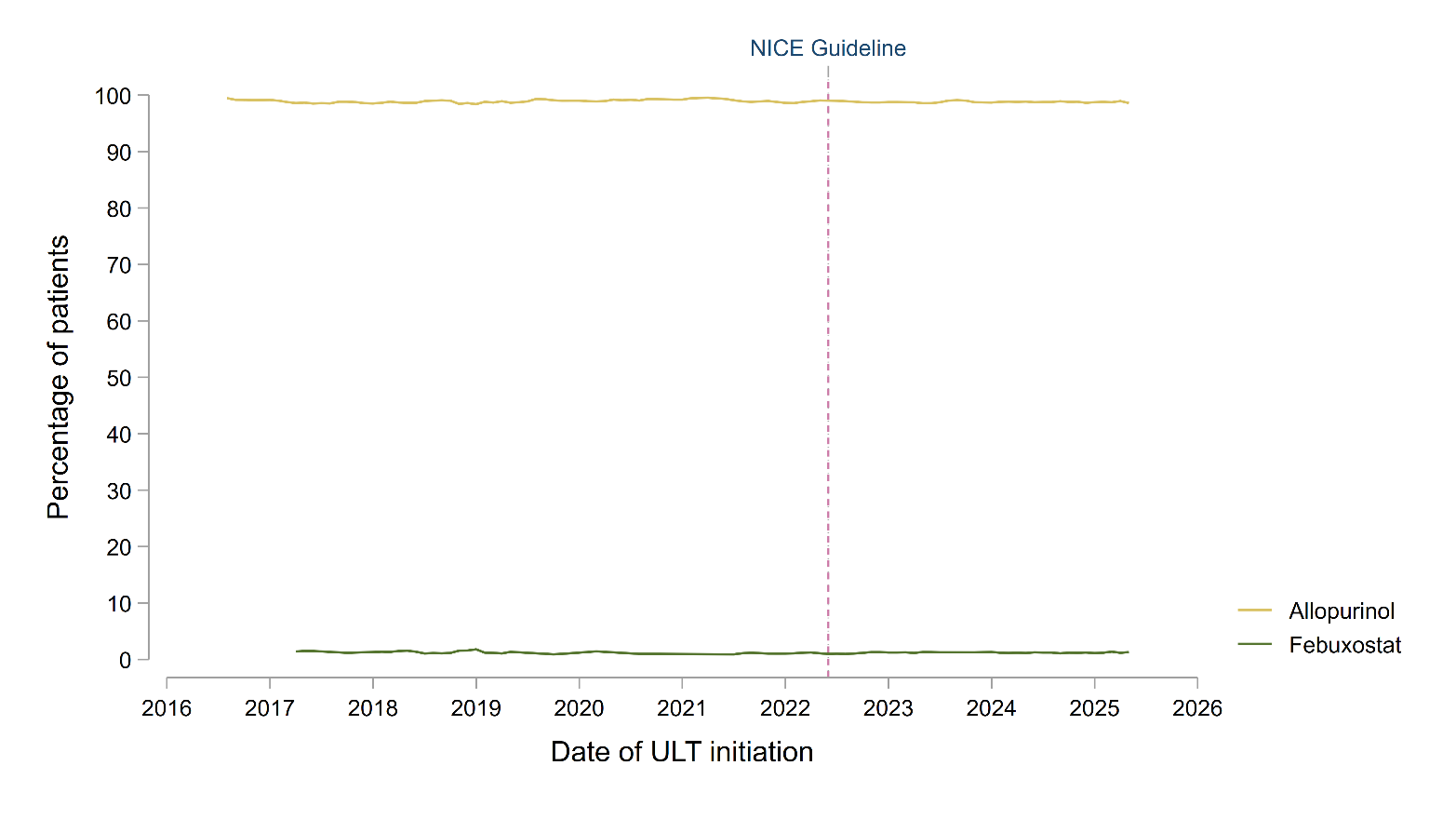
**Supplementary Figure S11.** Temporal trends in the choice of first urate-lowering therapy (ULT) medication in individuals diagnosed with gout during the study period. Individual trend lines represent 3-monthly rolling averages of the current, preceding and subsequent months. The vertical dashed line represents the publication of the NICE Gout Management Guideline in June 2022. Small number suppression has been applied.

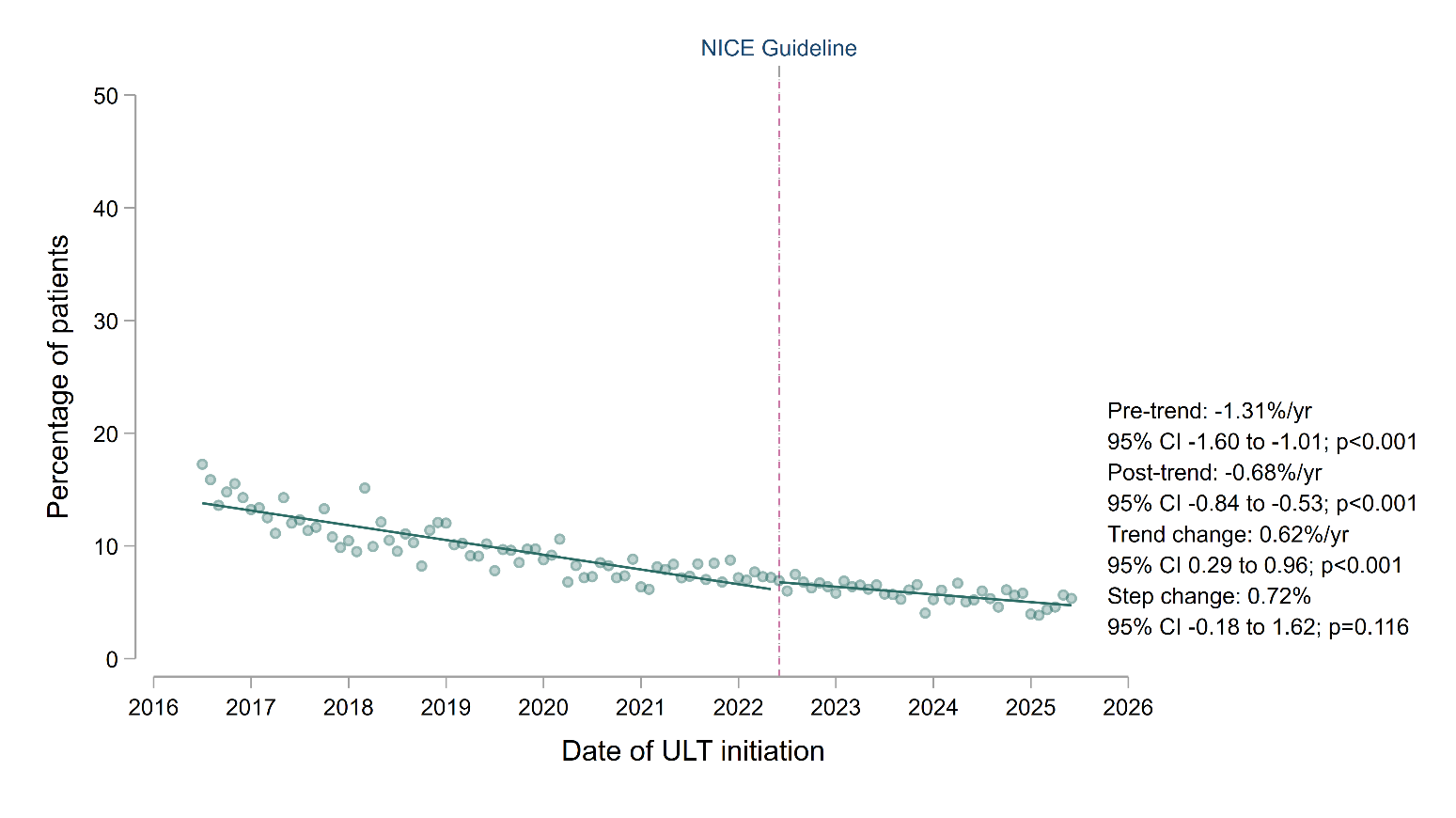
**Supplementary Figure S12.** Proportion of gout individuals initiated on ULT who received high doses of allopurinol or febuxostat as their initial ULT prescription, comparing trends before and after the publication of the NICE Gout Management Guideline in June 2022. Individual data points represent the mean proportion receiving high doses of allopurinol (300 mg tablets) or febuxostat (120 mg tablets) as their initial ULT prescription. Trend lines were fitted before and after publication of the NICE guideline, with annualised coefficients presented as percentage-point change per year, alongside the change in trends (after vs. before publication) and the immediate step change at the date of publication.

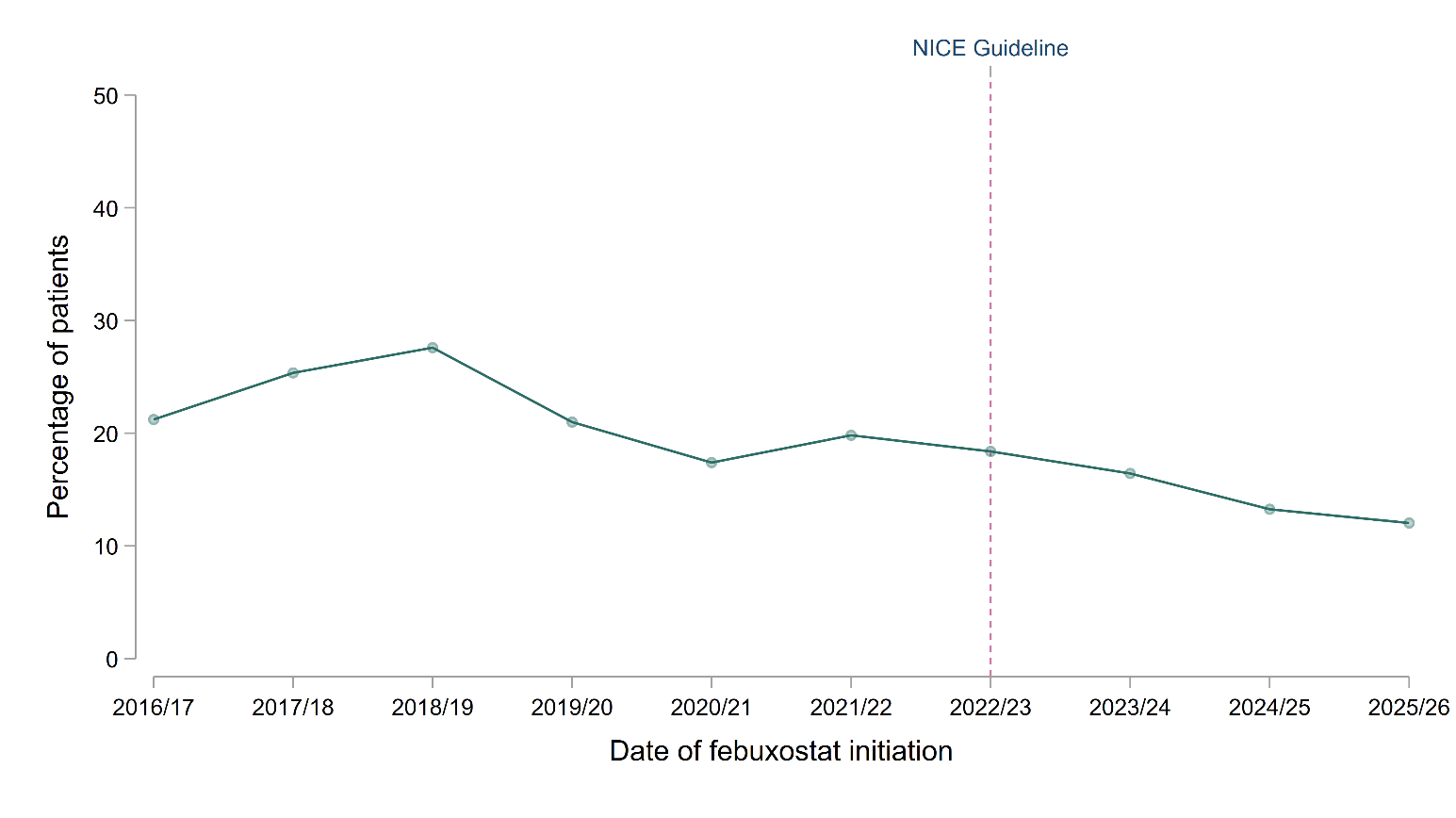
**Supplementary Figure S13.** Temporal trends in the proportion of individuals with gout initiated on febuxostat who had a pre-existing history of major cardiovascular disease (coronary heart disease and/or stroke or transient ischaemic attack). Data were annualised due to small numbers of individuals initiating febuxostat, with each data point representing a 1-year period from 1^st^ July to 30^th^ June. The vertical dashed line represents the publication of the NICE Gout Management Guideline in June 2022.

**Supplementary Figure S14.** Proportion of individuals with gout initiated on ULT after diagnosis who were prescribed colchicine, NSAIDs or corticosteroids on the same day as their initial ULT prescription and again 3 months later (used a proxy for flare prophylaxis), comparing trends before and after the publication of the NICE Gout Management Guideline in June 2022. Temporal trends are shown in the top panel (A) with 3-monthly rolling averages of the current, preceding and subsequent months. In the bottom panel (B), trends were fitted before and after publication of the NICE guideline using single-group ITSA, with annualised coefficients presented as percentage-point change per year, alongside the change in trends (after vs. before publication) and the immediate step change at the date of publication.

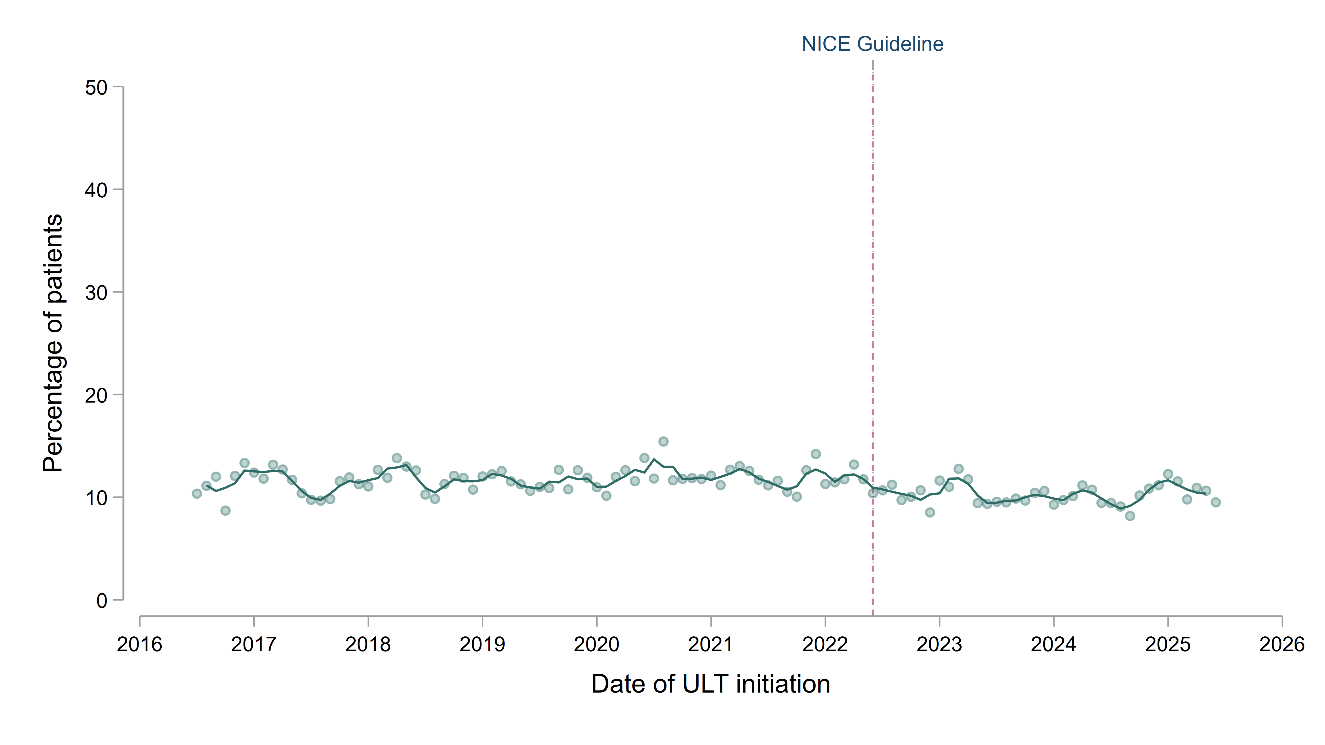

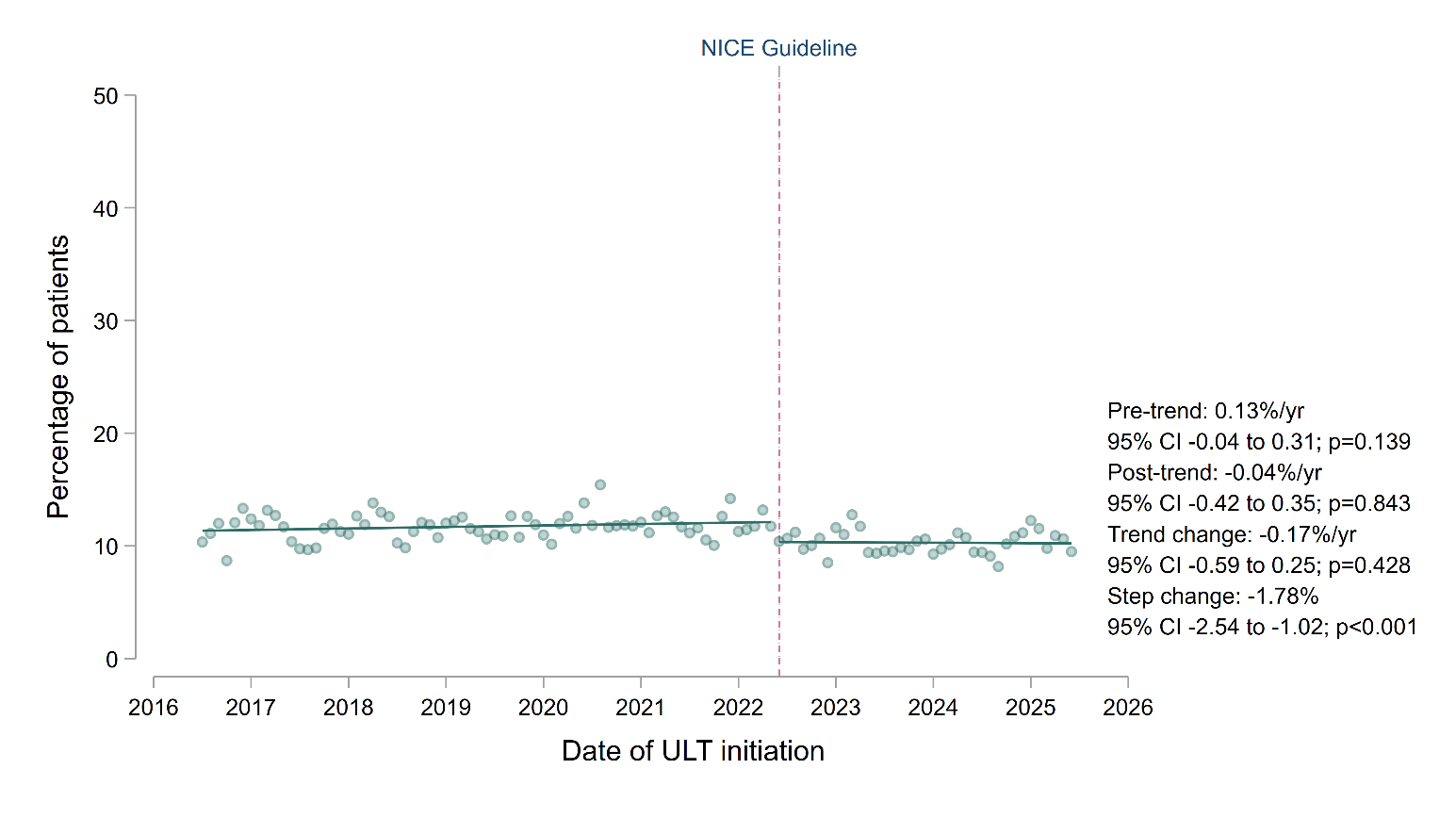

A

B

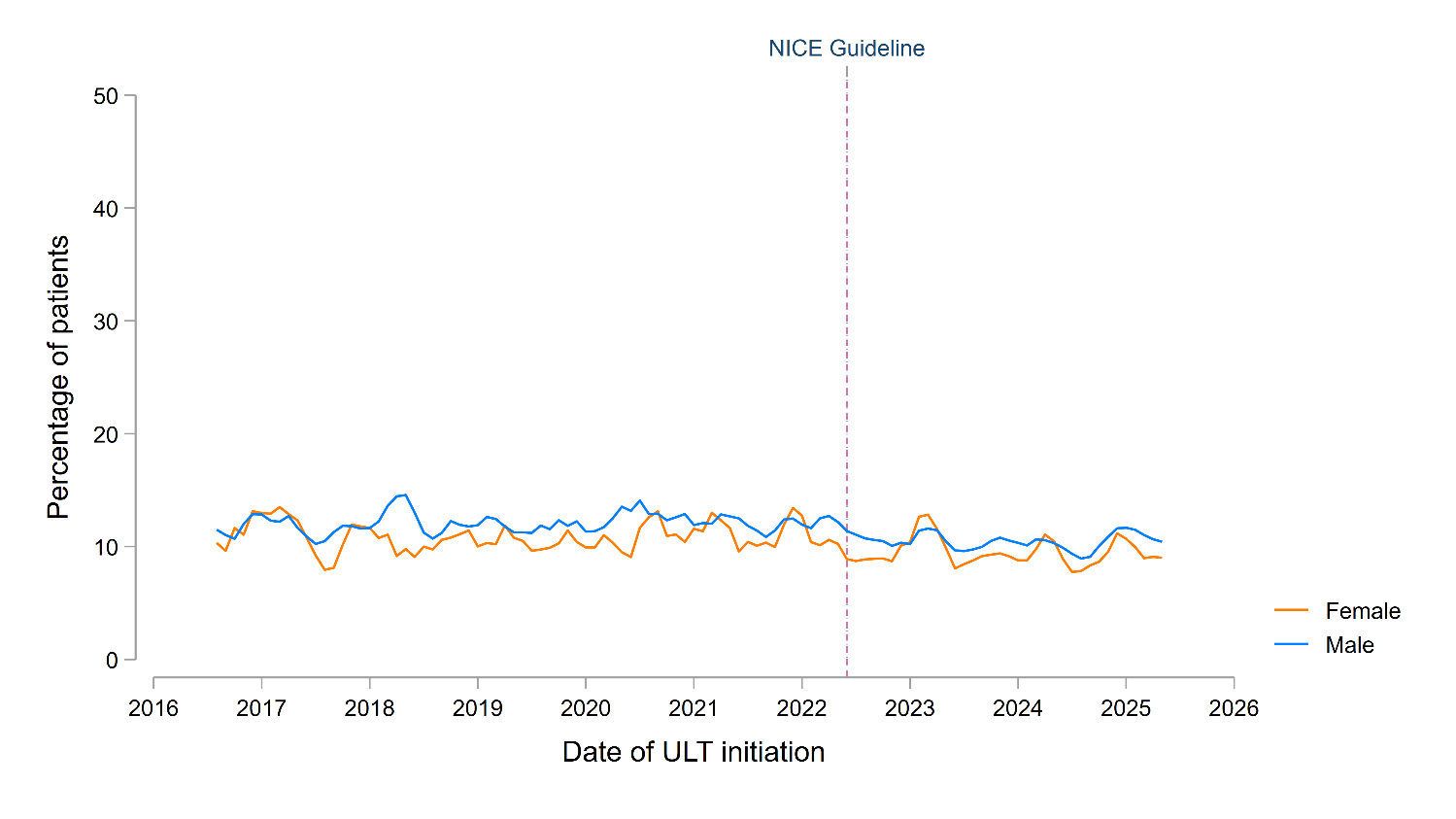
**Supplementary Figure S15.** Temporal trends in the proportion of individuals with gout initiated on ULT after diagnosis who were prescribed colchicine, NSAIDs or corticosteroids on the same day as their initial ULT prescription and again 3 months later, separated by sex. Individual trend lines represent 3-monthly rolling averages of the current, preceding and subsequent months. The vertical dashed line represents the publication of the NICE Gout Management Guideline in June 2022.

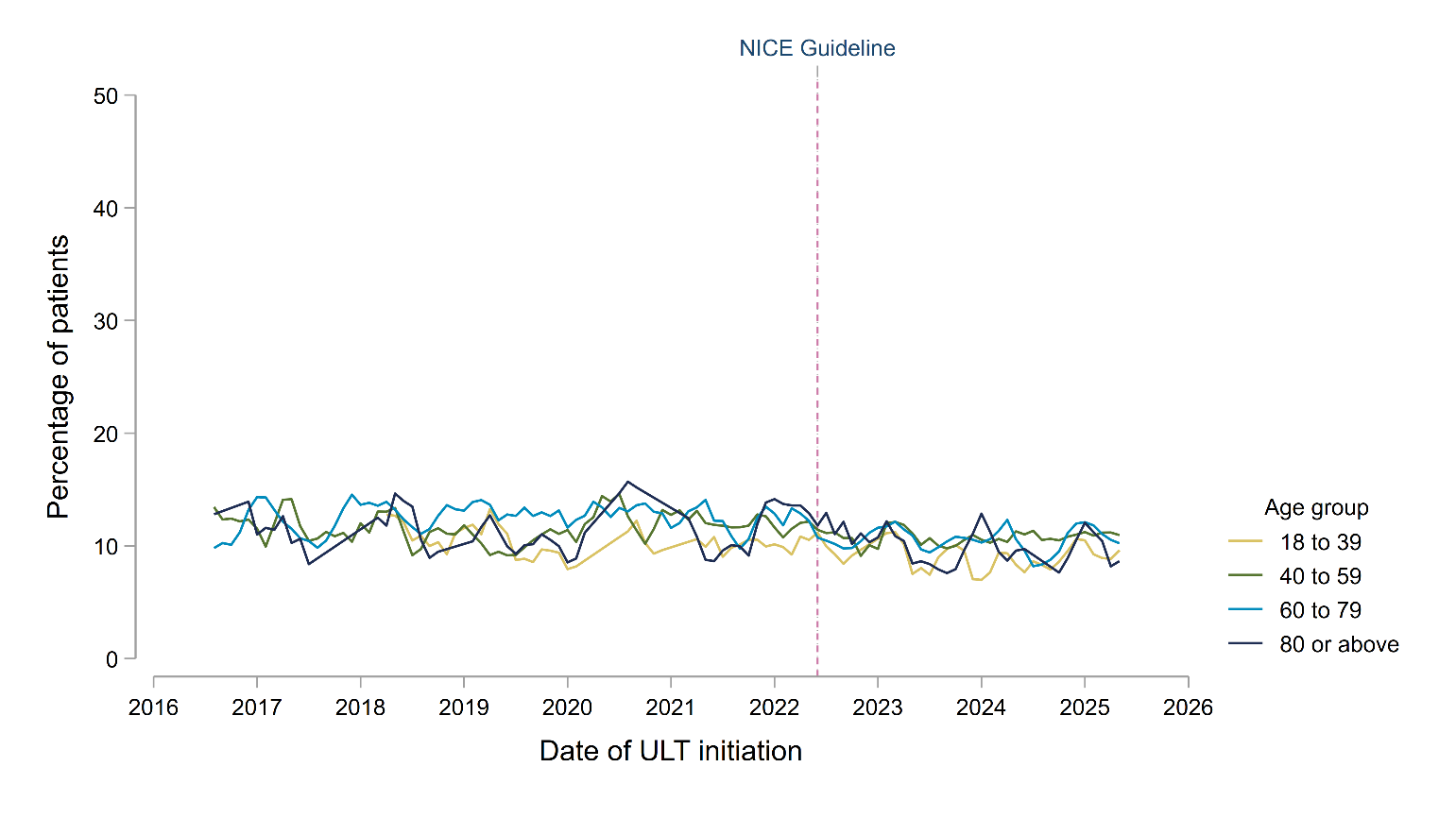
**Supplementary Figure S16.** Temporal trends in the proportion of individuals with gout initiated on ULT after diagnosis who were prescribed colchicine, NSAIDs or corticosteroids on the same day as their initial ULT prescription and again 3 months later, separated by age group. Individual trend lines represent 3-monthly rolling averages of the current, preceding and subsequent months. The vertical dashed line represents the publication of the NICE Gout Management Guideline in June 2022.

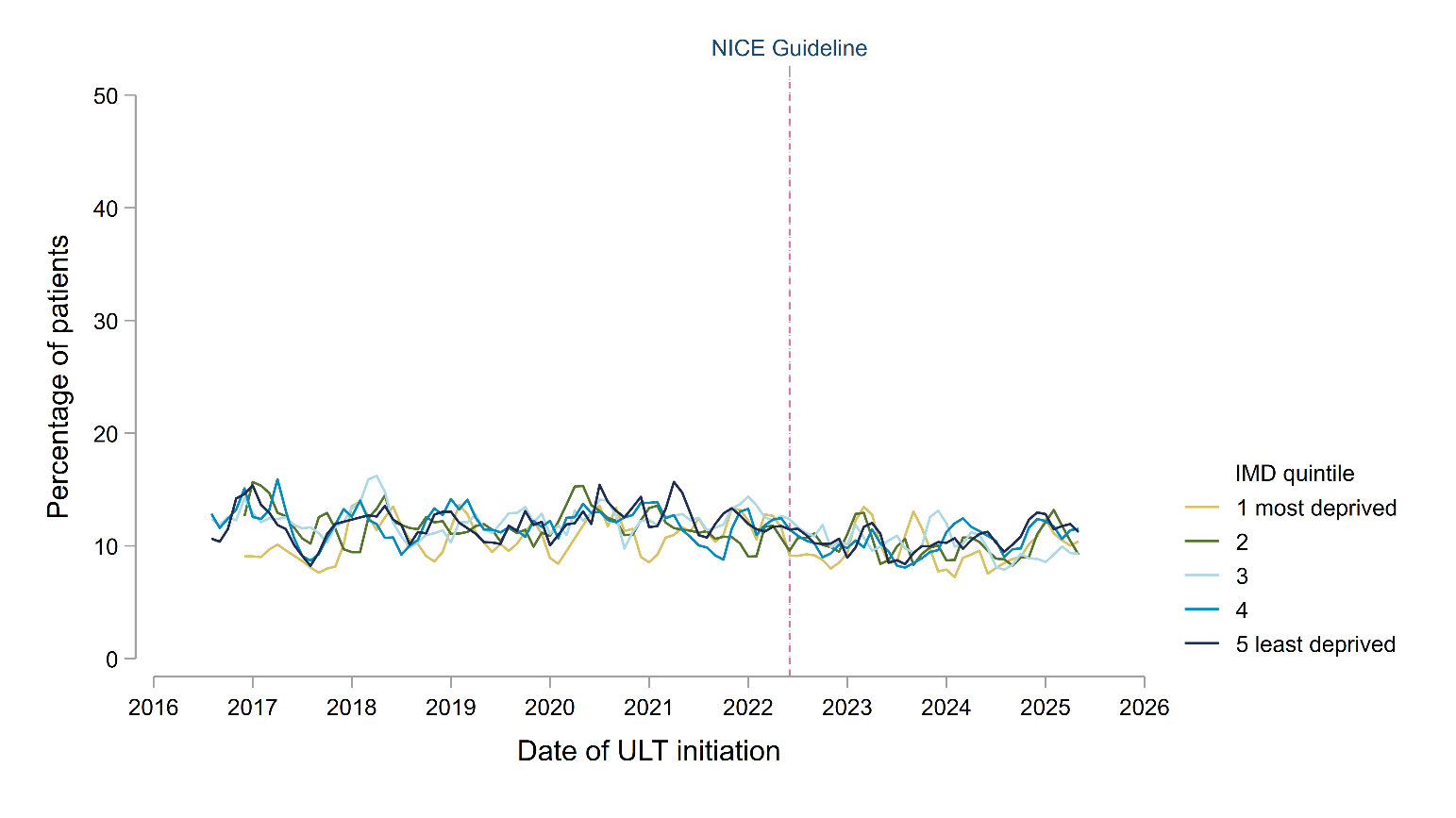
**Supplementary Figure S17.** Temporal trends in the proportion of individuals with gout initiated on ULT after diagnosis who were prescribed colchicine, NSAIDs or corticosteroids on the same day as their initial ULT prescription and again 3 months later, separated by socioeconomic status (defined by quintiles of index of multiple deprivation [IMD]). Individual trend lines represent 3-monthly rolling averages of the current, preceding and subsequent months. The vertical dashed line represents the publication of the NICE Gout Management Guideline in June 2022.

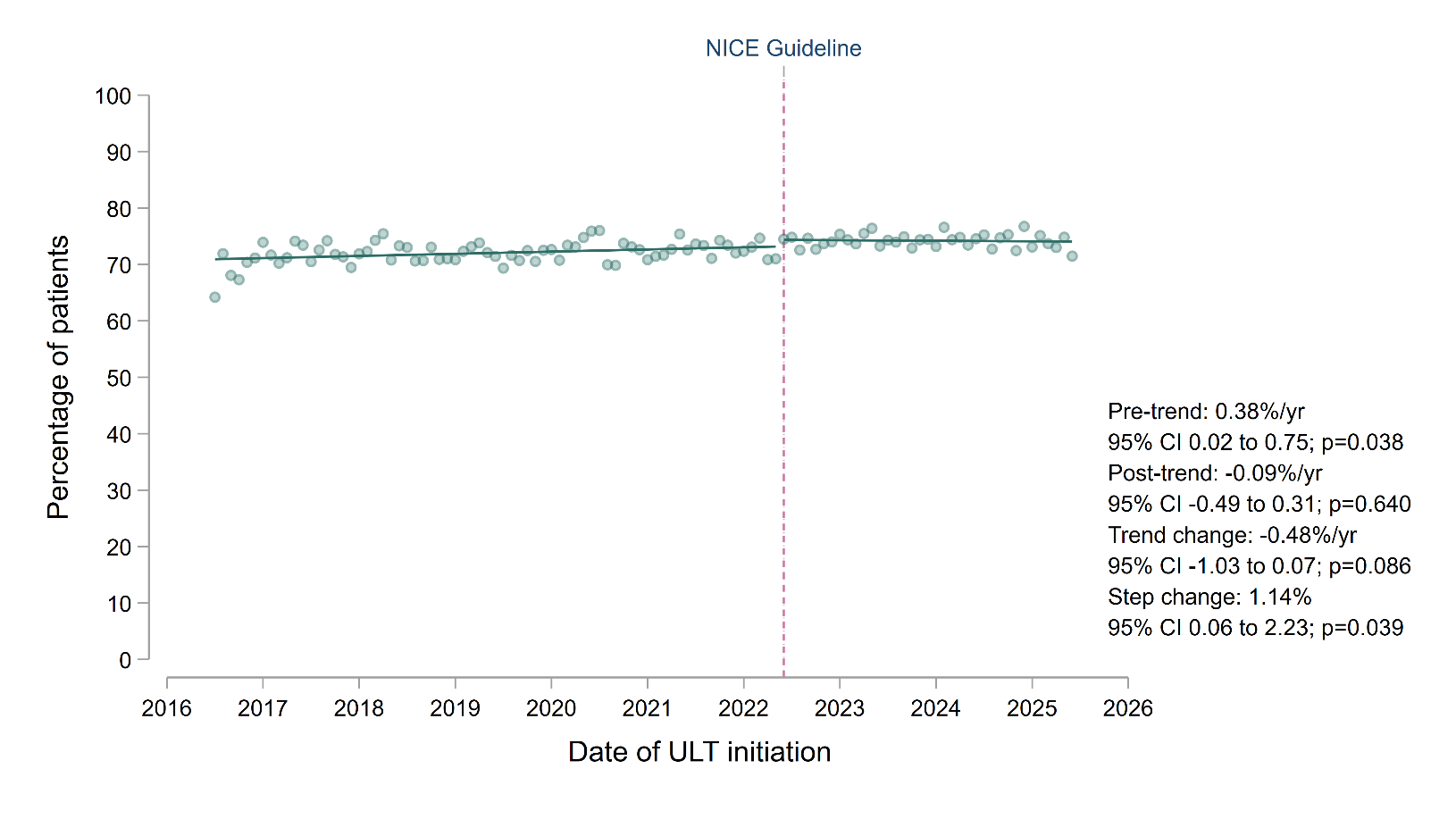
**Supplementary Figure S18.** Interrupted time-series showing the proportion of individuals with gout initiated on ULT who were still prescribed a ULT medication after 12 months (+/- 60 days), comparing trends before and after the publication of the NICE Gout Management Guideline in June 2022. Individual data points represent the mean proportion still prescribed a ULT medication. Trend lines were fitted before and after publication of the NICE guideline, with annualised coefficients presented as percentage-point change per year, alongside the change in trends (after vs. before publication) and the immediate step change at the date of publication.

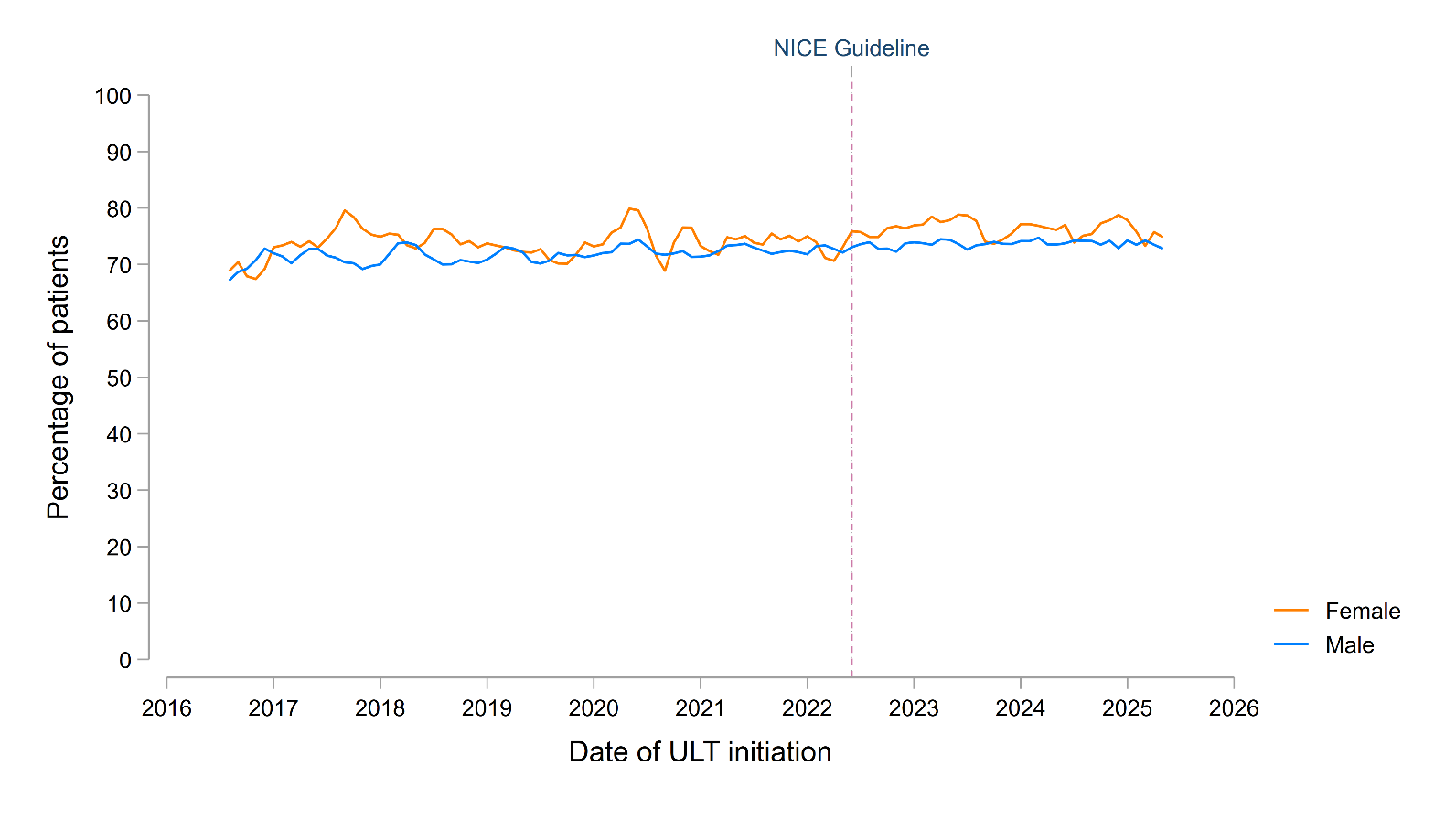
**Supplementary Figure S19.** Temporal trends in the proportion of individuals with gout initiated on ULT who were still prescribed a ULT medication after 12 months, separated by sex. Individual trend lines represent 3-monthly rolling averages of the current, preceding and subsequent months. The vertical dashed line represents the publication of the NICE Gout Management Guideline in June 2022.

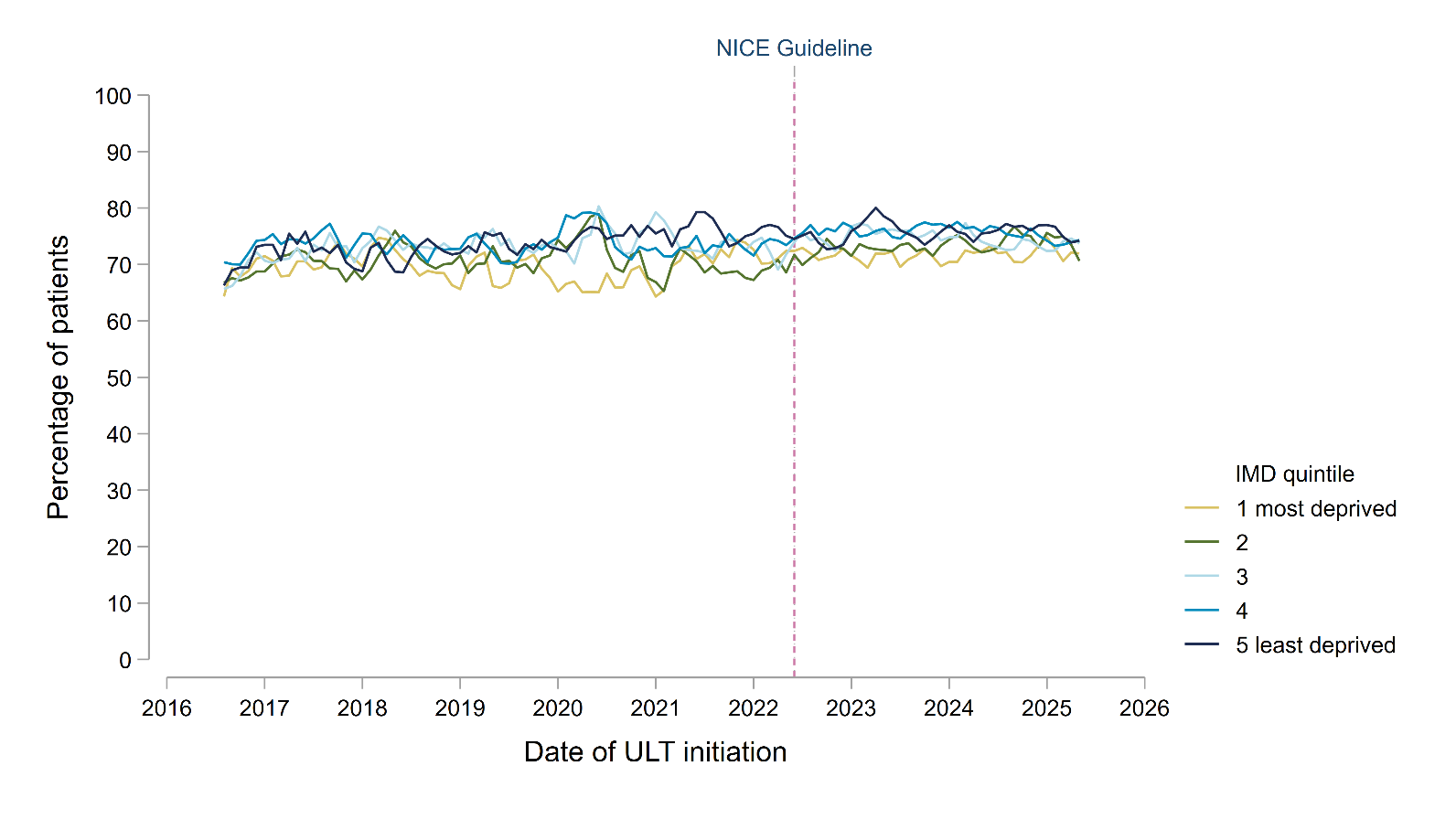
**Supplementary Figure S20.** Temporal trends in the proportion of individuals with gout initiated on ULT who were still prescribed a ULT medication after 12 months, separated by socioeconomic status (defined by quintiles of index of multiple deprivation [IMD]). Individual trend lines represent 3-monthly rolling averages of the current, preceding and subsequent months. The vertical dashed line represents the publication of the NICE Gout Management Guideline in June 2022.

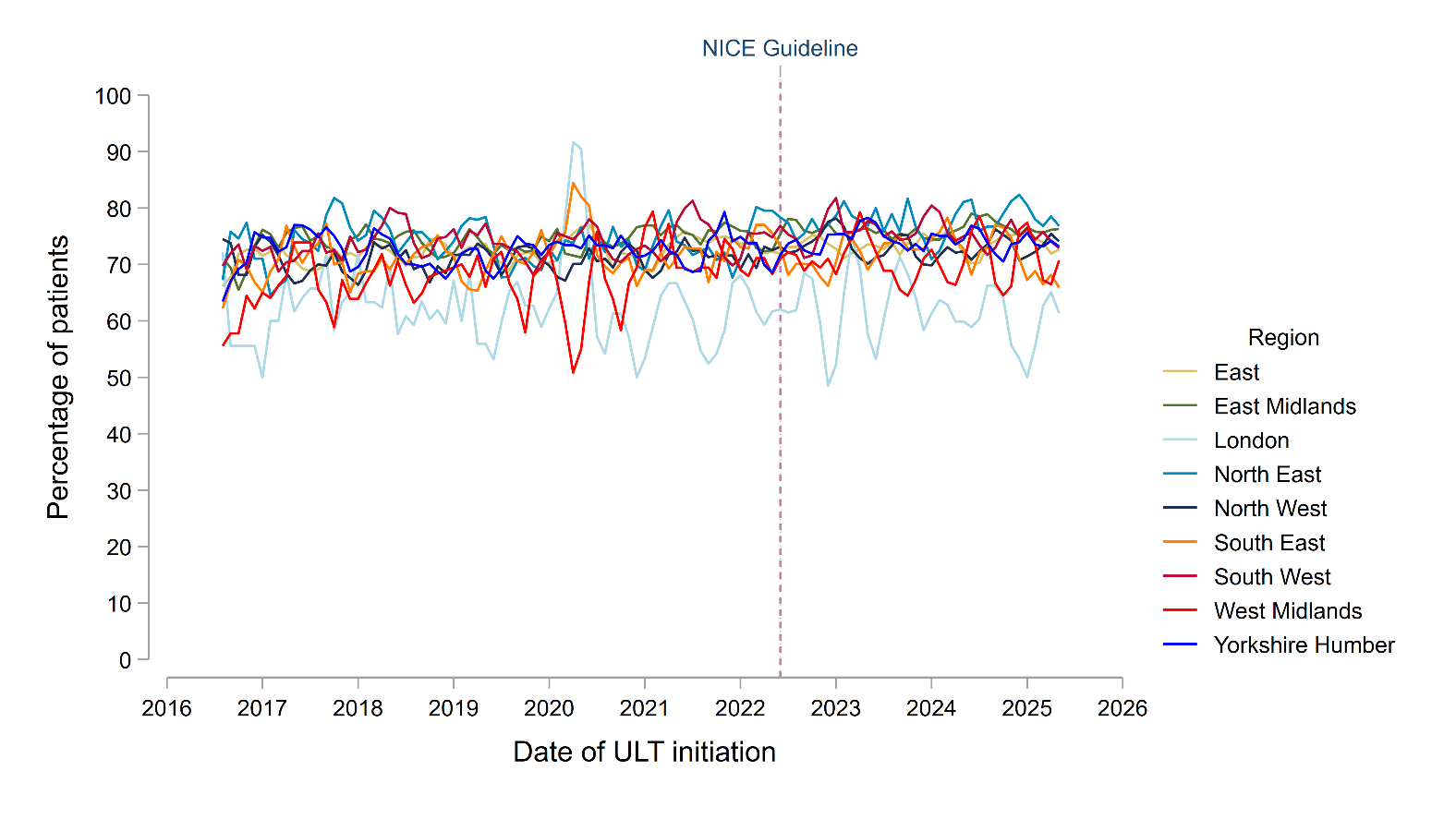
**Supplementary Figure S21.** Temporal trends in the proportion of individuals with gout initiated on ULT who were still prescribed a ULT medication after 12 months, separated by region. Individual trend lines represent 3-monthly rolling averages of the current, preceding and subsequent months. The vertical dashed line represents the publication of the NICE Gout Management Guideline in June 2022.

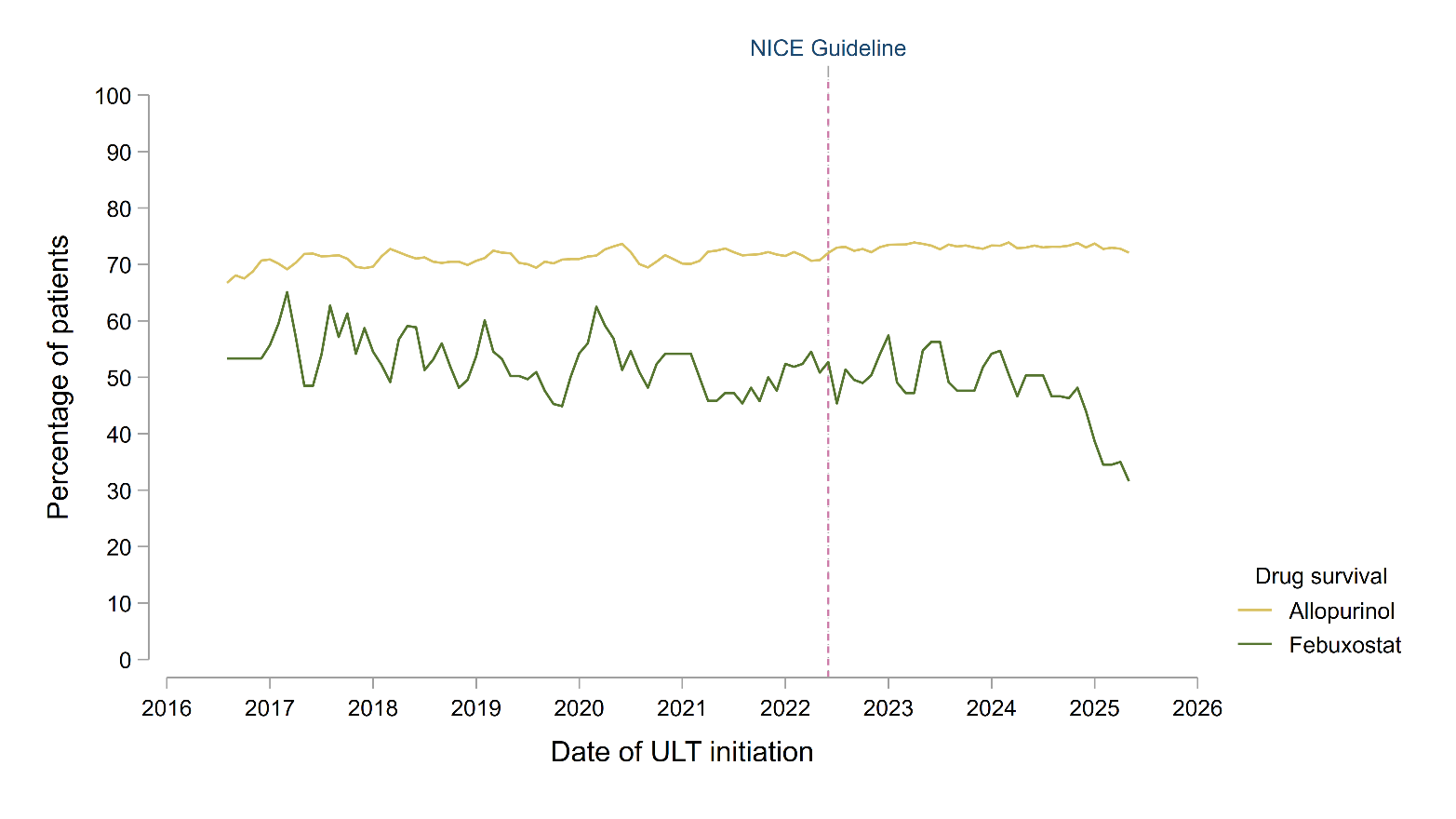
**Supplementary Figure S22.** Temporal trends in the proportion of individuals with gout initiated on allopurinol or febuxostat who were still prescribed the same ULT medication after 12 months. Individual trend lines represent 3-monthly rolling averages of the current, preceding and subsequent months. The vertical dashed line represents the publication of the NICE Gout Management Guideline in June 2022.

**Supplementary Figure S23.** Trends in the proportion of individuals with gout initiating ULT who had (A; top panel) at least one serum urate level and (B; bottom panel) at least two serum urate levels recorded within 12 months of initiating ULT, comparing the periods before and after the publication of the NICE Gout Management Guideline in June 2022. Individual data points represent the mean proportion of individuals initiating ULT who had serum urate levels recorded. Trend lines were fitted before and after publication of the NICE guideline, with annualised coefficients presented as percentage-point change per year, alongside the change in trends (after vs. before publication) and the immediate step change at the date of publication.

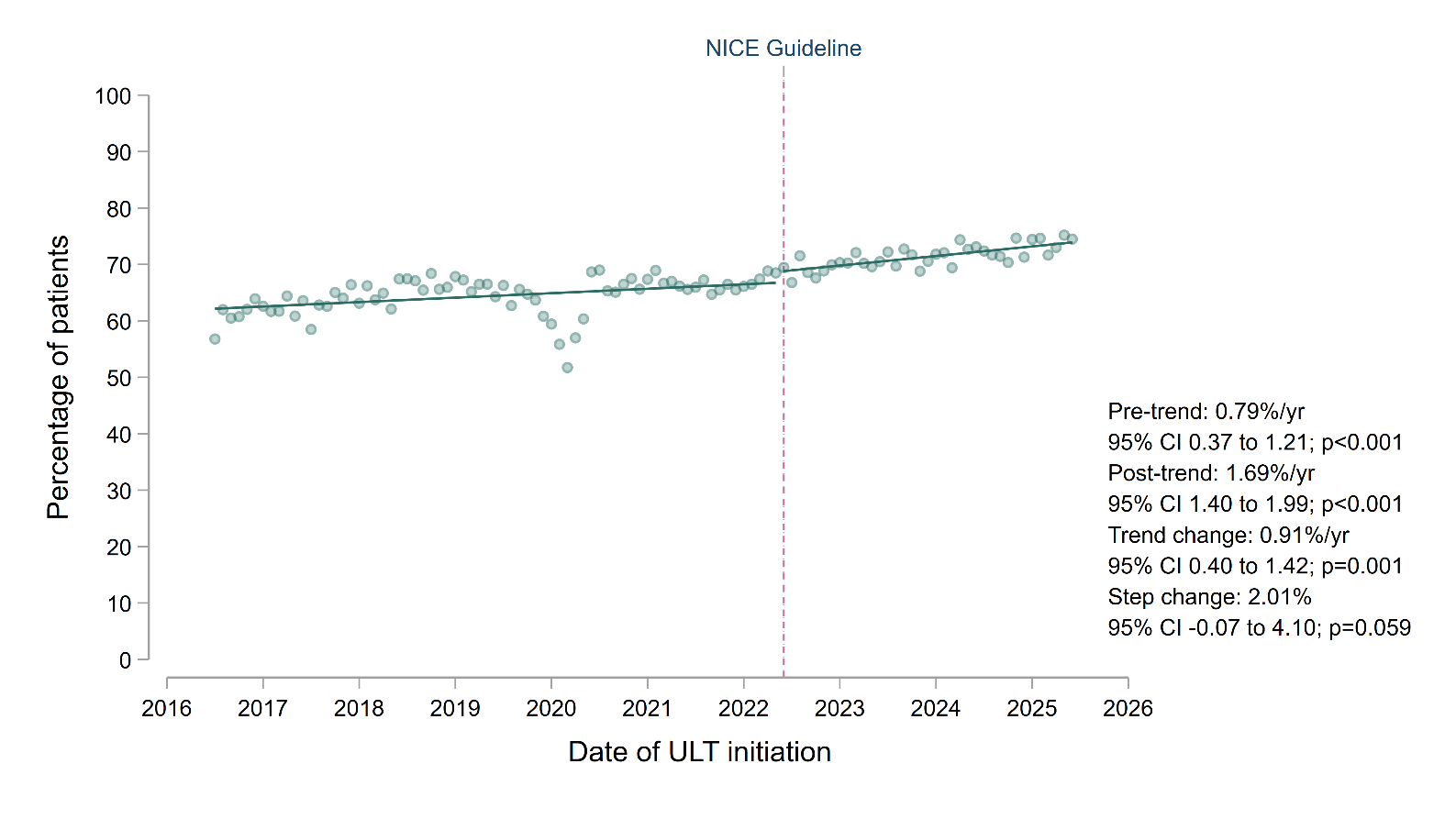

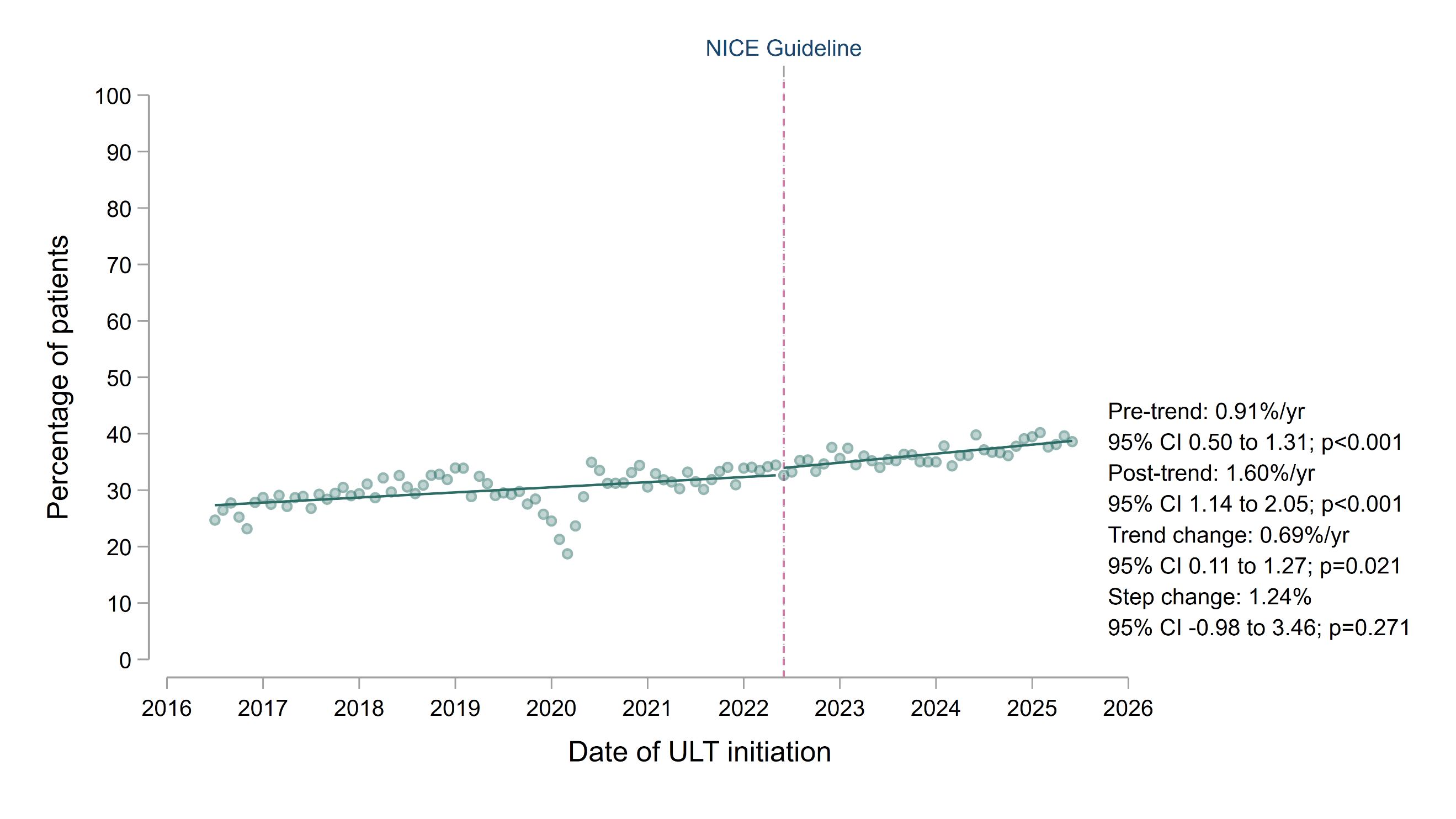

A

B

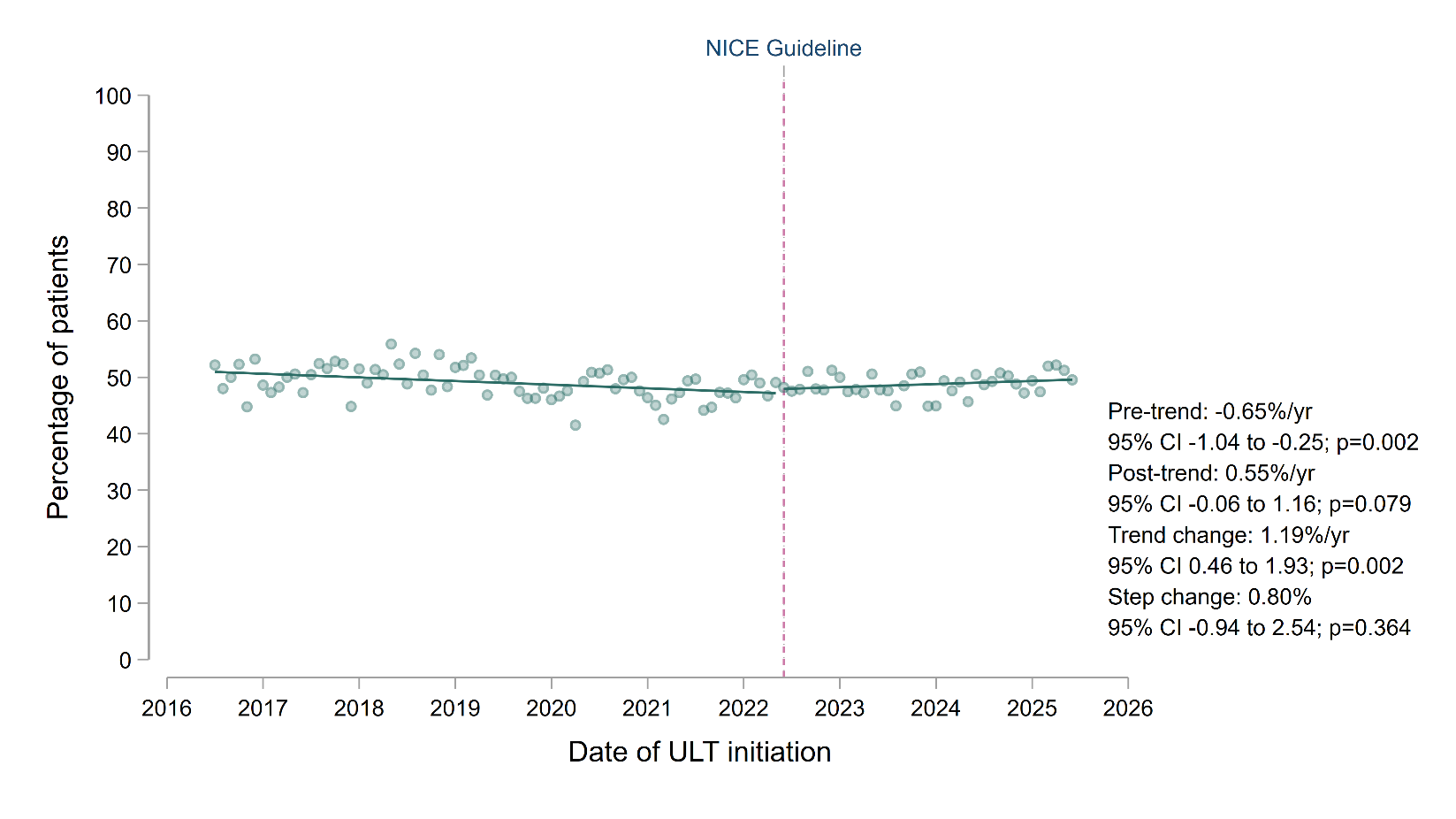
**Supplementary Figure S24.** Temporal trends in the attainment of serum urate levels below 360 micromol/L within 12 months of urate-lowering therapy (ULT) initiation, restricted to individuals who had at least one serum urate measurement within 12 months of initiating ULT. Trends before and after publication of the NICE Gout Management Guideline in June 2022; individual data points represent the mean monthly proportion of individuals achieving urate targets; trend lines were fitted using single-group interrupted time-series before and after publication of the NICE guideline, with annualised coefficients presented as percentage-point change per year, alongside thechange in trends and the immediate step change at the date of publication.

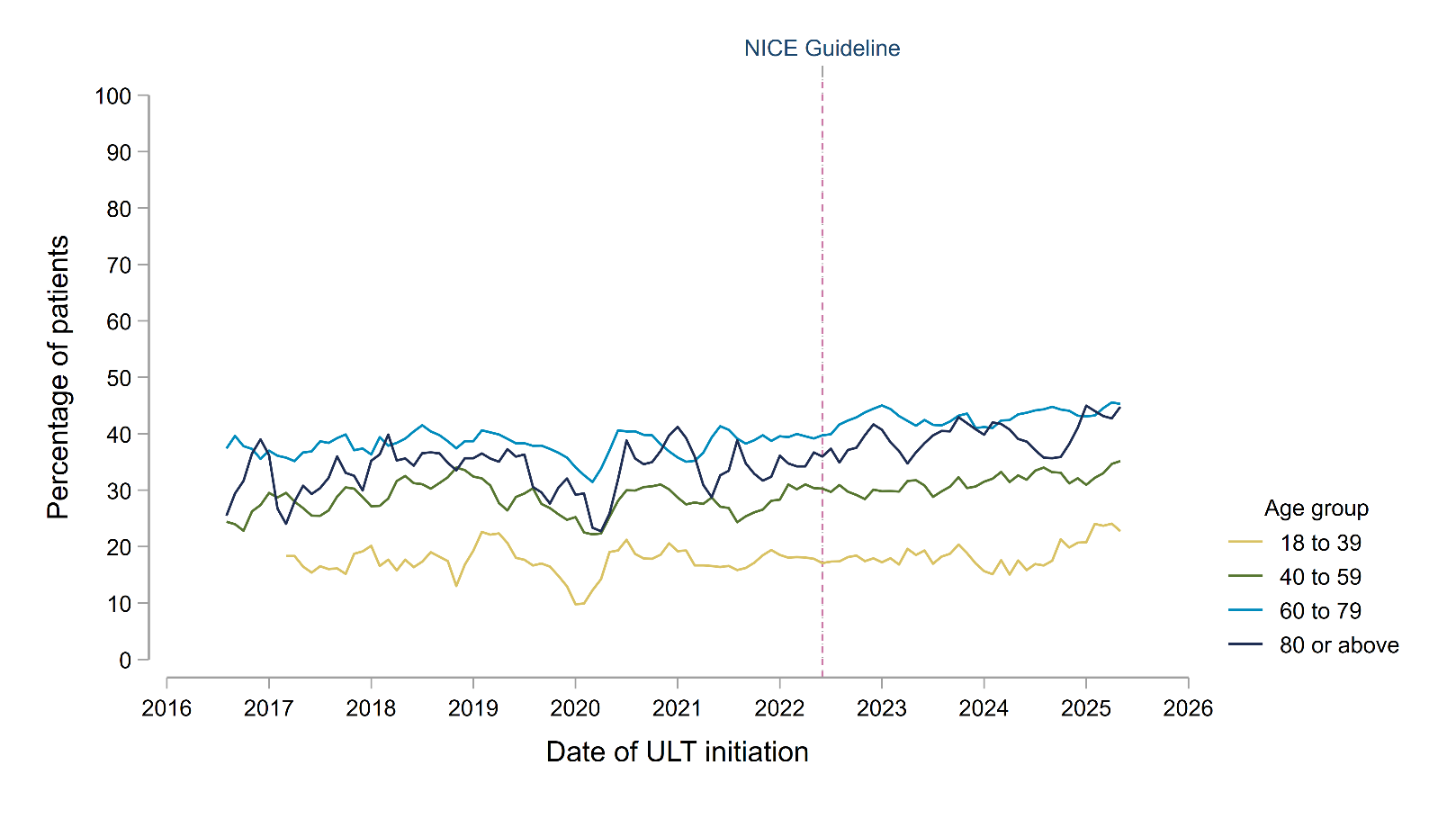
**Supplementary Figure S25.** Temporal trends in the attainment of serum urate levels below 360 micromol/L within 12 months of ULT initiation, separated by age group. Individual trend lines represent 3-monthly rolling averages of the current, preceding and subsequent months. The vertical dashed line represents the publication of the NICE Gout Management Guideline in June 2022.

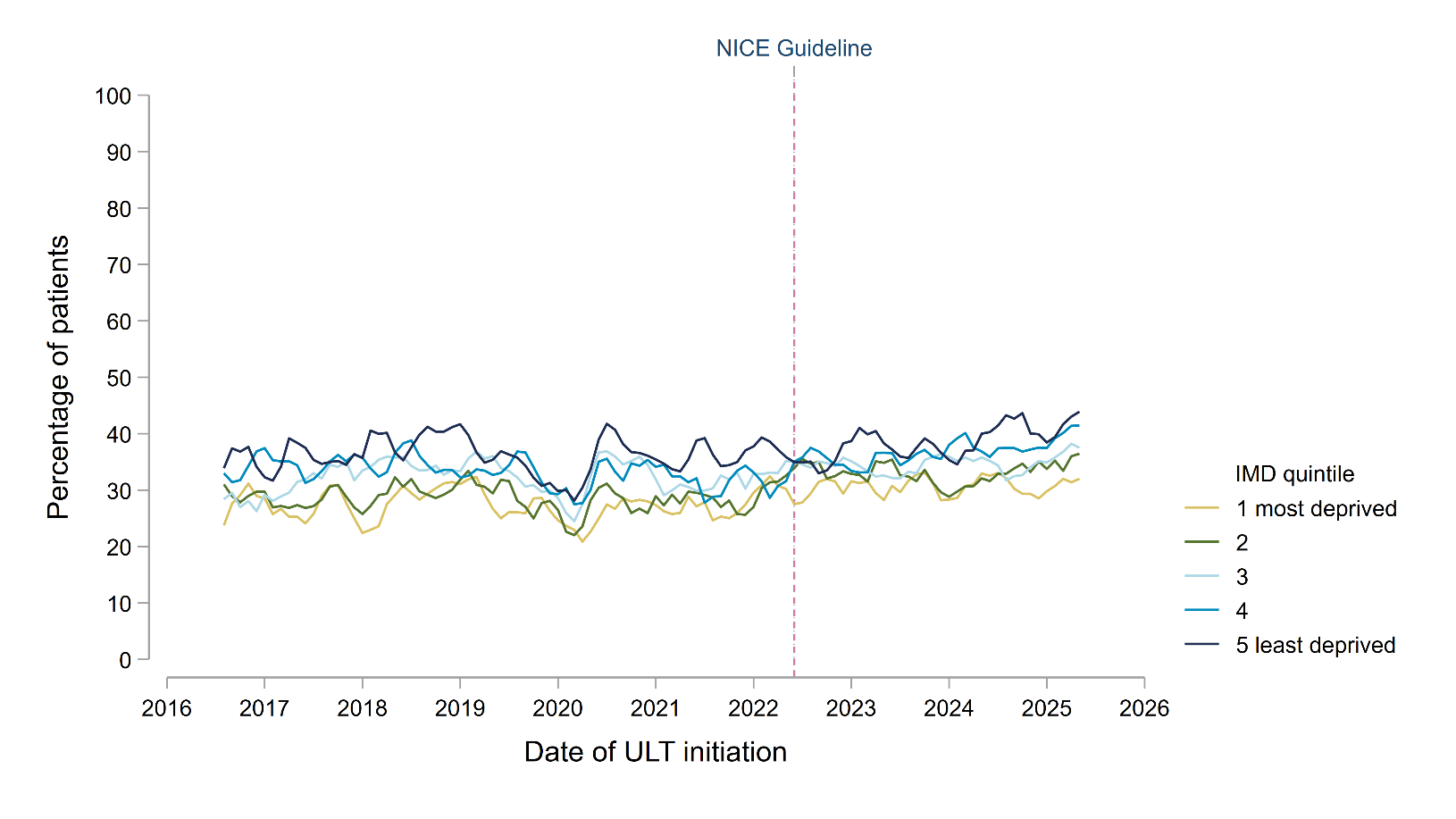
**Supplementary Figure S26.** Temporal trends in the attainment of serum urate levels below 360 micromol/L within 12 months of ULT initiation, separated by socioeconomic status (defined by quintiles of index of multiple deprivation [IMD]). Individual trend lines represent 3-monthly rolling averages of the current, preceding and subsequent months. The vertical dashed line represents the publication of the NICE Gout Management Guideline in June 2022.

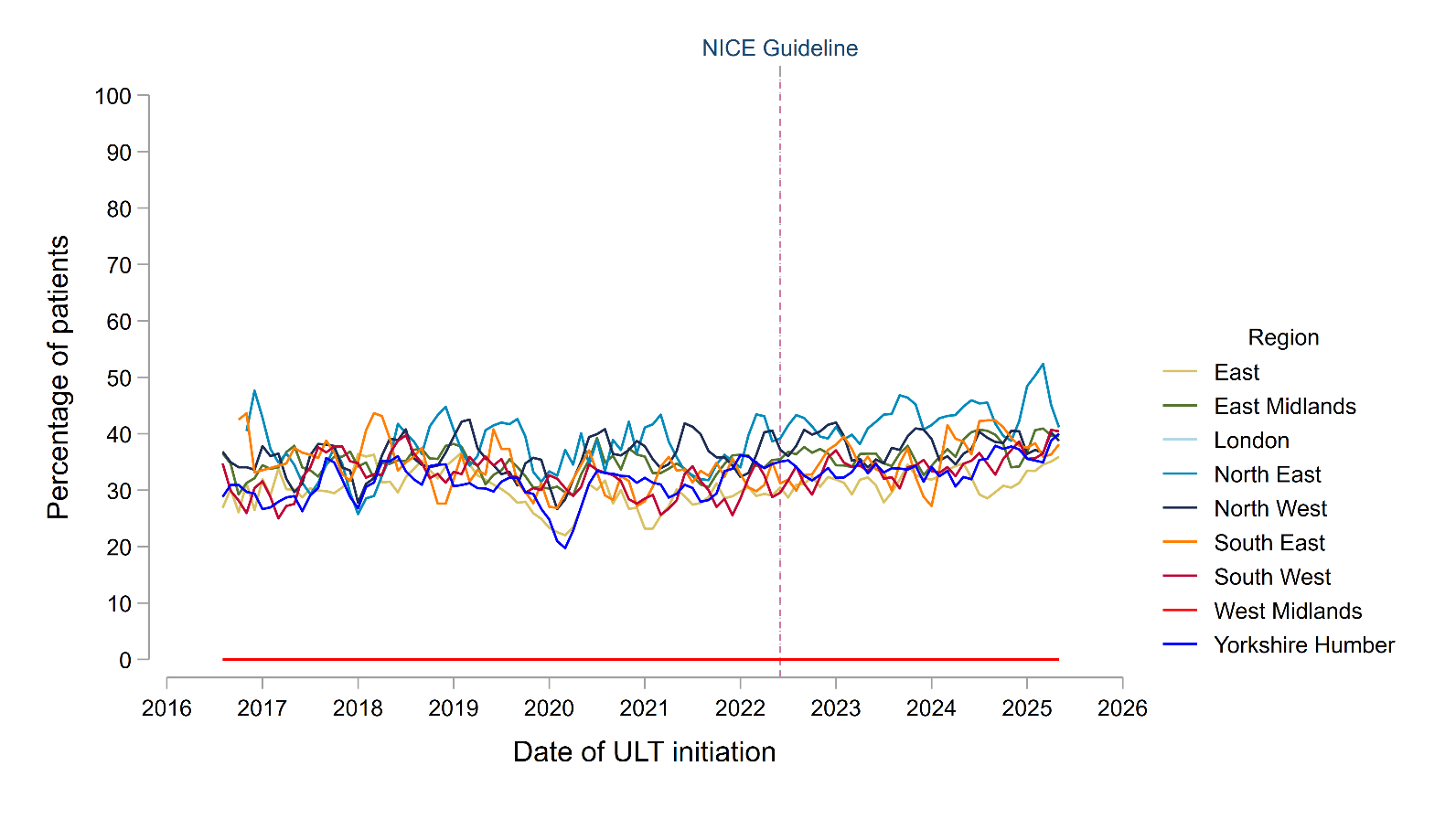
**Supplementary Figure S27.** Temporal trends in the attainment of serum urate levels below 360 micromol/L within 12 months of ULT initiation, separated by region. Data are suppressed for London and West Midlands due to small counts. Individual trend lines represent 3-monthly rolling averages of the current, preceding and subsequent months. The vertical dashed line represents the publication of the NICE Gout Management Guideline in June 2022.

**Supplementary Figure S28.** Trends in the proportion of individuals with gout achieving a urate below 360 micromol/L after initiating ULT who had (A; top panel) at least one repeat urate level within 6 to 12 months of target attainment and (B; bottom panel) remained below 360 micromol/L, comparing the periods before and after the publication of the NICE Gout Management Guideline in June 2022. Individual data points represent the mean proportion meeting these criteria. Trend lines were fitted before and after publication of the NICE guideline, with annualised coefficients presented as percentage-point change per year, alongside the change in trends (after vs. before publication) and the immediate step change at the date of publication.

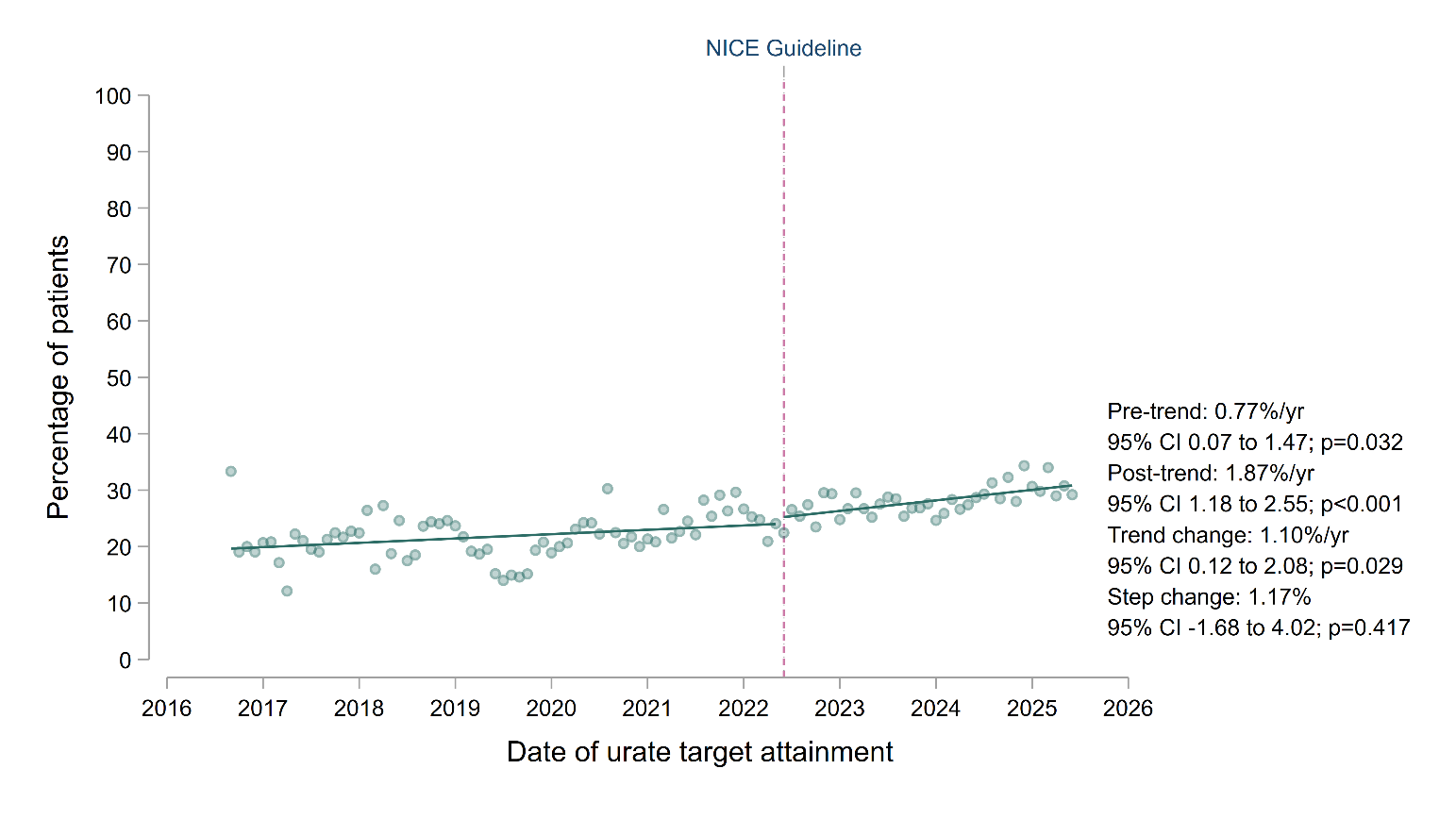

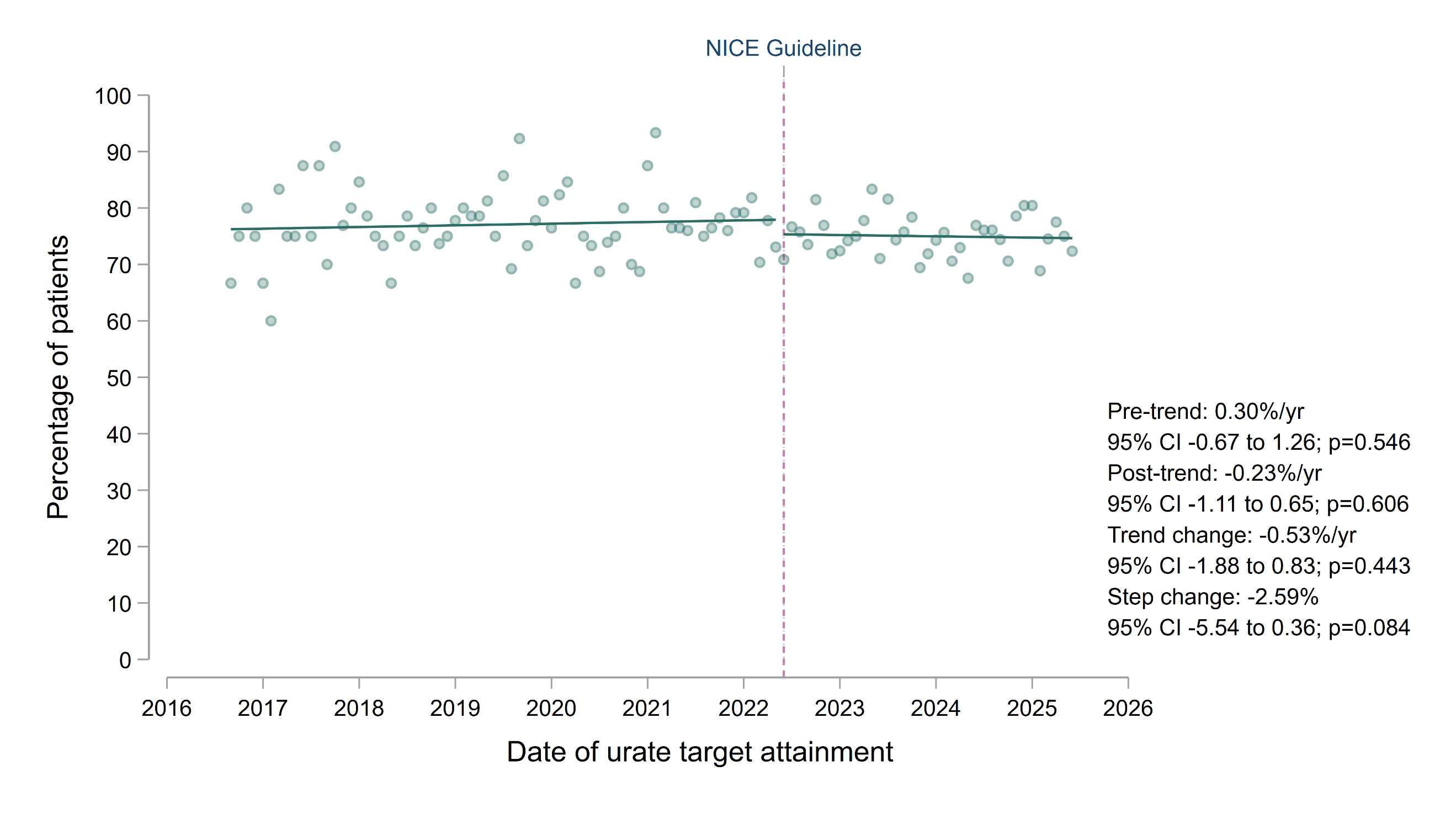

A

B

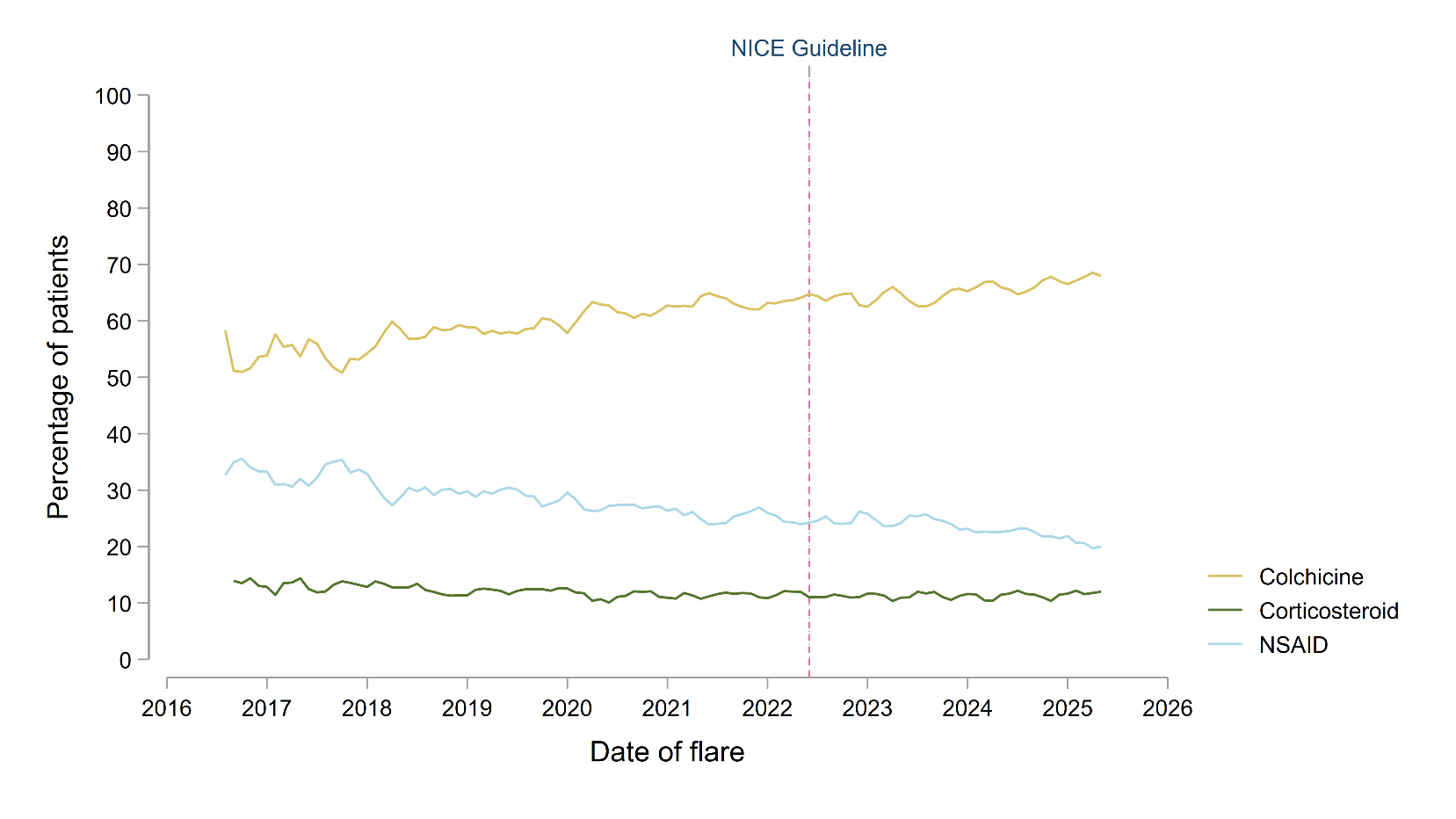
**Supplementary Figure S29.** Trends in the choice of treatment for non-index gout flares. Individual trend lines represent 3-monthly rolling averages of the current, preceding and subsequent months. The vertical dashed line represents the publication of the NICE Gout Management Guideline in June 2022. NSAID: non-steroidal anti-inflammatory drug.

**Supplementary Figure S30.** Sensitivity interrupted time-series analyses (ITSA) comparing trends in the initiation of ULT within 12 months of gout diagnosis. The top panel (A) shows trends using an intervention date 12 months after publication of the NICE gout management guideline (June 2023). The bottom panel (B) shows trends using an intervention date 12 months before publication (June 2021). Trend lines were fitted using single-group ITSA, with annualised coefficients presented as percentage-point change per year, alongside the change in trends (after vs. before the intervention dates) and the immediate step change at the intervention date.

A

B

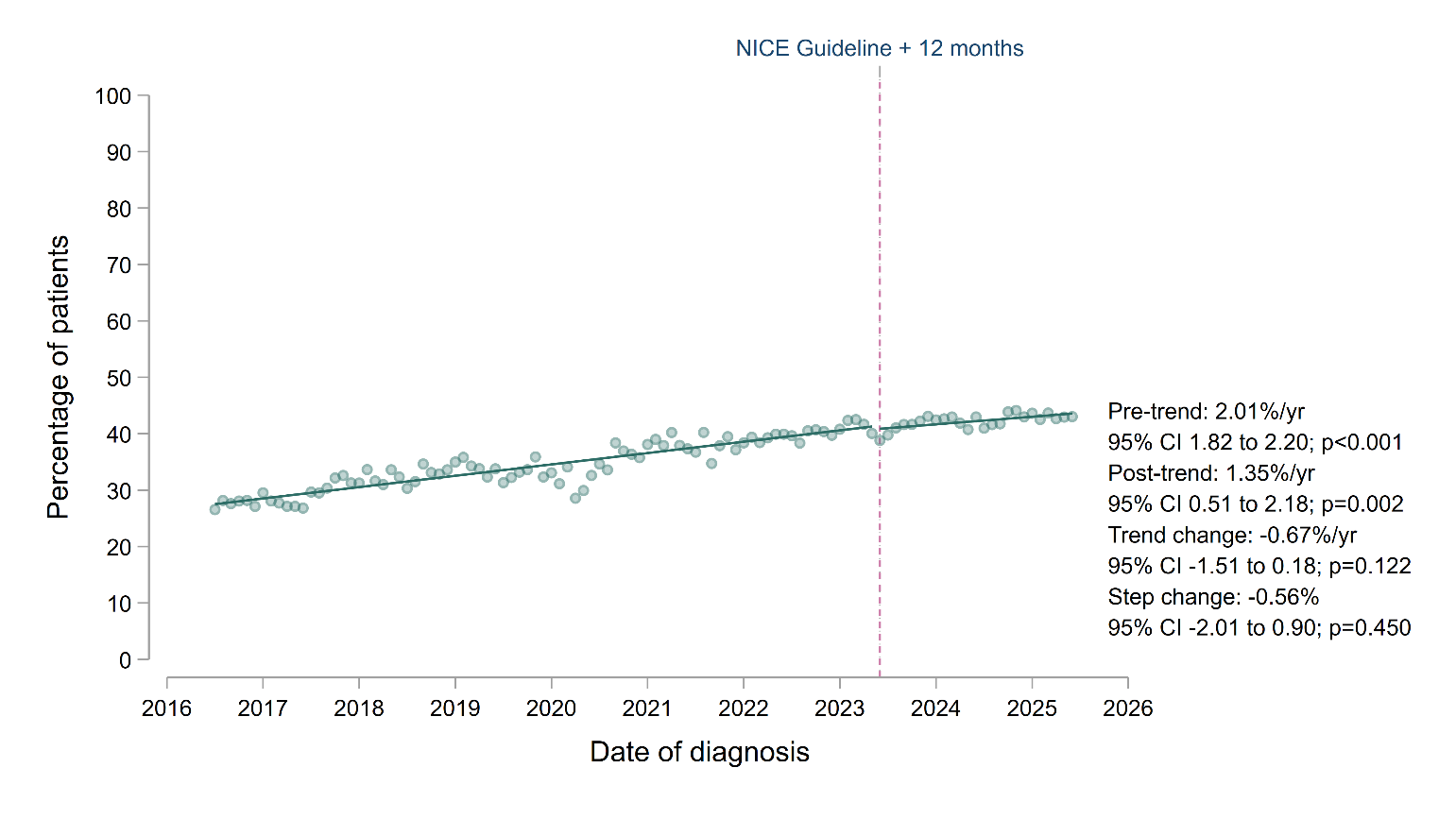

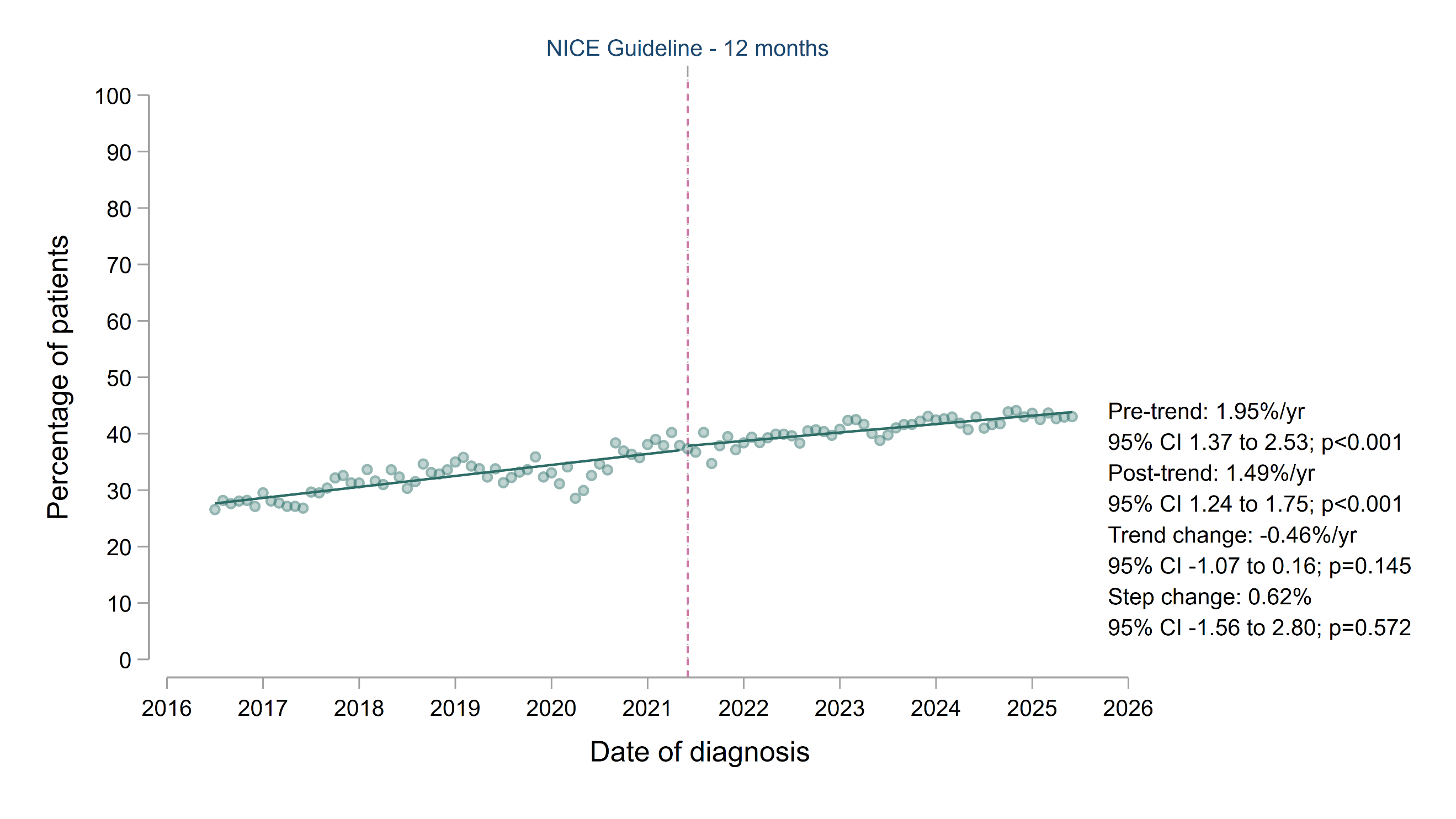

**Supplementary Figure S31.** Sensitivity interrupted time-series analyses (ITSA) comparing trends in the attainment of serum urate levels below 360 micromol/L within 12 months of ULT initiation. The top panel (A) shows trends using an intervention date 12 months after publication of the NICE gout management guideline (June 2023). The bottom panel (B) shows trends using an intervention date 12 months before publication (June 2021). Trend lines were fitted using single-group ITSA, with annualised coefficients presented as percentage-point change per year, alongside the change in trends (after vs. before the intervention dates) and the immediate step change at the intervention date.

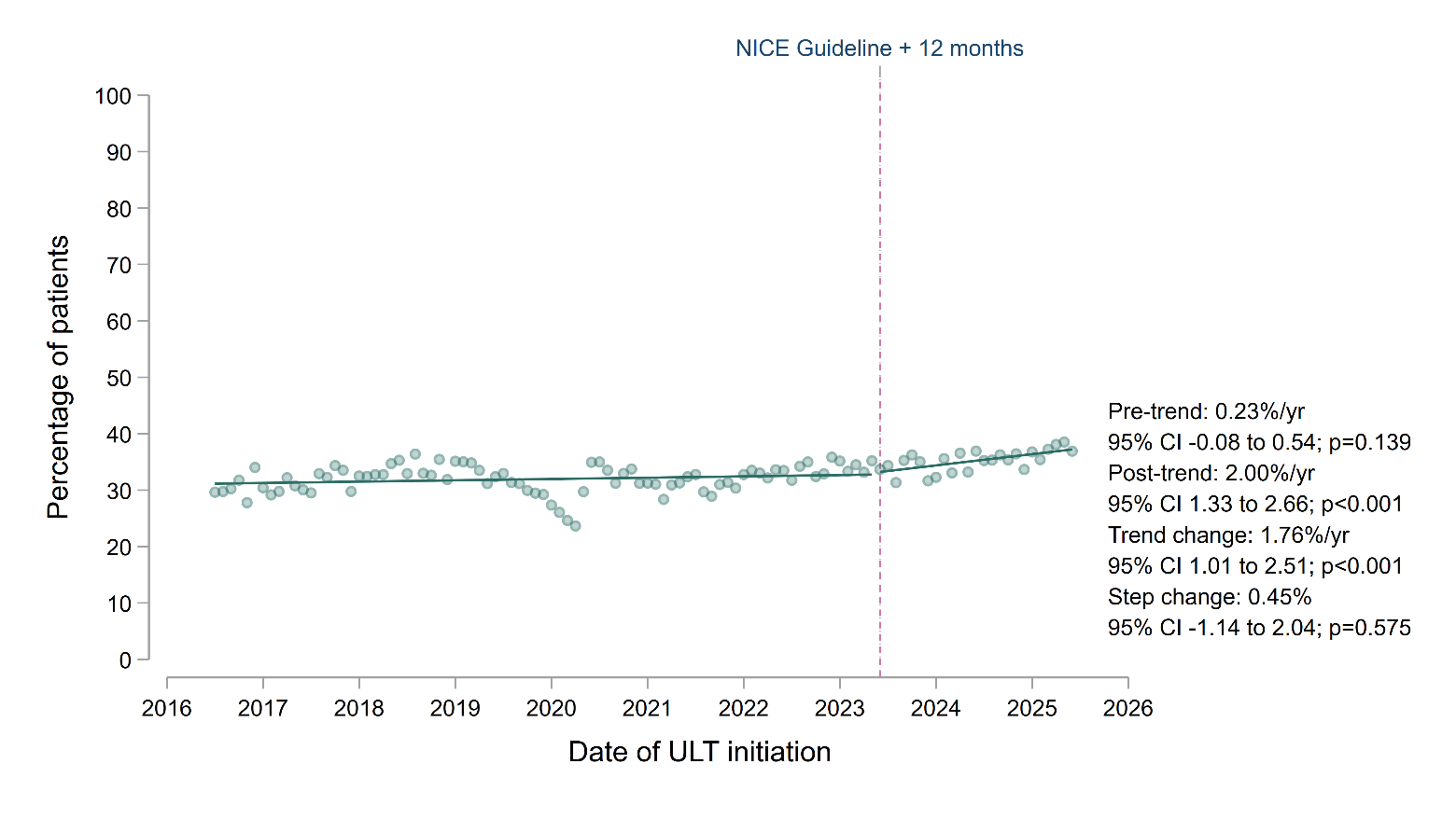

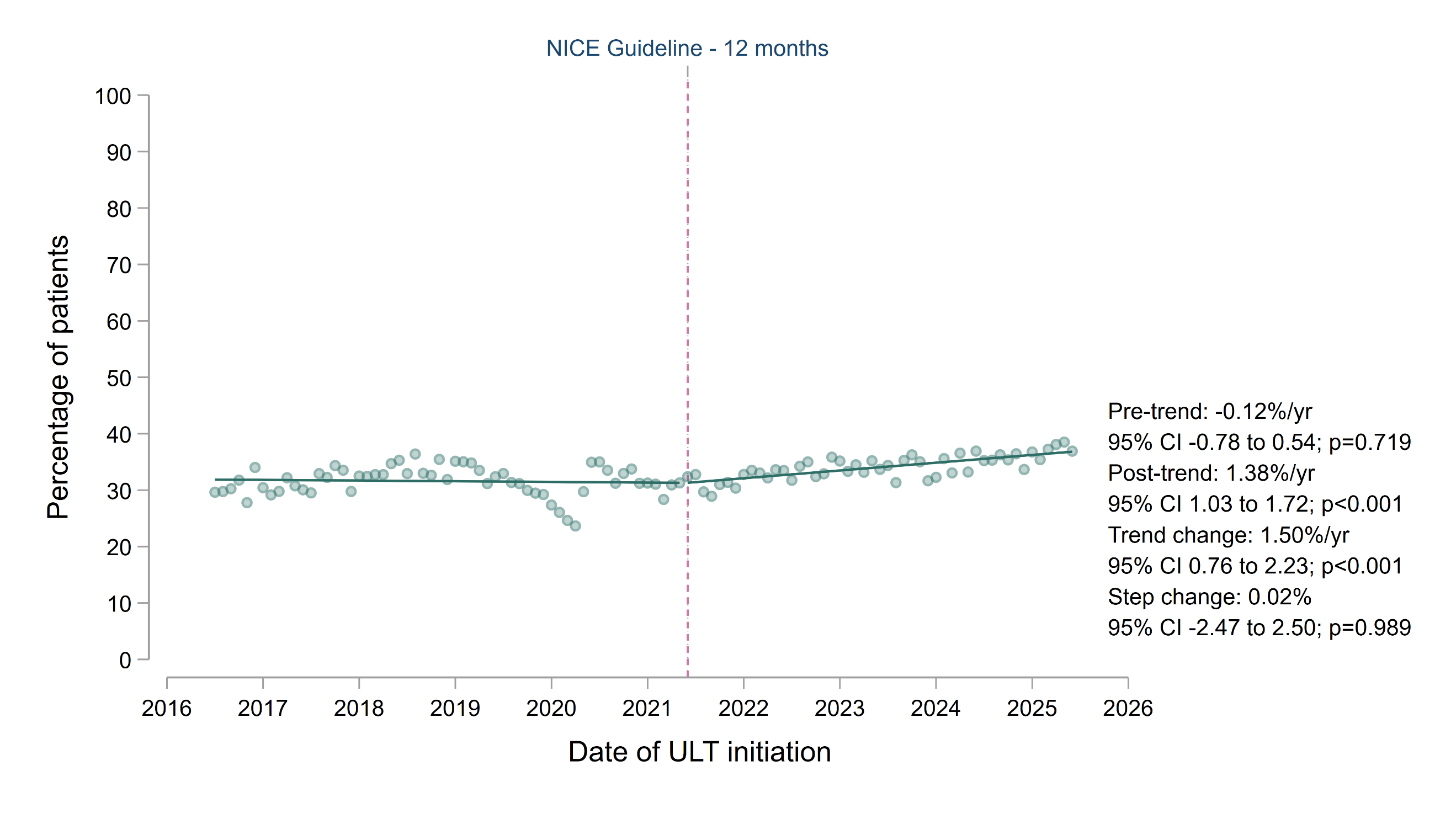

A

B

| **Baseline characteristic** | **Reference population** | **Individuals with gout** |
| --- | --- | --- |
|  | **n=22,618,945** | **n=262,925** |
| Mean age, years (SD) | 46.4 (18.8) | 60.7 (16.3) |
| Age group, years |  |  |
| 18-39 | 9,536,035 (42.2%) | 31,155 (11.9%) |
| 40-59 | 7,200,330 (31.8%) | 91,240 (34.7%) |
| 60-79 | 4,667,350 (20.6%) | 104,355 (39.7%) |
| 80+ | 1,215,225 (5.4%) | 36,175 (13.8%) |
| Sex |  |  |
| Female | 11,409,495 (50.4%) | 68,715 (26.1%) |
| Male | 11,209,450 (49.6%) | 194,205 (73.9%) |
| Ethnicity |  |  |
| White | 17,976,055 (79.5%) | 234,445 (89.2%) |
| Asian/Asian British | 1,875,605 (8.3%) | 14,570 (5.5%) |
| Black/Black British | 692,680 (3.1%) | 4,335 (1.7%) |
| Mixed | 327,455 (1.5%) | 1,895 (0.7%) |
| Chinese or other | 677,340 (3.0%) | 3,160 (1.2%) |
| Not known | 1,069,810 (4.7%) | 4,515 (1.7%) |
| Index of multiple deprivation quintiles |  |  |
| 1 most deprived | 4,333,080 (19.2%) | 44,505 (16.9%) |
| 2 | 4,504,815 (19.9%) | 47,320 (18.0%) |
| 3 | 4,843,335 (21.4%) | 57,030 (21.7%) |
| 4 | 4,427,105 (19.6%) | 55,790 (21.2%) |
| 5 least deprived | 4,002,565 (17.7%) | 52,745 (20.1%) |
| Not known | 508,040 (2.3%) | 5,540 (2.1%) |
| Body mass index |  |  |
| Underweight (<18.5) | 459,150 (2.0%) | 1,310 (0.5%) |
| Normal (18.5-24.9) | 6,479,250 (28.7%) | 35,585 (13.5%) |
| Overweight (25-29.9) | 5,693,765 (25.2%) | 85,145 (32.4%) |
| Obese I (30-34.9) | 2,737,055 (12.1%) | 61,880 (23.5%) |
| Obese II (35-39.9) | 1,019,015 (4.5%) | 26,155 (10.0%) |
| Obese III (40+) | 570,330 (2.5%) | 15,675 (6.0%) |
| Not known | 5,660,375 (25.0%) | 37,170 (14.1%) |
| Smoking status |  |  |
| Never | 10,754,785 (47.6%) | 102,550 (39.0%) |
| Former | 6,625,650 (29.3%) | 127,975 (48.7%) |
| Current | 4,031,225 (17.8%) | 29,985 (11.4%) |
| Not known | 1,207,285 (5.3%) | 2,420 (0.9%) |
| Hypertension | 3,680,020 (16.3%) | 112,735 (42.9%) |
| Type 2 diabetes mellitus | 1,314,575 (5.8%) | 37,475 (14.3%) |
| Coronary heart disease | 841,100 (3.7%) | 31,440 (12.0%) |
| Previous stroke or TIA | 479,110 (2.1%) | 14,175 (5.4%) |
| Heart failure | 112,325 (0.5%) | 10,605 (4.0%) |
| Chronic kidney disease | 1,147,155 (5.1%) | 62,125 (23.6%) |
| Depression | 3,796,810 (16.8%) | 54,410 (20.7%) |
| Diuretic at baseline | 1,359,115 (6.0%) | 66,495 (25.3%) |
| Tophaceous gout | N/A | 2,240 (0.9%) |
| Serum urate at diagnosis, micromol/L (SD) | N/A | 464 (98.7) |
| Not known | N/A | 108,340 (41.2%) |

**Supplementary Table S1.** Baseline demographics and comorbidities of the reference population and individuals with incident gout diagnoses between 1 July 2016 and 30 June 2025. Data are shown at the time of diagnosis for individuals with gout and at cohort entry for the reference population. Counts have been rounded to the nearest 5 to reduce the risk of disclosure; as such, column totals may differ from the sum of the individual variables. Body mass index represents the most recently recorded value, assuming this was within 10 years. Diuretic use at baseline was defined as one or more prescriptions for a diuretic medication in the 6 months before diagnosis (for individuals with gout) or cohort entry (for the reference population). SD: standard deviation; TIA: transient ischaemic attack; N/A: not applicable.

| **Outcome** | **Analysis** | **Change in trend, percentage points/year** | **95% confidence intervals** | **p-value** |
| --- | --- | --- | --- | --- |
| At least one serum urate measurement at baseline | NW lag 5 (primary) | 1.45 | 0.57 to 2.33 | 0.001 |
|  | NW lag 3 | 1.45 | 0.53 to 2.37 | 0.002 |
|  | NW lag 12 | 1.45 | 0.69 to 2.21 | <0.001 |
|  | Season adjusted | 1.65 | 0.52 to 2.78 | 0.005 |
|  | COVID adjusted | 0.09 | -0.93 to 1.11 | 0.863 |
| ULT within 12 months of diagnosis | NW lag 5 (primary) | -0.76 | -1.17 to -0.35 | <0.001 |
|  | NW lag 3 | -0.76 | -1.18 to -0.34 | <0.001 |
|  | NW lag 12 | -0.76 | -1.09 to -0.43 | <0.001 |
|  | Season adjusted | -0.77 | -1.18 to -0.36 | <0.001 |
|  | COVID adjusted | -0.91 | -1.35 to -0.46 | <0.001 |
| ULT within 12 months of risk factor onset | NW lag 5 (primary) | -1.07 | -1.67 to -0.47 | <0.001 |
|  | NW lag 3 | -1.07 | -1.72 to -0.43 | 0.001 |
|  | NW lag 12 | -1.07 | -1.50 to -0.65 | <0.001 |
|  | Season adjusted | -1.05 | -1.47 to -0.63 | <0.001 |
|  | COVID adjusted | -1.21 | -1.94 to -0.47 | 0.001 |
| High-dose first ULT prescription | NW lag 5 (primary) | 0.62 | 0.29 to 0.96 | <0.001 |
|  | NW lag 3 | 0.62 | 0.29 to 0.96 | <0.001 |
|  | NW lag 12 | 0.62 | 0.27 to 0.98 | <0.001 |
|  | Season adjusted | 0.62 | 0.23 to 1.00 | 0.002 |
|  | COVID adjusted | 0.61 | 0.16 to 1.05 | 0.008 |
| Co-prescription of prophylaxis with ULT | NW lag 5 (primary) | -0.17 | -0.59 to 0.25 | 0.428 |
|  | NW lag 3 | -0.17 | -0.64 to 0.30 | 0.471 |
|  | NW lag 12 | -0.17 | -0.48 to 0.14 | 0.274 |
|  | Season adjusted | -0.26 | -0.61 to 0.08 | 0.133 |
|  | COVID adjusted | -0.08 | -0.54 to 0.38 | 0.721 |
| Ongoing ULT prescription at 12 months following initiation | NW lag 5 (primary) | -0.48 | -1.03 to 0.07 | 0.086 |
|  | NW lag 3 | -0.48 | -1.01 to 0.05 | 0.076 |
|  | NW lag 12 | -0.48 | -0.98 to 0.02 | 0.060 |
|  | Season adjusted | -0.57 | -1.23 to 0.10 | 0.093 |
|  | COVID adjusted | -0.35 | -1.01 to 0.32 | 0.304 |
| At least one serum urate measurement within 12 months of ULT initiation | NW lag 5 (primary) | 0.91 | 0.40 to 1.42 | <0.001 |
|  | NW lag 3 | 0.91 | 0.38 to 1.43 | <0.001 |
|  | NW lag 12 | 0.91 | 0.44 to 1.38 | <0.001 |
|  | Season adjusted | 0.94 | 0.26 to 1.63 | 0.008 |
|  | COVID adjusted | 0.59 | -0.02 to 1.20 | 0.057 |
| Two or more serum urate measurements within 12 months of ULT initiation | NW lag 5 (primary) | 0.69 | 0.11 to 1.27 | 0.021 |
|  | NW lag 3 | 0.69 | 0.12 to 1.26 | 0.019 |
|  | NW lag 12 | 0.69 | 0.12 to 1.26 | 0.018 |
|  | Season adjusted | 0.69 | -0.004 to 1.39 | 0.051 |
|  | COVID adjusted | 0.54 | -0.22 to 1.29 | 0.162 |
| At least one serum urate level <360 µmol/L within 12 months of ULT initiation | NW lag 5 (primary) | 1.25 | 0.64 to 1.86 | <0.001 |
|  | NW lag 3 | 1.25 | 0.66 to 1.84 | <0.001 |
|  | NW lag 12 | 1.25 | 0.61 to 1.89 | <0.001 |
|  | Season adjusted | 1.28 | 0.59 to 1.98 | <0.001 |
|  | COVID adjusted | 0.95 | 0.27 to 1.64 | 0.007 |
| Serum urate <360 µmol/L among those with at least one urate measurement within 12 months of ULT initiation | NW lag 5 (primary) | 1.19 | 0.46 to 1.93 | 0.002 |
|  | NW lag 3 | 1.19 | 0.44 to 1.94 | 0.002 |
|  | NW lag 12 | 1.19 | 0.44 to 1.95 | 0.002 |
|  | Season adjusted | 1.22 | 0.44 to 2.00 | 0.003 |
|  | COVID adjusted | 0.97 | 0.21 to 1.73 | 0.013 |
| Repeat serum urate level recorded within 6 to 12 months of urate target attainment | NW lag 5 (primary) | 1.10 | 0.12 to 2.08 | 0.029 |
|  | NW lag 3 | 1.10 | 0.16 to 2.04 | 0.022 |
|  | NW lag 12 | 1.10 | 0.28 to 1.91 | 0.009 |
|  | Season adjusted | 1.13 | 0.25 to 2.00 | 0.012 |
|  | COVID adjusted | 1.79 | 0.59 to 3.00 | 0.004 |
| Serum urate remained <360 µmol/L among those with a repeat measurement 6 to 12 months after target attainment | NW lag 5 (primary) | -0.53 | -1.88 to 0.83 | 0.443 |
|  | NW lag 3 | -0.53 | -1.94 to 0.89 | 0.462 |
|  | NW lag 12 | -0.53 | -1.67 to 0.62 | 0.366 |
|  | Season adjusted | -0.42 | -1.84 to 1.01 | 0.563 |
|  | COVID adjusted | -0.76 | -2.66 to 1.14 | 0.430 |

**Supplementary Table S2.** Sensitivity analyses comparing differences in trends in outcomes for individuals with gout before and after publication of the NICE gout management guideline. Estimates were obtained from single-group interrupted time-series analyses. Trend differences (comparing the periods before and after June 2022) are reported as absolute percentage-point change per year. Primary models used Newey-West (NW) standard errors with 5 lags. Sensitivity analyses used alternative NW lag specifications (3 and 12 lags), adjustment for calendar month, and adjustment for the COVID-19 pandemic period (using an indicator for March 2020 to February 2022).

**Supplementary Data:** Codelists and outcome definitions

***Index gout diagnosis definition***

The index diagnosis date for gout was defined as the first recorded incident diagnostic code for gout in an individual’s primary care record or a hospital admission with gout recorded as the primary discharge diagnosis. To reduce the likelihood of prevalent cases being misclassified as incident cases, individuals were required to have at least 12 months of continuous registration before the index diagnostic code. Individuals with prescriptions for urate-lowering therapy (ULT) more than 30 days before their index diagnostic code were not considered to have an incident diagnosis.

- ***Incident gout diagnosis in primary care (SNOMED):***

<https://www.opencodelists.org/codelist/user/markdrussell/gout/75b2996a/>

- ***Hospitalisations for gout (ICD-10):*** <https://www.opencodelists.org/codelist/user/markdrussell/gout-admissions/296966be/>

***Gout flare definition:***

Criteria for defining gout flares were adapted from a previously published definition^1^, as any of the following:

1) The presence of a non-index diagnostic code for gout flare, using a dedicated flare codelist:

<https://www.opencodelists.org/codelist/user/markdrussell/gout-flaresattacks/78655960/>

2) A non-index hospital admission with gout recorded as the primary discharge diagnosis:

<https://www.opencodelists.org/codelist/user/markdrussell/gout-admissions/296966be/>

3) A non-index emergency department attendance with gout recorded as the primary diagnosis:

<https://www.opencodelists.org/codelist/user/markdrussell/gout/75b2996a/>

4) A non-index gout diagnostic code with a prescription for a flare treatment (colchicine, NSAID or corticosteroid) issued on the same day:

<https://www.opencodelists.org/codelist/user/markdrussell/gout/75b2996a/>

<https://www.opencodelists.org/codelist/user/markdrussell/colchicine-dmd/161e18be/>

<https://www.opencodelists.org/codelist/nhs-drug-refsets/oralnsaiddrug_cod/004e8c52/>

<https://www.opencodelists.org/codelist/nhs-drug-refsets/c19corstedrug_cod/41d73ef1/>

Events occurring within 14 days of one another were excluded to avoid double counting of the same flare.

***Baseline characteristics and comorbidity definitions***

Baseline characteristics were assessed at the index diagnosis date for individuals with incident gout and at cohort entry for the reference population. Unless otherwise specified, a comorbidity was considered present where a relevant diagnostic code had been recorded on or before the baseline date; a comorbidity was assumed not to be present if the relevant diagnostic code was absent from the medical record.

- ***Ethnicity***: obtained from coded primary care records, supplemented using data from secondary care admission records where primary care ethnicity records were unavailable:

<https://www.opencodelists.org/codelist/opensafely/ethnicity-snomed-0removed/22911876/>

- ***Body mass index:*** defined using the most recently recorded SNOMED code (60621009, 846931000000101) for BMI within the preceding 10 years, provided the individual was aged 16 years or older at the time of measurement. Categories were defined as underweight (<18.5 kg/m²), normal weight (18.5-24.9 kg/m²), overweight (25.0-29.9 kg/m²), obesity class I (30.0-34.9 kg/m²), class II (35.0-39.9 kg/m²), and class III (≥40.0 kg/m²).
- ***Smoking status:*** most recently recorded smoking status code, categorised as never, former or current smoking:

<https://www.opencodelists.org/codelist/opensafely/smoking-clear/2020-04-29/>

- ***Hypertension:*** Any recorded diagnosis of hypertension on or before baseline:

<https://www.opencodelists.org/codelist/nhsd-primary-care-domain-refsets/hyp_cod/521a90af/>

- ***Type 2 diabetes mellitus***: Any recorded diagnosis of type 2 diabetes mellitus on or before baseline:

<https://www.opencodelists.org/codelist/nhsd-primary-care-domain-refsets/dmtype2audit_cod/130b114a/>

- ***Coronary heart disease***: Any recorded diagnosis of coronary heart disease on or before baseline:

<https://www.opencodelists.org/codelist/nhsd-primary-care-domain-refsets/chd_cod/0f32f87c/>

- ***Heart failure:*** Any recorded diagnosis of heart failure on or before baseline:

<https://www.opencodelists.org/codelist/nhsd-primary-care-domain-refsets/hflvsd_cod/31a633c6/>

- ***Depression:*** Any recorded diagnosis of depression on or before baseline: <https://www.opencodelists.org/codelist/nhsd-primary-care-domain-refsets/depr_cod/477e1261/>
- ***Previous stroke or transient ischaemic attack***: Any recorded diagnosis of stroke or transient ischaemic attack on or before baseline:

<https://www.opencodelists.org/codelist/nhsd-primary-care-domain-refsets/strk_cod/78779ff3/>

<https://www.opencodelists.org/codelist/nhsd-primary-care-domain-refsets/tia_cod/1625df86/>

- ***Chronic kidney disease:*** CKD stages 3 to 5 were defined as a recorded CKD diagnostic code on or before baseline and/or an eGFR below 60 mL/min/1.73 m² based on the baseline creatinine measurement. The baseline measurement was the measurement closest to the index diagnosis within 2 months before or after diagnosis:

<https://www.opencodelists.org/codelist/nhsd-primary-care-domain-refsets/ckdatrisk2_cod/4529f7b4/>

<https://www.opencodelists.org/codelist/nhsd-primary-care-domain-refsets/cre_cod/48c03197/>

- ***Diuretic use***: One or more prescriptions for a diuretic medication in the 6 months before baseline:

<https://www.opencodelists.org/codelist/user/markdrussell/diuretics-dmd/63ff3253/>

- ***Tophaceous gout***: Any recorded diagnosis of tophaceous gout on or before the index gout diagnosis:

<https://www.opencodelists.org/codelist/user/markdrussell/gouty-tophi/1174cc96/>

- ***Baseline serum urate:*** The serum urate measurement closest to the index diagnosis date within 2 months before or after diagnosis, provided that the measurement occurred before ULT initiation:

<https://www.opencodelists.org/codelist/user/markdrussell/serum-urate/6208a621/>

***Outcome definitions***

- ***Serum urate at diagnosis:*** At least one serum urate measurement within 2 months before or after the index diagnosis, provided that the measurement occurred before ULT initiation:
- ***ULT initiation within 12 months of diagnosis***: At least one newly recorded prescription for allopurinol or febuxostat within 12 months of the index gout diagnosis:

<https://www.opencodelists.org/codelist/user/markdrussell/allopurinol-dmd/2c7acbfd/>

<https://www.opencodelists.org/codelist/user/markdrussell/febuxostat-dmd/7dcad94c/>

- ***ULT initiation within 12 months of onset of risk factors indicating that ULT should be offered***: Newly recorded prescription for allopurinol or febuxostat within 12 months of the onset of at least one NICE-specified indication to be offered ULT, as follows: at least one additional gout flare (defined above) after diagnosis, CKD stages 3 to 5 (defined as the earliest recorded CKD diagnostic code or eGFR below 60 mL/min/1.73 m²), diuretic use, chronic gouty arthritis (codelist below) or tophaceous gout (defined above). For risk factors developing after gout diagnosis, the first qualifying date was used as the risk-factor onset date. For individuals with risk factors present at gout diagnosis, the date of gout diagnosis was used as risk factor onset date:

<https://www.opencodelists.org/codelist/user/markdrussell/gout-chronic-arthritis/14278c8c/>

- ***First ULT medication:*** Whether the first recorded prescription for ULT after the index gout diagnosis was for allopurinol or febuxostat.
- ***High-dose ULT initiation***: Whether the first recorded prescription for ULT after the index gout diagnosis was allopurinol 300 mg tablets or febuxostat 120 mg tablets:

<https://www.opencodelists.org/codelist/user/markdrussell/allopurinol-300mg-doses-dmd/7a5be592/>

<https://www.opencodelists.org/codelist/user/markdrussell/febuxostat-120mg-doses-dmd/00161664/>

- ***Febuxostat initiation in individuals with major cardiovascular disease***: Initiation of febuxostat in an individual with a pre-existing record of coronary heart disease, stroke or transient ischaemic attack (as defined above).
- ***Gout flare prophylaxis during ULT initiation:*** Prescription of colchicine, an NSAID or an oral corticosteroid (codelists defined above) on the same day as the first ULT prescription, with a further prescription for the same prophylactic medication 3 months (+/-30 days) later.
- ***ULT persistence at 12 months***: A further prescription for allopurinol or febuxostat at 12 months (+/- 60 days) after their initial ULT prescription.
- ***Serum urate monitoring after ULT initiation***: The proportion of patients with at least one recorded serum urate measurement within 12 months after ULT initiation, and the proportion with two or more serum urate measurements within 12 months after ULT initiation.
- ***Serum urate target attainment***: The proportion of patients starting ULT who had at least one recorded serum urate concentration below 360 µmol/L within 12 months after ULT initiation.
- ***Repeat serum urate monitoring after target attainment***: At least one further serum urate measurement recorded 6 to 12 months after first achieving a serum urate concentration below 360 µmol/L.
- ***Maintenance of serum urate target***: Serum urate remained below 360 µmol/L on the first repeat serum urate measurement 6 to 12 months after initial target attainment.
- ***Treatment of gout flares***: A prescription for colchicine, an NSAID, or an oral corticosteroid on the same day as a recorded gout flare (defined above) occurring at least 14 days after diagnosis.
